# Type I IFN-activated lung monocytes and macrophages as initiators and drivers of fibrosis at the alveolar barrier in IPF

**DOI:** 10.1101/2025.05.09.25326523

**Authors:** Vishal Nathwani, Praveen Weeratunga, Jeongmin Woo, Laura Denney, Chaitanya Vuppusetty, Tian Hu, Sabrina Zulfikar, Andrew Achaiah, Harriet Parker, Huei-Wen Chuang, Michalina Mazurczyk, Aleksandr Fedorov, Alison Simmons, Colin Clelland, Jan Rehwinkel, Agne Antanaviciute, Ling-Pei Ho

**Affiliations:** MRC Translational Immune Discovery Unit, Weatherall Institute of Molecular Medicine, University of Oxford, UK; Chinese Academy of Medical Science Oxford Institute, University of Oxford, Oxford, UK; MRC WIMM Imaging Facility, Weatherall Institute of Molecular Medicine, University of Oxford, UK; Respiratory Medicine Unit, Nuffield Department of Medicine, University of Oxford, Oxford UK

## Abstract

Idiopathic pulmonary fibrosis (IPF) is a progressive and fatal lung disease with limited therapeutic options. Although macrophages have been implicated in pathogenesis in murine models, their role in human disease remains unclear. Here, we present a comprehensive, unbiased single-cell transcriptomic and regulon atlas of myeloid cells from the alveolar barrier and lung tissue. We demonstrate that alveolar barrier macrophages are transcriptionally distinct from their lung tissue counterparts, exhibiting a striking upregulation of type I interferon (IFN) signalling in IPF. Circulating monocytes from IPF patients are primed for type I IFN responses, particularly in early disease, and monocytes are enriched with type I IFN-associated genes at lung sites of early fibrosis. These findings suggest a central role for type I IFN-activated monocytes in the initiation of fibrosis, and for alveolar barrier-related, type I IFN-activated macrophages in perpetuating fibrosis. These insights provide a rationale for therapeutically targeting monocytes in early stages of IPF.

## Introduction

Idiopathic pulmonary fibrosis (IPF) is a progressive and fatal form of lung fibrosis, with a median survival of 3–5 years despite the advent of anti-fibrotic therapies ^1^. These treatments only modestly slow disease progression and are often associated with significant off-target effects, leading to treatment discontinuation in up to 45% of patients ^2^. A deeper understanding of IPF pathogenesis and the identification of new therapeutic targets remain urgent priorities.

Immune cells are increasingly recognised as key contributors to pathogenesis of IPF^3^. In particular, macrophages and monocytes have been implicated in the pathogenesis of lung fibrosis in murine studies^4, 5, 6^. Depletion of monocyte-derived macrophages in such models has been shown to attenuate fibrosis ^4, 5^. In human studies, elevated levels of circulating monocytes in IPF have been associated with disease severity and poorer outcomes ^7, 8, 9^.

In the lung, monocytes differentiate into macrophages that are essential for immune homeostasis, tissue maintenance, and repair. Under steady-state conditions, self-renewing alveolar macrophages, derived from yolk sac progenitors, protect alveolar structures with support from colony stimulating factor 2 (CSF2) secreted by alveolar type II epithelial cells^10^. Other macrophage populations in the lung interstitium can be found in peri-bronchial and peri-vascular sites, where they regulate the immune activity around endothelial cells and smaller bronchi^11,12^.

During infection, circulating monocytes infiltrate the lungs and either reside transiently and die, or assimilate into the long term alveolar space and interstitium, acquiring self renewal capacity in the process^13^. An added layer of complexity imbues the macrophage landscape in chronic diseases like IPF, as monocyte-derived macrophages are surrounded by and can interact with a broader array of immune and stromal cells not typically present in healthy lungs. Much less is known of the monocyte and macrophage composition and alteration in their transcriptomic states in such chronically diseased lungs.

Previous single cell transcriptomic studies in IPF have emphasised the presence of a pro-fibrotic macrophage population in lung tissue digests - variously labelled ‘pro-fibrotic macrophages’ (Adams, Ayaub)^14, 15^, ‘SPP1^+^ macrophages’ (Morse)^16^, ‘PLA2G7^+^ macrophages’ (Wang)^17^ and SPP1, MMP7-expressing macrophages (Reyfman)^18^. However, it is likely that there are other macrophage and monocytic subsets and/or states in the fibrotic lung which have not been defined. In addition, these studies do not differentiate between monocytes and macrophages found in the interstitium of the lungs and those in the alveolar space. Studies in the bronchoalveolar space will provide insight into the immune activity at the barrier of the lungs, and initiating events here, which are likely different from that in the interstitium.

In this study, we use single cell RNA sequencing of cells in the bronchoalveolar space paired with peripheral blood mononuclear cells, and supplemented by single cell transcriptomic data from lung digest cells to examine the transcriptomic states, composition and potential pathogenic molecular pathways in monocytes and macrophages in IPF lungs. We identify a macrophage profile in the alveolar space that is distinct from lung tissue-derived macrophages, characterized by a striking upregulation of type I IFN transcriptional program in IPF. Upregulation of type 1 IFN signalling was not observed in lung tissue-based macrophages. Additional analyses—supported by phospho-protein assays and immunofluorescence—demonstrate a link between type I IFN-activated monocytes and early stages of disease. Our findings, from different compartments, contributed by 54 different IPF patients, suggest a key role for type 1 IFN-activated monocytes in initiating fibrosis in IPF, and involvement of barrier-related type-1 activated macrophages in driving chronic fibrosis.

## Results

### Macrophages in alveolar space are defined by with distinct homeostatic and pro-inflammatory transcriptomic states at the alveolar barrier in IPF and HC

To gain an overall understanding of the transcriptomic activities of macrophages and monocytes at the alveolar barrier, we first conducted single cell RNA sequencing of cells obtained from the alveolar space and paired blood peripheral mononuclear cells (PBMCs) [n=3 IPF and n=3 controls with no lung disease (hereon, referred to as ‘healthy controls’ or HC), total single cells-26,686] (‘Oxford BAL cells’ and ‘Oxford PBMC’; Cohort A) (Figure 1A). This was supplemented by analysis of a single cell transcriptomic dataset from lung digest of explanted lungs and their controls (‘Morse lung digest’; n=3 each group, total single cells – 48,728)^16^.

**Figure 1.**
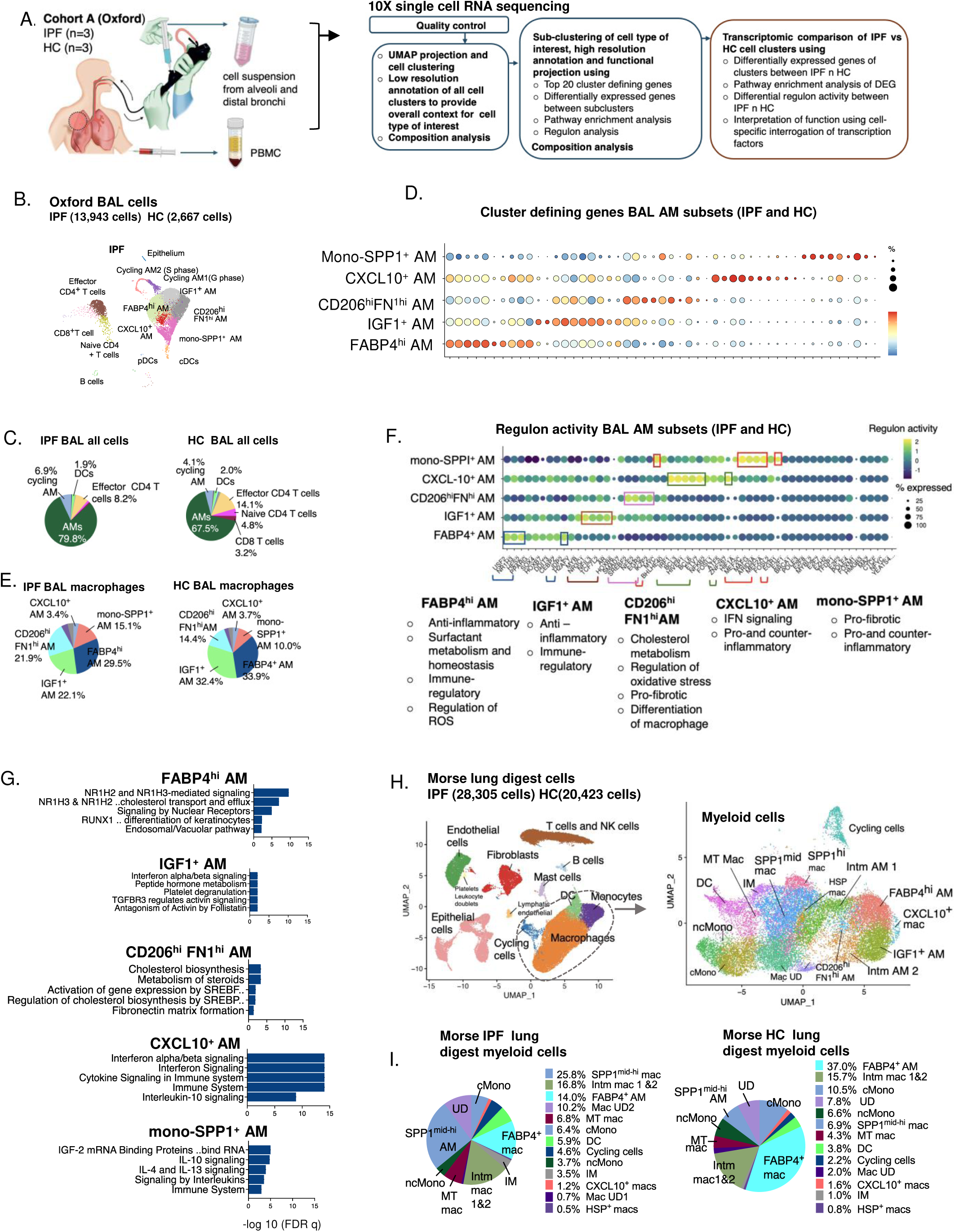
scRNAseq reveals macrophage landscape characterised by distinct homeostatic and pro-inflammatory transcriptomic states at the alveolar barrier. A. Overview of study and analysis pathway for Oxford Cohort A (BAL sampling). IPF – idiopathic pulmonary fibrosis, HC – healthy control. PBMC – peripheral blood mononuclear cells, BAL – bronchoalveolar lavage B. UMAP for BAL cells (IPF and HC combined); n=3 IPF patients and n=3 HC individuals. AM – alveolar macrophage C. Composition of cell types for IPF and HC BAL (as % of all cells) D. Top 5 cluster defining genes for AM subsets in IPF and HC. E. Composition of BAL alveolar macrophages (AMs) subsets (as % of all AMs, not including cycling AMs) F. Bubble plot showing regulon activity (colour scale) and % of cells in the AM subset that expressed the specified regulon. Bullet points shows functional interpretation of the most highly upregulated regulon activity (in coloured boxes). ‘% expressed’ – % of cell expressing specified regulon with regulon score >0.Regulon activity refers to average regulon activity for the cells in the AM subset and compared to other cell types by AUCell methods. G. Pathway enrichment analysis of DEGs amongst the five AM subsets in BAL (hypergeometric test; Reactome database). ‘NR1H3 & NR1H2. cholesterol transport and efflux’ term was shorted from NR1H3 & NR1H2 regulate gene expression linked to cholesterol transport and efflux; ‘RUNX1 ..differentiation of keratinocytes from RUNX1 regulates transcription of genes involved in differentiation of keratinocytes H. UMAP of publicly deposited lung digest single cell transcriptomic dataset from Morse et al 2019. Single cell dataset were analysed with same pipeline as in (A) and annotated, with a focus on myeloid cell subset (circled). Right panel - UMAP of sub-clustered myeloid cells in. Intm mac– intermediate macrophage; cMono-classical monocytes, ncMono – non-classical monocytes, TM – transitional monocyte-macrophages, Mac UD – undefined macrophages; IM – interstitial macrophages; DC – dendritic cells, MT mac – metallothionein macrophages I. Composition of myeloid cells in Morse’s lung digest as % of all myeloid cells for IPF and HC. Intm Mac 1 and 2; and SPP1mid and SPP1^hi^ macs are merged (due to similarity in transcriptomes)

Single cell RNA sequencing was performed using the 10X chromium platform and analysed according to workflow described in Figure 1A and previously^19, 20^. Demographic data are shown in Suppl Table 1.

17 cell clusters from BAL cells (IPF and HC combined) were identified using unsupervised clustering followed by uniform manifold approximation and projection (UMAP) plots (Figure 1B). First level clustering and annotation yielded 5 closely related clusters of alveolar macrophages (AM) amongst two DC subsets, and two CD4 T cell subsets, one CD8 T cell and B cell cluster, 2 cycling macrophage subsets, and one small cluster of epithelial cells (Figure 1B, Suppl Figure 1A). In both IPF and HC, macrophages dominate the immune cell landscape in the alveolar space (79.8% of IPF cells and 67.5% of HC cells in BAL) followed by CD4 T cells, then cycling macrophages (Figure 1C). The composition of immune cells in IPF and HC were comparable (Figure 1C). We labelled the macrophages ‘alveolar macrophages’ (AMs) as they were derived from the alveolar space although this does not preclude these also being found in interstitium or distal airways.

AM cell clusters were annotated according to cluster defining genes (Figure 1D, Suppl Table 2) and in conjunction with findings from six other key publications (mostly in lung digests)^16, 21, 22, 23, 24, 25^ including the integrated lung health and disease dataset^26^ (Suppl Table 3A). AM cell clusters were termed ‘subsets’ but for some, ‘transcriptomic states’ are more likely and discussed later. The two previously published lung macrophage subsets in IPF (from lung *digest*)^16^ – FABP4^hi^ macrophage and SPP1^+^ macrophages can be identified in our BAL macrophage populations. SPP1^+^ macrophage subset was found within the cluster of cells that also contained cell types with markers for monocytes (CD14, S100 A8/9) (Suppl Table 2 and 3A). We labelled this cluster, mono-SPP1^+^ AM. This subset is likely monocytes that are differentiating to SPP1^+^ macrophages. Three subsets, hitherto undescribed in IPF lungs, were identified and termed CXCL10^+^ AM, IGF1^+^ AM and CD206^hi^ FN1^hi^ AM. CXCL10^+^ AMs have been described in COVID-19 lungs,^23^ as well as inflammatory disease like rheumatoid arthritis and inflammatory bowel disease^23^. IGF1^+^ AM uniquely expressed *IGF1* within a population that also displayed key genes of the resident FABP4^hi^ AM subset. A population with a similar transcriptomic profile to IGF1^+^ AM (Suppl Table 3A) has also been observed in healthy volunteer BAL samples^22^. CD206^hi^FN1^hi^ AMs have recently been described in our imaging mass cytometry study in IPF as a cell type found co-located with aberrant alveolar intermediate cells^27^. All AM subsets or states were found in both control and IPF BAL. In both HC and IPF, FABP4^hi^ AM and IGF1^+^ AM formed the majority of AMs (66.2% of AMs in HC and 51.6% in IPF). There is a small increase in mono-SPP1^+^ AM (15.1% of myeloid cells in IPF vs 10.0% HC) and CD206^hi^FN1^hi^ AM (21.9% v 14.4%) in IPF, while IGF1^+^ AM showed lower frequency (22.1% vs 32.4%) (Figure 1E).

Findings from pathway enrichment analysis (PEA) of the AM subsets and their gene regulatory network analysis (regulons) (using SCENIC (Single-Cell rEgulatory Network Inference and Clustering)^28^ demonstrated likely functional transcriptomic changes that supported each other (Figure 1F-G). Thus, FABP4^hi^ AMs expressed genes and regulons that reflect homeostatic lipid metabolic function and physiological control of inflammation; IGF1^+^ AM - anti-inflammation; CD206^hi^FN1^hi^ - lipid metabolism, regulation of oxidative stress and inflammation, and CXCL10^+^ AMs - IFN-signalling and pro-inflammatory processes (Figure 1F, Suppl Table 3C). Mono-SPP1^+^ AM showed a mixed functional signature in keeping with this being a mixed cell cluster of monocytes and SPP1^+^ AMs – upregulation of both pro-fibrotic, pro-inflammatory and anti-inflammatory processes. Overall, AM subsets can be functionally categorised into those with homeostatic lipid metabolic activities (FABP4^+^ AM and CD206^hi^FN1 ^hi^ AMs) and those that do not (mono-SPP1^+^ AMs, CXCL10^+^ AM and IGF1^+^AM). Amongst these, subsets with greatest evidence of cytokine signalling were the CXCL10^+^ AMs.

To determine if these AM subsets were also found in lung tissue digest, we used the Morse publicly deposited lung cell single cell transcriptomic dataset^16^. Data underwent the same QC and analysis as our BAL dataset to allow direct comparison. 13 subsets of myeloid cells were identified based on cluster-defining genes and in conjunction with the same references as for BAL cells (Figure 1H, Suppl Fig 1B, Suppl Table 4A). Of these, 8 were macrophage subsets– FABP4^hi^ macrophages, SPP1^+^ macrophages, HSP^+^ mac, intermediate AMs (intmMac), transitional AMs (TM), CXCL10^+^ macrophages, Metallothionein macrophage (MT mac) and interstitial macrophages (IM) (Figure 1H). A full exposition of these annotations are presented in Suppl Table 4C. All 5 AMs from the Oxford BAL sample were found in Morse. SPP1^+^ macrophages were the dominant subset in this IPF lung digest as shown before^16, 24^, increased at the expense of FABP4^+^ macs. However, the defining genes were different from that found in mono-SPP1^+^ AMs in the BAL, possibly due to presence of differentiating monocytes in the latter. CD206^hi^ FN1^hi^ AM and CXCL10^+^ AMs were less distinct in Morses’ lung digest data (Figure 1I). This lung digest sample also contained a population of monocytes which were not evident in BAL – non-classical monocytes (ncMono) and classical monocytes (cMono) made up 10.1% of lung digest’s myeloid population (Figure 1I). This could be due to presence of lung -based monocytes and/or circulating monocytes in the lung digest.

In summary, single cell transcriptomic analyses of macrophages at the alveolar barrier revealed a cellular landscape which is distinct from the lung tissue and interstitium. In contrast to lung digest, only five transcriptomic states and/or subsets of AMs were observed in the alveolar space. These can be classified into two functional groups, one consisting of FABP4^hi^ AM and CD206^hi^FN1^hi^ AMs which expressed high homeostatic lipid metabolic functional and regulon gene activities indicating a housekeeping and clearance function amongst their roles, while the second group, CXCL10^+^ AM, mono-SPP1^+^ AM and IGF1^+^ AMs showed more prominent inflammatory and counter-inflammation cytokine signalling gene profile.

### Increased type 1 IFN activity in CXCL10^+^ AM and IGF1^+^ AMs defines key macrophage activity at the alveolar barrier in IPF

We next compared IPF and HC AMs (hereon termed ‘IPF AM’ and ‘HC AM’), to identify IPF-specific transcripts and regulatory networks.

There was only a short list of differentially expressed genes (DEG) between IPF and HC AMs (Figure 2A). At single gene level, there was striking upregulation of FN1 in CD206^hi^ FN1^hi^ AM, CXCL10^+^ AM and FABP4^+^ AM; and CXCL10 in mono-SPP1^+^ AM and IGF1^+^ AM. CCL-18, previously shown to be high in IPF BAL fluid^29^, was highly upregulated in IPF’s FABP4^+^ AM (Figure 2A).

**Figure 2.**
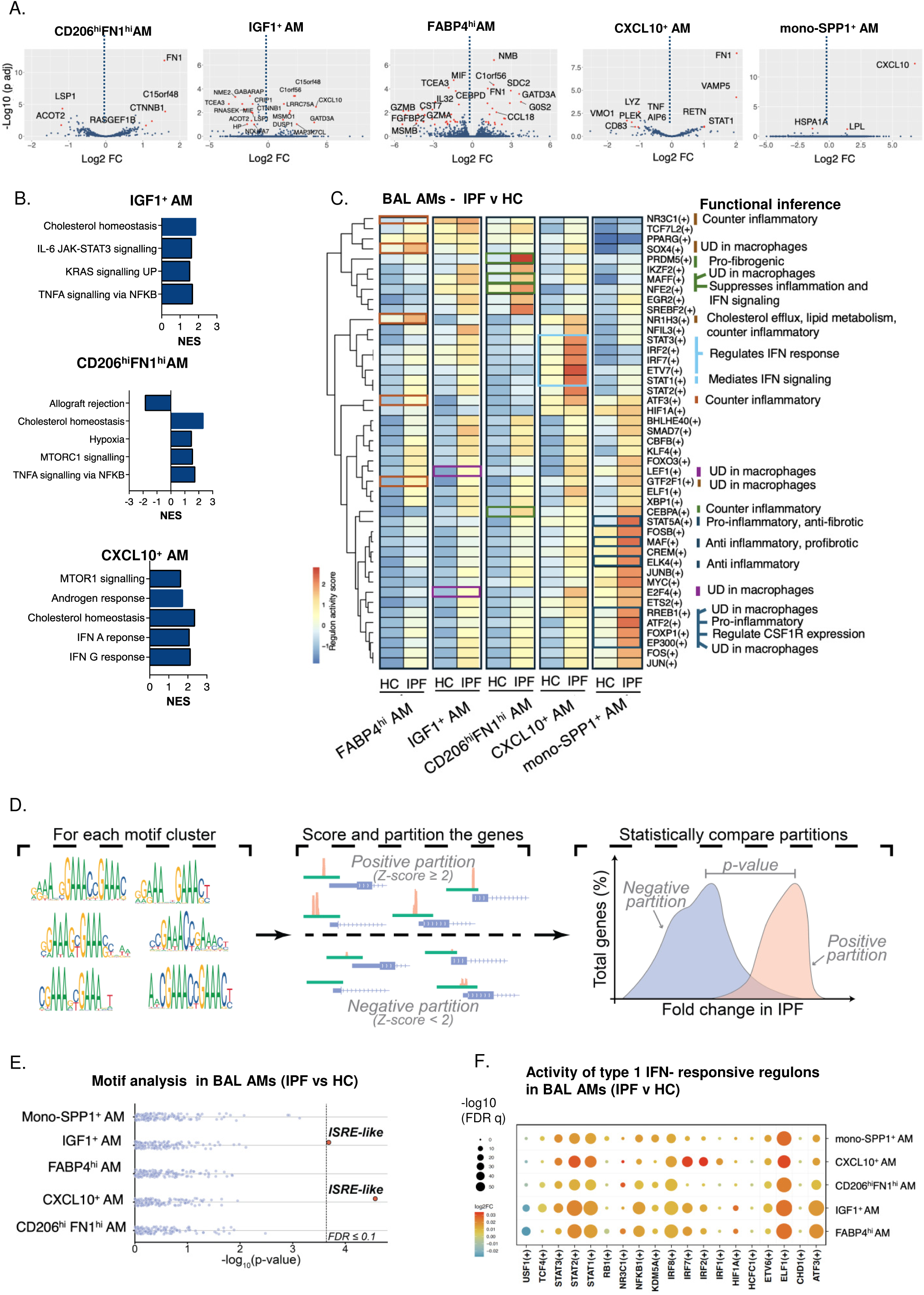
Increased inflammatory and type I IFN signalling activities in macrophages are disease-specific abnormalities at the IPF alveolar barrier. A. Volcano plots showing differentially expressed genes for BAL AM subsets, comparing IPF and HC. Red dots – genes with FDR q <0.1 and Log2FC>1.0 or < −1.0. B. Enriched gene sets in the three AMs with gene sets showing significant normalised enrichment scores (NES). No significant enrichment was found in mono-SPP1^+^ AMs and FABP4^hi^ AMs comparing IPF to HC. Gene sets from Hallmark database, analyzed using GSEA due to small numbers of DEGs C. Regulon activity analysis comparing IPF to HC; heatmap shows union of top 20 differential regulon activity (adj p< 0.05 and Log2FC >0.02). Boxes capture differentially expressed regulons (between IPF and HC) that are only seen in the specified AM subset with macrophage-specific function collated from literature search (Suppl Figure 3B). UD – undefined (as no publications in macrophages). D. Overview of the motif analysis workflow. Each motif cluster from the JASPAR database was used to score annotated genomic promoters, and the resulting cluster scores were subsequently Z-normalized. Then, for each cell population, we retained only genes whose estimated fold change between IPF and HC exceeded their standard error. Next, for each motif cluster (left), we categorized genes whose promoters had a motif cluster Z-score ≥ 2 as a positive partition (putatively regulated, middle top) and those with Z-score < 2 as the negative partition (baseline group, middle bottom). The average fold change in expression between IPF and HC (right, horizontal axis) for the genes in the positive partition was then compared with the negative partition (baseline) to determine whether motif presence was significantly associated with transcriptional changes in IPF relative to HC samples (Wilcoxon signed-rank test). E. Significance of the association between transcriptomic changes in IPF and motif enrichment in promoters of genes. Each dot represents a cluster of motifs from the JASPAR Core database, x axis - significance of the association between motif enrichment and gene expression in for each AM subset (Wilcoxon rank signed test; see also Suppl Figure 2Aa). Only genes containing ISRE-like motifs in IGF1⁺ and CXCL10⁺ AM subsets were significantly associated with upregulated expression in IPF relative to HC F. Bubble plot showing difference in type I IFN –responsive regulons in IPF vs HC BAL AMs. List of regulons activated after stimulation with type 1 IFN was derived from Mostafavi S et al 2016. Greatest number of significantly increased regulon activity in IPF were in CXCL10^+^ AMs

Only IGF1^+^ AM, CD206^hi^FN1^hi^ AM and CXCL10^+^ AMs showed significant enrichment of genes in pathway analysis between IPF and HC AMs. Gene set enrichment analysis (GSEA) found enrichment of proinflammatory gene sets in all three IPF AMs - ‘IL-6-JAK-STAT3’ and ‘TNFA signalling via NFKB’ in IGF1^+^ AM, ‘IFNG response’ and ‘IFNA response’ in CXCL10^+^ AM, and ‘TNFA via NFKB signalling’ in CD206^hi^FN1^hi^ AM (Figure 2B, Suppl Table 5A). For all three AMs, genes associated with cholesterol homeostasis were also enriched in IPF compared to HC AMs, potentially reflecting increased phagocytosis of surfactant from degrading type II alveolar epithelial cells and their catabolism^30, 31^. Genes associated with MTORC1, a master regulator of cell metabolism and cell growth, were increased in IPF’s CXCL10^+^ AM and CD206^hi^ FN1^hi^ AM (Figure 2B).

Examining the difference in regulon activities between IPF and HC AMs, three findings stood out. Firstly, there was an overall increase in transcription activities in all IPF AMs compared to HC AMs, pointing to a general increase in activity in IPF AMs. Secondly, all the IPF AM subsets showed increased BHLHE40, CBFB, SMAD7, TCF7L2 and FOS regulon activities in the top 20 differentially regulon activity list (references provided in Suppl Table 5B-C). These regulons reflect increase in inflammatory (BLHE40, SMAD7) and corresponding counter-inflammatory (FOS) activities, self-renewal (CBFB) and differentiation to M2-like macrophages (TCF7L2) (Suppl Table 5B-C). Thirdly, amongst these differentially expressed regulons, between 2 to 5 subset-specific regulons could be identified for each AM subset, providing insight into how different AM subsets might contribute to disease in IPF (Figure 2C and Suppl Table 5B-C). Here, we found that FABP4^hi^ AM, the subset- and IPF-specific regulons were anti-inflammatory; in CD206^hi^FN1^hi^AM –pro-repair/fibrotic, anti-inflammatory and anti-IFN signalling; and CXCL10^+^ AM – IFN signalling and counter-IFN signalling. Mono-SPP1^+^ AM’s upregulated regulons were associated with both pro- and anti (or counter)-inflammatory and pro- and anti-fibrotic activities. These data indicate increased activity in regulons involved in inflammatory, IFN signalling and fibrotic pathways in IPF AMs, strengthening the pathway enrichment findings in Figure 2B. Amongst these, CD206^hi^FN1^hi^ AM is unique in showing increased activity of gene regulatory network associated with fibrotic function (PRDM5) without a corresponding anti-fibrotic regulatory counterpart. IFN signalling activity was the most consistently upregulated pathway in IPF-at single gene, gene set and regulon level for CXCL10^+^ AMs.

The SCENIC pipeline infers a significant regulon activity when expression of transcription factors matches that of their target genes and related motifs. It treats each transcription factor as a separate entity even though they often act as a multiprotein complex (e.g. STAT1 and STAT2). It could therefore ‘over-predict’ involvement of individual regulons. To complement this we took another approach where we examined clusters of motifs provided by the JASPAR database^32^. Here, where possible, motif clusters broadly represents an entire pathway (e.g., ISRE-like motifs for Type I IFN response and GAS-like motifs for Type II IFN response). Starting with all genes expressed in IPF and HC AMs, we assessed if motif clusters (from the JASPAR database^32^ ^33^) are present in their promoters (Figure 2D). We then determined which motif statistically associates with transcriptional changes in IPF relative to HC samples for each of the AM subset. We observed a striking result. Amongst all the motifs in all AM subtypes, only one motif cluster was statistically associated with increased expression of its genes in IPF vs HC – the ISRE-like motifs in CXCL10^+^ AM and IGF1^+^ AMs (Figure 2E; Suppl Figure 2A-B). ISREs are IFN-stimulated response elements or motifs found in the promoters of type I IFN-responsive genes (ISGs). These findings indicate that amongst the differential regulon activities identified by the SCENIC pipeline, we can be most certain of the ISRE-like transcription machinery (motif-gene expression) as it showed the strongest statistical association with upregulation of gene expression in IPF. Importantly, the GAS-like (gamma-activated sequences) motifs, responsible for IFN-γ activation of target genes, were not enriched in this comparison (Figure 2D, Suppl Figur 2C-D), pinpointing the IFN signalling activity to type 1 rather than type 2 IFNs in CXCL10^+^ AM and the IGF1^+^ AM. An analysis of regulons associated with type 1 IFN stimulation (curated from Mostafavi et al ^34^) using the SCENIC pipeline provides further support for this finding (Figure 2F).

Overall, our data suggest that AMs at the alveolar barrier in IPF patients are transcriptionally and metabolically more active, with high level of regulatory activity related to inflammatory and IFN signalling pathways. Unbiased and comprehensive motif analysis provides greatest confidence for increased type 1 IFN activity in two populations of AMs (CXCL10^+^ AM and IGF1^+^ AMs) in IPF.

### Increase in type 1 IFN regulon activity in paired circulating classical monocytes in IPF patients

Having paired BAL and PBMC samples in the same patient provides a unique opportunity to determine if the lung findings are reflected in the circulating monocyte compartment. We first annotated the cell clusters using labels from our previous publication (COMBAT) and Nehar-Belaid et al ^35, 36^ (Figure 3A, Suppl Fig 3A). COMBAT phenotyped PBMC samples from SARS-CoV2 infected and healthy individuals and integrated Cite-Seq and scRNAseq to provide an enhanced annotation of immune cells. 10 cell clusters were identified, of which two were classical monocyte (cMono) and non-classical monocyte (ncMono) subsets. As shown previously^37^, monocytes made up a bigger proportion of PBMC in IPF compared to HC (Figure 3B). Re-clustering these two monocyte populations, 5 further subsets were found, and annotated as cMono. S100A8/9/12^hi^, cMono.HLA II^hi^, ncMono, mono.type1 IFN and monoDC (Figure 3C-E; Suppl Figure 3A-B) adopting COMBAT’s annotation style^36^. Proportion of cMono S100A8/9/12^hi^ (as % of all monocyte population) was higher in IPF compared to HC (Figure 3D). Mono.type 1 IFN subset (defined because of enrichment in type 1 IFN pathways in the cluster defining genes)(Suppl Figure 3B) made up only a small proportion of monocytes (less than 6%) in both IPF and HC, and there was no difference between the two groups.

**Figure 3.**
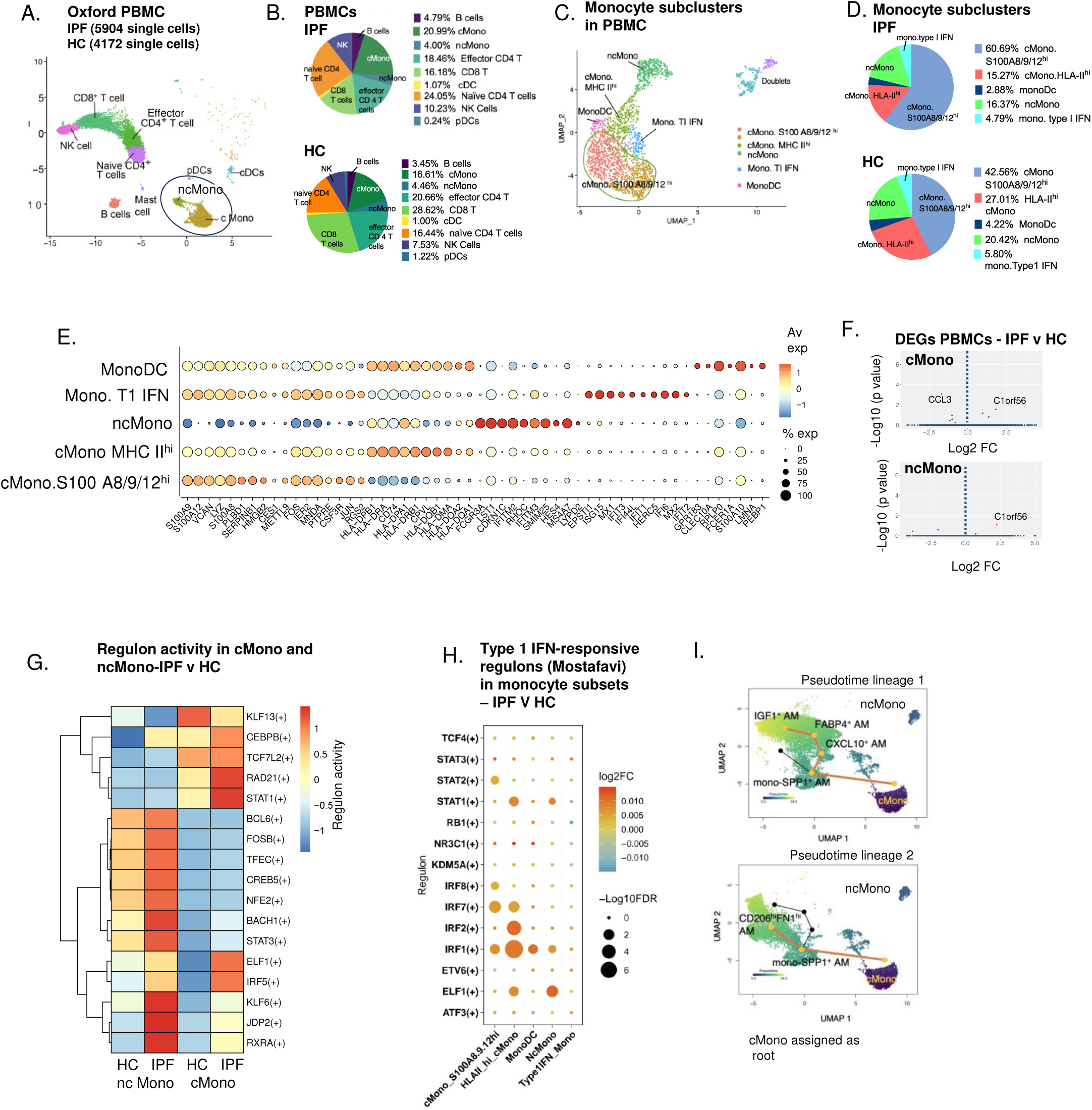
Small increase in type 1 IFN regulon activity in circulating classical monocytes is main transcriptomic difference between IPF and HC’s PBMC at single cell level. A. UMAP of PBMCs from IPF and HC (combined) at first level annotation. Monocytes are circled. B. Composition of PBMC (as % of all cells) for IPF and HC. Mast cells not included as <0.05% in both IPF and H C. Sub-clustering of monocytes reveals five monocyte subsets, annotated using COMBAT (see text) nomenclature D. Composition of monocyte subclusters (as % of all circulating monocytes) E. Top 5 cluster defining genes for monocyte subsets in PBMC (IPF and HC) F. Volcano plots showing differentially expressed genes for PBMC monocyte subsets, comparing IPF and HC. Red dots – genes with FDR q <0.1 and Log2FC>1.0 or < −1.0. G. Differential regulon activity (FDR q <0.05) for ncMono and cMono subtypes comparing IPF and HCs (no significant difference was found when the 5 subclustered monocyte subtypes were analysed) H. Fold change in expression for the type 1 IFN-specific regulons curated from Mostafavi in the 5 monocyte subtypes between IPF and HC. No significant difference was seen in the main ncMono and cMono cluster but in the subtypes, the two cMono subtypes showed strongest evidence of higher type 1 IFN regulon activity I. Slingshot trajectory inference for the matched BAL and blood samples for IPF patients from Oxford Cohort A, displaying two lineages of pseudotime. Directionality from cMono to mono-SPP1^+^ AM provided by biological inference that monocytes differentiate to macrophages.

At single cell gene expression level, we found almost no transcriptomic difference in monocytes between IPF and HC patients. Only one differentially expressed gene was identified (fold change >log2 1.0; FDR q <0.1) - *C1orf56* (unclear function) in cMono and ncMono (Figure 3F). Specifically, there was no difference in expression of any IFN signalling pathway related genes between IPF and HC.

Regulon analysis of all the PBMC cell types also revealed only very small differences between IPF and HC samples (10 fold less than observed in BAL cells) (Suppl Figure 4A) - there were no difference in regulon activity that was greater than 1.05 fold change. However, in the cMono and ncMono subsets in PBMCs, several regulons associated with type 1 IFN were statistically upregulated (Figure 3G, Suppl Fig 4A). Activity of several type I IFN-associated regulons in Mostafavi’s Type 1 IFN-associated regulons were also significantly increased in two subsets of classical monocyte (cMono.HLA II^hi^ and cMono.S100 A8/9/12^hi^), and to a lesser extent, also in the ncMono subsets in IPF (Figure 3H)^34^.

With this subtle but relevant finding, we applied a single cell trajectory analysis (Slingshot)^38^ to the explore how blood monocytes might be linked to BAL AMs. We selected Slingshot due to its specific design to detect multiple branching lineages in single-cell data and its accommodation of biological application of a starting point. Thus, since monocytes differentiate to macrophages, we added circulating monocyte clusters (PBMC’s ncMono and cMono) with AM subsets and fixed cMono as the start point (or ‘root’) in the trajectory. We identified two main trajectories - ‘lineage’ 1 which ‘travels’ from cMono to mono-SPP1^+^ AM, then CXCL10^+^ AMs, FABP4^+^ AM and IGF-1^+^ AM; and lineage 2, from cMono to mono-SPP1^+^ AM to CD206^hi^FN1^hi^ AM (Figure 3I). Befitting its likely monocyte-macrophage transitional and mix states, mono-SPP1^+^ AMs appear to be the first point in the lung after cMono and the branch point into CD206^hi^FN1^hi^ AM or CXCL10^+^ AMs. CD206^hi^FN1^hi^ AM and CXCL10^+^ AMs segregated distinctly while CXCL10^+^ AMs, IGF1^+^ AM and FABP4^hi^ AMs were more similar transcriptomically. Circulating ncMono did not contribute to these pseudotime lineages.

In summary, single cell RNA sequencing of monocytes did not show significant difference in the gene expression in monocyte subsets between IPF and HC. However, there was a small increase in regulon activity associated with type 1 IFN signalling in IPF’s classical monocyte subsets. Trajectory inference using paired BAL and blood monocytes proposes that circulating classical monocytes are most similar transcriptomically to mono-SPP1^+^ AMs at the alveolar barrier. Overall, the findings suggest that circulating classical monocytes in IPF patients are transcriptomically primed to respond to type 1 IFN stimulation and upon arriving in the lungs, differentiate to subsets of macrophages (CXCL10^+^ AM and IGF1^+^ AM) with enhanced type 1 IFN signalling activity.

### Circulating cMono and intmMono in IPF patients show greater magnitude of response to type I IFN stimulation

Given the potential link between enhanced type 1 IFN signalling activity in circulating monocytes and alveolar AMs, we examined if the increased type 1 IFN pathway regulon activity in circulating monocytes is reflected functionally, using protein-based phospho-studies. Cryopreserved PBMCs from IPF patients (n=21) and age-matched controls with healthy lungs (n=11) (Oxford Cohort B) (Figure 4A, Suppl Table 1) were stimulated with recombinant type I IFN (250 U/ml of IFN-β for 15 minutes) and stained pre and post stimulation with a panel of antibodies optimised to simultaneously quantify phosphorylation of multiple proteins in the IFN signalling pathway (Figure 4A, Suppl Table 6) ^39^. This allowed us to map the canonical (pSTAT1) and non-canonical (pSTAT3, pSTAT5-6, pMAPKAPK2, pERK1/2, pp38, pNFKBp65) type 1 IFN signalling pathways^40^. Single cell clustering was performed using *Flowsom* (k=20) (Figure 4B). The monocytes and dendritic cells (DCs) clusters were sub-clustered and annotated using labels from the CyTOF component of the COMBAT study^36^. Four subsets of DCs [plasmacytoid DC, classical cDC 1 (CD141^+^) and cDC2, and a monocyte-derived DC (monoDC)]and 5 subsets of monocytes – CD16^mid^ monocyte, CD64^hi^ monocytes (CD64^hi^ mono), classical monocyte (cMono), non-classical monocytes (ncMono) and intermediate monocytes (intmMono) could be annotated using protein markers. These complemented our single cell RNA-led annotation(Figure 4C) and allowed identification of intmMono (CD14^hi^CD16^hi^ monocytes) which we did not capture on single cell RNA annotation. IntmMono subset and CD64^hi^ mono subset (likely equivalent to mono.type 1 IFN subset) were very small (both less than 0.5% of monocytes -DC population) (Figure 4D). CD16^mid^ mono population is possibly an early monocyte-derived DC population (Figure 4C). Abundance comparisons are shown in Suppl Figure 5.

**Figure 4.**
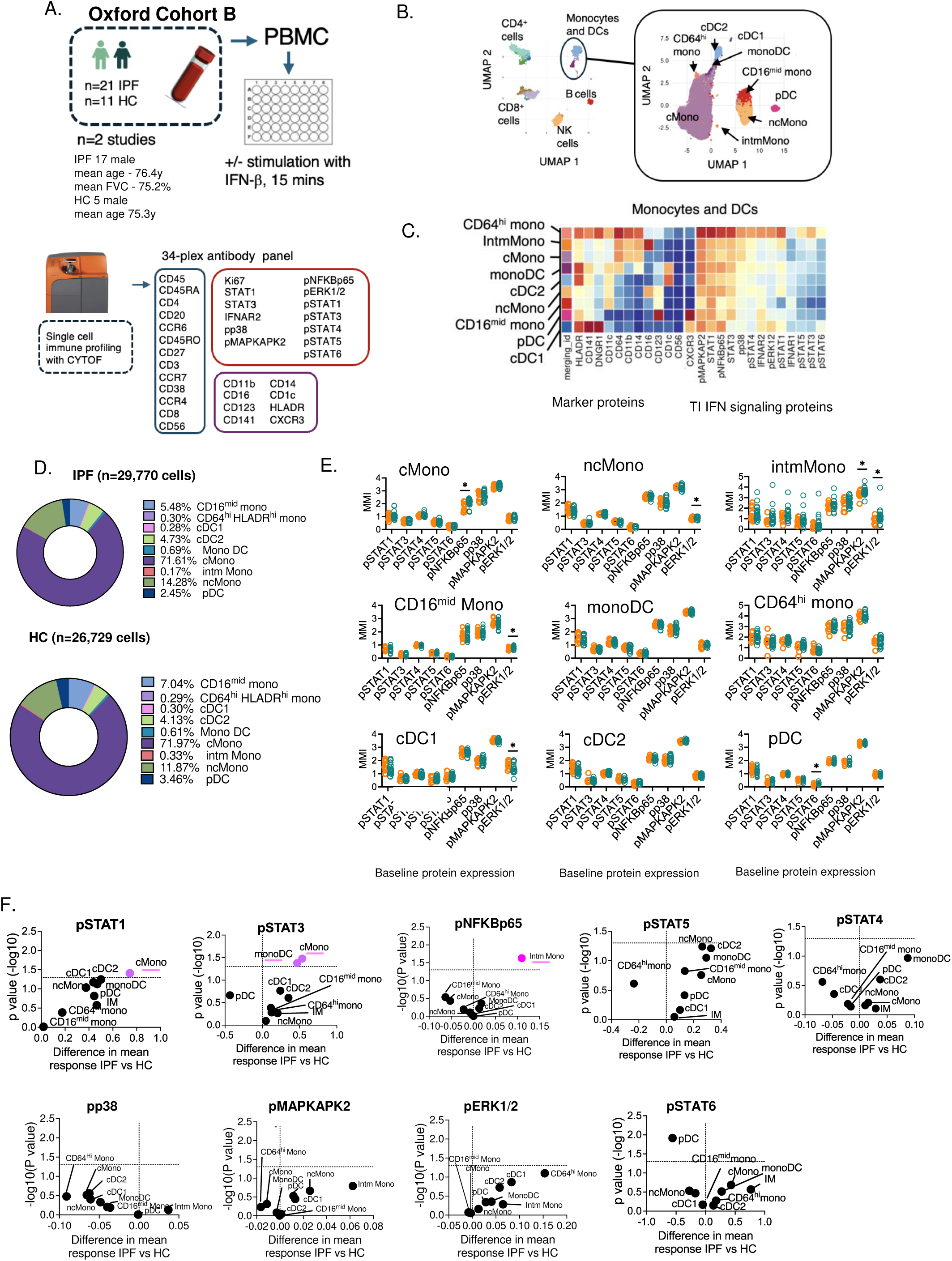
Circulating cMono and intmMono in IPF patients show greater magnitude of response to type I IFN stimulation. A. Oxford Cohort B and overview of study and CyTOF antibody panel B. UMAP of PBMC clustered using Flowsom. Monocytes and DCs are circled and further subclustered. Monocyte and DC subclusters annotated according to expression of proteins and analysed against labels generate by CyTOF analysis in COMBAT for annotation C. Baseline (unstimulated) expression of ‘marker proteins’ used for immune phenotyping and ‘T1 IFN signalling proteins’ D. Composition of monocyte and DC subtypes for IPF and HC (% of all circulating monocytes and DCs) E. Mean metal intensity (MMI) of different phospho-proteins in the type 1 IFN signalling pathway at baseline, comparing IPF to HC monocytes and DC subtypes. * adj p<0.05, Kruskal-Wallis multiple comparison F. Statistical analysis of response to IFN-b stimulation for all monocytes and DC subtypes between IPF and HC; analysed using Student t test; graphed together to allow simultaneous visualisation of all cell subtypes. Purple coloured points are those with statistically significantly higher response in IPF vs HC (p<0.05). Vertical dotted line refers to point where there is no difference in response between IPF and HC; points above horizontal dotted line refers responses that reached statistical significance.

At baseline (without in vitro stimulation), we observed that IPF monocytes already showed increased levels of phosphorylated entities in some of the non-canonical type 1 IFN signalling proteins - pNFKBp65 in cMono, pMAPKAPK2 in intmMono, and pERK1/2 in ncMono, intmMono and CD16^mid^ mono respectively (Figure 4E). But in general, canonical type 1 IFN signalling pathway appears quiescent in keeping with the transcriptomic and regulon findings. However, with IFN-β stimulation, the magnitude of pSTAT1 response was significantly higher in IPF cMono. pSTAT3, a negative regulator of STAT1 activation ^40, 41, 42^, was also significantly higher after IFN-β stimulation in cMono and monocyte-derived DC (monoDC) but not other monocyte or DC subsets (Figure 4F). Notably, there was no difference in response to type I IFN between IPF and HC for pDC, the archetypal producer of viral-induced type 1 IFN, nor for CD64^hi^HLADR^hi^, the monocyte subset with the strongest type 1 IFN signature at baseline (Figure 4F). The only other difference in post stimulation was upregulation of pNFKBp65, a key transcription factor in immune and inflammatory responses, in ncMono; and down regulation of pSTAT6, a key mediator of TH2 signalling, in IPF patients (Figure 4F).

Thus, here, in a high resolution, protein-based functional study of circulating monocytes and DCs, we show that stimulation with type 1 IFN resulted in a significantly higher expression of pSTAT1 and accompanying pSTAT3 in IPF’s classical monocytes. At baseline, several monocyte subsets from IPF patients showed higher levels of phosphorylated non-canonical type 1 IFN signalling molecule, indicating an overall increased baseline inflammatory state.

### Monocytes in blood and lung enriched with type I IFN-associated genes are preferentially found in earlier stages of disease

Finally, we explored if monocytes and/or macrophages with increased type 1 IFN activity have a role in fibrogenesis and progression of disease in IPF. We recruited a third cohort of IPF patients (Oxford Cohort C) (n=28) from the same clinical setting as Cohort A and B (Figure 5A, Suppl Table 1). In IPF, we have a unique opportunity to assess disease stage as all disease show progressive and unremitting deterioration in lung function, regardless of treatment. Therefore, normalised values of lung function (% of predicted for age, height and gender) represent a temporal point in advancing disease. This is unlike other lung disease (e.g. sarcoidosis or asthma) where lung function can recover after treatment. In this cohort, lung function [forced vital capacity (FVC)] was obtained within a month of blood sampling. CD14^hi^ monocytes were obtained from blood using CD14 magnetic microbeads (Macsbeads^TM^) and RNA isolated for quantification of STAT1, eight ISG transcripts and IFN-β ex vivo (without stimulation), to reflect downstream effect of increased or primed type 1 IFN signalling suggested by findings from Figure 4. We found a significant positive correlation between monocytic gene expression level of STAT1, MX1 and IRF7 in monocytes, and lung function (forced vital capacity, FVC) (Figure 5B). All other ISGs also showed a positive correlation trend. This indicates that increased type 1 IFN signalling activities in monocytes is a feature of early disease.

**Figure 5.**
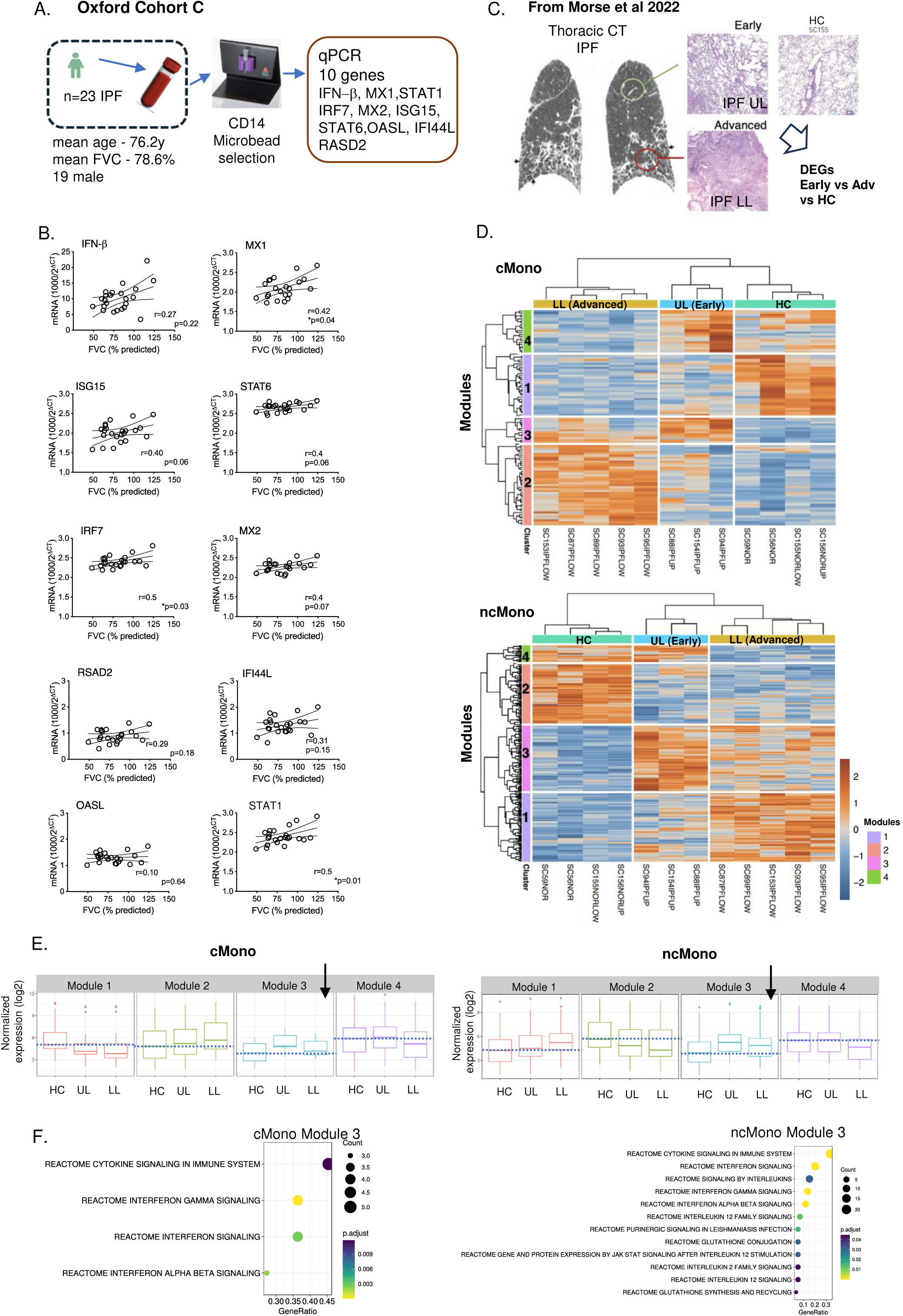
Monocytes in blood and lung with increased type I IFN associated genes are preferentially found in earlier stages of disease. A. Oxford Cohort C and overview of study. B. Correlation between type 1 IFN signalling genes expression in CD14^hi^ monocytes and lung function (FVC%) at point of blood sampling. C. Thoracic CT scan image from one of the patients from Morse et al, showing sites in the lungs where samples were obtained. Image shows typical honeycombing pattern for IPF fibrosis and peripheral or subpleural and basal distribution of disease. Red circle indicates advanced disease, green circle earlier disease. H&E stained lung sections show typical areas of dense fibrosis in lower lobe (LL) and presence of normal alveoli in early disease from the upper lobe (UL). An H&E section from one of Morse’s healthy lungs (HC) is also shown for comparison D. Heat map showing DEGs from three-way comparison between LL, UL and HC lung samples, for cMono and ncMono subsets. Unsupervised hierarchical clustering of these DEGs identified 4 gene modules (modules 1-4). Labelling of columns (e.g. ‘SC153IPFLOW’) was taken directly from Morse’s original labelling of patient and HC samples. E. Boxplot of the median (95% C.I.) normalised gene expression for each gene modules for cMono (left) and ncMono(right) subsets in UL, LL and HC lung samples. Arrows identify the pattern of change [different (in this case higher) in IPF (both UL and LL) compared to HC, and higher in UL compared to LL]. This indicates an IPF-specific enrichment of gene expression module in early disease (UL). Dotted blue line refers to baseline (median expression in HC) F. Pathway enrichment analysis (hypergeometric test; Reactome database) for Module 3 in cMono (left) and ncMono (right)

To complement this blood finding, we asked if monocytes found in the lungs also have a type 1 IFN transcriptomic signature and if these were associated with earlier disease. As negligible numbers of monocytes were found in the alveolar barrier (BAL), we turned to Morse’s lung digest scRNAseq dataset^16^ to answer this specific question. Here, discrete monocyte populations (cMono and ncMono) were present (Figure 1H), possibly representing circulating and/or patrolling monocytes within the lung vasculature. We took advantage of another distinctive feature of the IPF lung disease – the pathognomonic disease pattern, called ‘UIP’ or usual interstitial pneumonia CT and histopathology pattern^43^. One characteristic finding in UIP is the progression of disease from the lower to upper lobes and centripetally from the subpleural sites. In Morse’s study, biopsies had been taken from lung sites with early [upper lobe(‘UL’)] (n=3 samples from 3 patients) and advanced disease [lower lobe (‘LL’)] (n=5 lung samples from 3 patients)and healthy lungs (‘HC’) (n= 4 samples from 3 HC individuals) (Figure 5C). This allowed us to examine if lung monocytes and/or macrophages in earlier stages of disease were enriched for type 1 IFN signalling compared to later disease.

Transcriptomes of all the subsets of myeloid cells in Morse’s dataset - monocytes (cMono, ncMono) and macrophages (SPP1^+^ mac, HSP mac, FABP4^hi^ AM, CXCL10^+^ AM, Intm mac, TM and IM)(Figure 1H) were compared three-ways - between those from the upper lobe, lower lobe and HC lungs. Our focus is the monocytes but the macrophage comparisons were also performed, mainly to determine if there were also enrichment of type 1 IFN signalling pathways in IPF lung digests. Statistically significant transcriptomic differences (likelihood ratio testing; adj p <0.05; fold change >1.5) between UL, LL and HC in these cell types were retrieved, subjected to unsupervised hierarchical clustering and labelled Modules 1 to 4 (Figure 5D). Pathway enrichment analysis (PEA) (using GO Reactome database) were then carried out on all these modules.

From the entire monocyte-macrophage 3-way analysis (UL vs LL vs HC), only three cell types demonstrated gene modules which were significantly differentially expressed between disease and HC, *and* the difference to HC is greater in UL compared to LL. These were cMono (module 3), ncMono (module 3), where module 3 is upregulated in IPF compared to HC, and also in UL compared to LL (Figure 5D-E) and to a much smaller extent, modules 3 and 4 in SPP1^mid-hi^ macs (Suppl Figure 6A). PEA of the modules in cMono and ncMono showed enrichment of IFN signalling (Figure 5F). These modules also displayed enrichment of pro-inflammatory gene sets-multiple ‘cytokine signalling’ gene sets in cMono and ncMono, and IL-12 in ncMono (Figure 5F). Notably, no other gene modules which were statistically increased in IPF (UL and LL) were enriched with genes associated with type 1 IFN signalling.

This unbiased and comprehensive analysis of the lung tissue show that type 1 IFN signalling pathways are activated and enriched in lung monocytes in IPF, and associated with earlier disease. This analysis of Morse’s lung digest macrophage subsets also showed that that there was no enrichment of type 1 IFN signalling in any macrophage subsets in IPF lung digest (in contrast to alveolar space) indicating that upregulation of type 1-IFN signalling in IPF macrophages is confined to the alveolar barrier.

We tested these transcriptomic findings qualitatively with our final cohort of patients (Oxford Cohort D)(Figure 6A, Suppl table 1). We identified n=4 patients who underwent video-assisted lung biopsy to obtain good quality tissue for confirmation of clinical suspicion of IPF and cut sequential lung sections for histopathology analysis [haematoxylin and eosin (H&E) with two independent lung histopathologists and a senior pulmonologist] and immunofluorescence staining (two separate panels of 4-colour staining; Figure 6A). Six good quality, large (approximately 4×5mm) lung sections were obtained from four patients where infection was excluded clinically, and where the histopathologists have identified clear picture of overall differentiation between advanced and early fibrosis on H&E (Figure 6B-C; Suppl Fig 7). In the advanced fibrotic lung sections, the fibrotic areas were dominated by presence of collagen and elastin, smooth muscle hyperplasia, some bronchiolisation of alveolar epithelia (Figure 6B, Suppl Fig 7A-C). In early fibrosis, there were striking presence of sheets of (myo)fibroblasts, fibroblastic foci, type II alveolar metaplasia and greater number of inflammatory cell (Figure 6C, Suppl Fig 7D-F). Temporal and spatial heterogeneity was observed as per clinical definition for UIP histology pattern^43^. Once these sections were selected on those criteria, we performed pSTAT1 phospho-staining concomitantly with CD14 (monocyte) and CD68 (macrophage) staining.

**Figure 6.**
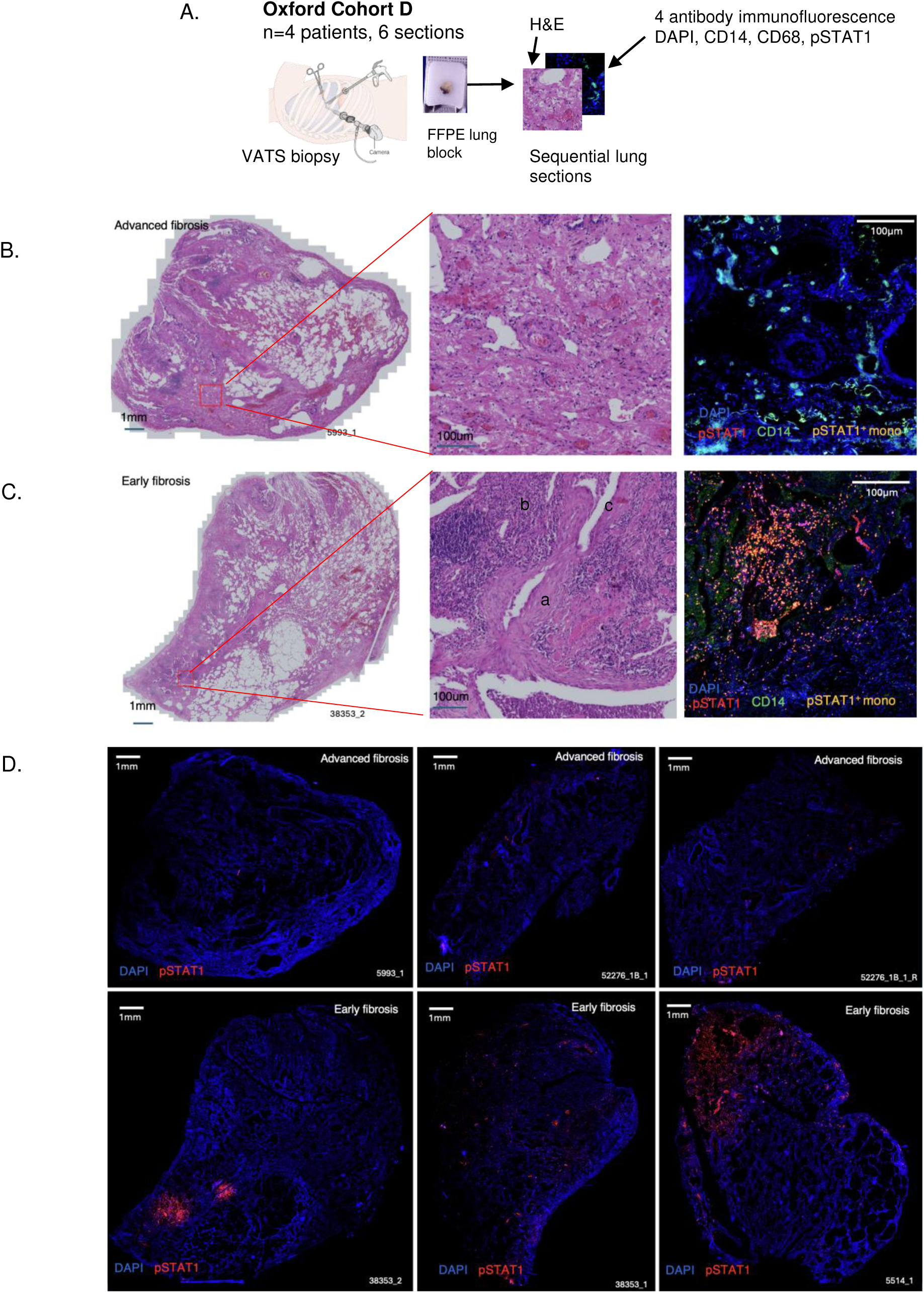
Immunofluorescence study show pSTAT1-expressing monocytes in IPF lung tissue with early fibrotic disease. A. Oxford Cohort D and overview of study. B. H&E sections at low and high magnification of lung section showing advanced fibrosis. Higher magnification section shows presence of elastin and collagen in interstitium of lung. Immunofluorescence section shows no staining with pSTAT1 or CD14 (monocytes). C. H&E sections at low and high magnification of lung section showing advanced fibrosis. Higher magnification H&E section from a representative region of inset (red box) in fibrotic area of lung shows established, advanced fibrosis with dominant presence of elastin and collagen in interstitium of lung. Immunofluorescence section shows no staining with pSTAT1 or CD14 (monocytes). D. H&E sections at low and high magnification of lung section showing early stages of active fibrosis. Higher magnification H&E section shows presence of fibroblastic foci (’a’), inflammatory cells (’b’) and alveolar metaplasia (’c’). elastin and collagen in interstitium of lung. Immunofluorescence section shows pSTAT1^+^ CD14^+^ cells (monocytes). E. Overview of all 6 lung sections at low magnification showing pSTAT1^+^ cells in early but not advanced fibrotic lung sections.

We found a striking presence of pSTAT1 staining in lung sections with early disease (Figure 6D), and specifically in areas with clear fibroblastic and type II alveolar metaplastic activity. These pSTAT1 were expressed predominantly by CD14-expressing cells (Figure 6C). Although qualitative, these findings from blinded studies confirm the presence of pSTAT1-expressing monocytes in areas of early disease activity in IPF.

Overall, these findings show that monocytes in the blood from IPF patients displayed evidence of type 1 IFN signalling activity that was most prominent in the early part of the clinical disease. In the lung tissue, a similar finding was found in two scenario – by single cell transcriptomic profile of lung digest from sites of early vs later disease, and by pSTAT1 protein expression and monocyte analysis of histopathologically polar ends of disease stages. These findings further support a role for type 1 IFN-active monocytes in initiation of fibrosis in IPF.

## Discussion

Current evidence suggests that repeated micro-injury to a vulnerable, senescent alveolar epithelium and cumulative genetic and oxidative stress, is a key trigger for the disease. However, the immune mechanisms underlying this impaired wound-healing response remain poorly understood. While myeloid cells are likely to be involved, their specific role is still unclear. Here, we provide a comprehensive, cross-compartment (blood, lung tissue and alveolar space) and unbiased, transcriptome-wide interrogation of myeloid cells in IPF lung tissue and alveolar space. We found monocytes and macrophages with type 1 IFN signalling and enhanced inflammatory pathways as the key differentiating factor between IPF and HC macrophages at the alveolar barrier (but not in lung tissue). Classical monocytes in the blood are primed to respond to type 1 IFN and in the lung, showed evidence of type 1 IFN pathway activation. These features are more pronounced in early stages of disease, supporting a role for type 1 IFN-activated monocytes in initiation of fibrosis in IPF while barrier-related type 1 IFN-activated macrophages in drive chronic fibrosis^44^.

Type 1 IFN signalling is a double-edged sword, while critical for defence against pathogens^45^ it has also been shown to epithelial injury in various contexts ^46^. CXCL-10, an interferon-stimulated gene (ISG) and also the highest differentially expressed genes in mono-SPP1+ and IGF1^+^ AMs, is a pro-inflammatory cytokine which induces chemotaxis, differentiation and activation of NK cells, monocyte, and T cells ^47^. CXCL10-positive macrophage have been reported in several studies. In Mulder’s monocyte-macrophages single cell compendium dataset, CXCL10^+^ macrophages were detected across different types of disease, including hepatocellular carcinoma, Sars-Cov-2 infected lung, osteoarthritic and rheumatoid arthritis^24^. While the cause of type I IFN activation in these macrophages and circulating classical monocytes is unclear, it is probably connected. Potential reasons are increased latent viral burden^48^, a sequelae of auto inflammation and, we think most plausible, re-circulating monocytes from the lungs. In the lung, interactions between monocytes and damage-associated molecular patterns (DAMPs) from injured alveolar epithelium likely contribute to type I IFN activation. Notably, elevated levels of mitochondrial DNA—a DAMP and known inducer of type I IFN signalling—have been observed in the serum and BAL fluid of IPF patients^49^. Alveolar epithelial cells and fibroblasts in IPF also exhibit signs of cellular injury, senescence, mitochondrial dysfunction, and accelerated aging ^49, 50^, and may thus serve as sources of DAMPs.

The increased expression of ISGs in circulating monocytes may be explained by the higher activity of type I IFN-stimulated monocytes in early-stage IPF. Consequently, the lungs may serve as a source of circulating type I IFN-primed monocytes. An alternative hypothesis is that type I IFN-activated macrophages in IPF originate from pre-primed monocytes in the blood, although this seems less likely given the observed reduction in ISG expression in blood monocytes during more advanced stages of the disease. Either way, these findings suggest that targeting monocyte activation, trafficking, and differentiation—especially in early disease—could influence disease progression. A pro-inflammatory, type I IFN-primed monocyte population may also contribute to the severe inflammatory response observed during acute exacerbations of IPF (AE-IPF), which carry a high mortality rate (∼80%). ^51^. Here infection could unleash overwhelming monocyte and macrophage induced injury on the alveolar epithelium and trigger accelerated fibrotic response to injury in a predisposed host. In this context, infection may unleash a hyperinflammatory response from monocytes and macrophages, leading to severe alveolar injury and accelerated fibrosis observed in these patients. Clinically, prioritizing infection prevention and prompt antimicrobial treatment could reduce mortality in AE-IPF.

Notwithstanding the importance of Type 1 IFN-activated monocytes and macrophages, we also observed a very high level of FN1 expression in CD206^hi^ FN1^hi^ AMs and CXCL10^+^ AMs in IPF. FN1 encodes fibronectin, an extracellular matrix that promotes fibro to myofibroblast differentiation^52^. Monocyte-derived macrophages appear also to over-express fibronectin after IL-4 stimulation^53^. Of relevance to immune-epithelial regeneration, Hewitt reported that interaction between fibronectin and alveolar epithelial cells in IPF inhibited epithelial cell migration, required for regeneration of an injured epithelium^54^. We recently showed in single cell imaging and spatial analysis of macrophages and regenerating alveoli that CD206^hi^FN1^hi^ AM are the main cells that are co-located with aberrant regenerating alveolar epithelium^27^. Independent to this paper, analysis of deposited data from Haberman^55^ in that study^27^ also isolated FN1 pathway as the key receptor-ligand pathway in this interaction. Together, these findings suggest a potentially pathogenic role for IPF CD206^hi^FN1^hi^ AM at the alveolar barrier in IPF.

An open question remains as to whether the macrophage clusters identified represent distinct cell types or dynamic states of the same population. While neither transcriptomic profiling nor trajectory analysis offers a definitive answer, the similarity in gene expression and functional profiles between FABP4^hi^ AMs and yolk-sac-derived macrophages suggests that FABP4^hi^ AMs are likely self-renewing, homeostatic resident macrophages. In contrast, CXCL10^+^ AMs may represent inflammatory states of monocyte-derived macrophages. IGF1^+^ AMs share transcriptional similarities with FABP4^hi^ AMs and may represent an inflammatory state of resident alveolar macrophages. CD206^hi^FN1^hi^ AMs appear distinct from other clusters, though their origin remains unresolved.

The lack of DEGs in circulating monocytes (Oxford Cohort A) (Figure 3F) in contrast to lung tissue monocytes (Morse cohort) (Figure 5D) suggest a the profound influence of the tissue niche on monocytes. While blood monocytes only show a modest upregulation of type I IFN-related pathways, lung-based monocytes display an amplified IFN response (Module 3; Figure 5D). This is the case for classical monocytes but interestingly also for non-classical monocytes which patrols the vascular lumen^56^. In non-classical monocytes, additional gene pathways are upregulated in module 3 (Figure 5F), in particular, IL-12 signalling pathway. Enrichment of IL-12 gene sets in ncMono and its association with early disease is noteworthy as IL-12 bridges innate and adaptive immunity by promoting Th1 differentiation and increasing IFN-γ and TNF-α production in T and NK cells^57^.

This study should be considered within certain limitations. The number of individual patients used for the scRNAseq study is small although the number of single cells is high, the cohort is representative of the disease and this is the only study to date profiling the alveolar space in IPF. This is also the only study with paired BAL and blood samples allowing for direct comparison of monocytes in blood and lungs. Furthermore, we tested our hypothesis using multiple approaches involving a total of 54 independent IPF patients. Secondly, the work is primarily descriptive although this was complemented by stimulation studies for the key identified pathway. We refrained from inferring cell-cell cross talk as we have previously shown that such analyses can be misleading without spatial confirmation of cellular proximity ^27^.

In conclusion, our high-resolution, single-cell transcriptomic analysis reveals a spectrum of pro-inflammatory alveolar macrophage states in IPF, with augmented type I IFN signalling activity in IPF, particularly during early disease. We also provide the first detailed description of alveolar macrophage heterogeneity in both the alveolar barrier and lung digest of IPF lungs.

## Materials and Methods

### Study subjects, sample collection and handling

For Oxford Cohort A (and B) IPF patients were recruited from Oxford Interstitial Lung Disease Clinical Service, Oxford University Hospitals. A diagnosis of IPF was made following review of clinical and radiological features at a multi-disciplinary team meeting and using criteria defined by the 2018 joint ATS/ERS/JRS/ALAT statement^43^. Patients did not have coexisting pulmonary disease, immune-mediated diseases, cancer or infection.

For Oxford Cohort A, HC subjects were those undergoing a clinically relevant bronchoscopy for discrete episodes of unexplained and resolved haemoptysis. Further follow up for up to a year showed no clinical evidence of abnormalities or respiratory pathologies. In both IPF and HCs, BAL were performed by a senior pulmonologist using the Olympus Key bronchoscope (Olympus KeyMed, Essex, UK), with methods previously described^58^. All samples were processed within 30 minutes. BAL fluid was passed through a 100um filter, centrifuged at 400g for 10min, cells resuspended in RBC Lysis Solution (QIAGEN, Manchester UK) for 10min, washed, counted and used. PBMCs were separated via density gradient separation using Lymphoprep™ (Axis-Shield, Dundee, UK).

For Oxford Cohort B – PBMCs from IPF and HCs were derived as described above and stored in FCS/DMSO using standard procedures previously described^37^. HC’s PBMC samples were collected from patients attending routine pre-operative eye clinic appointments prior to cataract surgery.

For Oxford Cohort C, monocytes were isolated from PBMCs by positive selection using anti CD14 microbeads (Miltenyi Biotec) according to the manufacturer’s instructions. Post separation, CD14^+^ monocyte purity was assessed by flow cytometry (CD3, CD19, CD15, CD16 and CD14). Only samples with purity of greater than 98% were used.

Oxford Cohort D utilised archived diagnostic formalin-fixed paraffin-embedded (FFPE) blocks for immunofluorescence from Oxford University Hospitals NHS Foundation Trust via the Oxford Radcliffe Biobank (REF 19/SC/0173).

For all cohorts, HC were defined as those without lung disease, active cancer (cancer still on treatment or those within 10 years of diagnosis) and symptoms or signs of infection or on respiratory system examination. On recruitment all patients and subjects were screened to include only never-smokers or those abstinent for at least 1 year. 1 IPF and 1 HC in Cohort B, and 1 IPF in Cohort C were found to be current smoker on final checks and documented in Suppl Table 1).

The studies received ethical approval from the Health Research Authority and South-Central National Research Ethics Service (14/SC/1060, 18/SC/0227, 21/WM/0244 and 19/SC/0173).

### Droplet-based scRNA sequencing

Droplet-based scRNA-seq was undertaken using the 10x Chromium Single Cell platform in accordance with the manufacturer’s instructions (10x Genomics, 3′ v3 chemistry). BAL cells and PBMCs were further filtered through a 70um filter and stained with TotalSeq™-A anti-human Hashtag antibodies numbers 1-8 (BioLegendTM London, UK) according to manufacturer’s instructions. Approximately 30,000–40,000 cells were loaded per pool. The workflow involved encapsulating individual cells within gel beads in emulsion (GEMs), barcoding, reverse transcription of complementary DNA (cDNA) within GEMs, cDNA clean-up, amplification, and library construction for gene expression (GEX) and antibody-derived tags (ADT) data.

Library quality and integrity were assessed using Agilent Bioanalyzer TapeStation to ensure compliance with sequencing requirements. Final libraries were pooled at a 4 nM concentration and sequenced on an Illumina NextSeq 500/550 (High Output v2.5, 150 cycles) or outsourced to a NovaSeq PE150 using 150 base-pair paired-end reads. Sequencing depth and run parameters followed the specifications provided by the manufacturer (CG000208, Rev F).

### CyTOF antibody preparation methods

Metal tagged antibodies were purchased directly from Standard BioTools or conjugated in-house (Suppl Table 6). For Lanthanide isotope conjugations the Maxpar® X8 Antibody Labelling Kit (Cat. 201300, Standard BioTools) was used and for Cadmium conjugations the Maxpar® MCP9 Antibody Labelling Kit (Cat. 201114A, Standard BioTools) was used; both according to relevant protocols in Maxpar Antibody Labelling User Guide ver. 14. CD8 was conjugated with 198Pt directly without initial polymer loading by conjugating partially reduced antibody with 198-Cisplatin at 250uM for 90 mins at 37°C. To confirm antibody conjugation with metal isotopes, one drop of Ultracomp™ ebeads (reactive to mouse, rat and hamster origin antibodies, Cat. 01-2222-42, Invitrogen) was incubated with 1uL of conjugated antibody and counterstained with CisPt before acquisition on the 3^rd^ generation mass cytometer (Helios, Standard BioTools) using CyTOF Software v7.0.

### IFN-β stimulation, metal-tagged antibody staining and acquisition

Cryopreserved PBMCs in Oxford Cohort B were thawed and initially stained with a selected number of antibodies which were sensitive to fixation prior to stimulation with IFN-β as previously described^39^. PBMCs for each sample were split into 2 equal portions and either treated with IFN-β 250 unit/ml or culture media (RPMI/10% FCS) for 15 mins at 37°C.

Each sample was barcoded using Standard Biotools 20-plex barcoding kit according to manufacturers instructions. Barcode samples were pooled and stained sequentially with cell surface and intracellular antibody staining mixture, fixed and incubated with 125 mM Cell-ID Intercalator at 4°C overnight prior to acquisition. On the day of acquisition, cells were washed filtered with a cell strainer and mixed with a 1 in 10 dilution EQ beads. Pooled samples were acquired at a rate of < 300 events/second on a Helios Mass Cytometer (Standard Biotools).

### qPCR

CD14 positively-selected monocytes were lysed in RLT buffer (QIAGEN) supplemented with beta-mercaptoethanol (Sigma-Aldrich) before RNA extraction using the RNeasy Mini Kit (QIAGEN) and frozen at −80°C. Batch RNA extraction was performed using the RNAeasy Mini Kit (QIAGEN) according to manufacturer’s protocol. RNA integrity was assessed using nanodrop and Agilent technology. cDNA was synthesized using High Capacity cDNA RT Kit (Applied Biosystems). qPCR was performed using the 2X Fast SYBR® Green Master Mix (Applied Biosystems) in a 7500 Fast Real-Time PCR system (Applied Biosystems). Primer sets are shown in Suppl Table 7

### Immunofluorescence staining of formalin fixed paraffin embedded tissue sections

Sequential 0.5 μm sections were cut from the FFPE tissue blocks for H&E (Sigma) and immunofluorescence staining. Prior to antibody staining slides were incubated on a heat block set at 60 degrees for 10 mins followed by incubation in Histoclear II (National diagnostics) to dewax. Sections were rehydrated using Ethanol (100% for 10 mins repeated three times) and permeabilized with Triton X 0.1%. Heat mediated epitope retrieval was performed at 96 degrees for 30 minutes using Tris-EDTA buffer (pH9). After washing, sections were incubated with blocking buffer containing 2% BSA in tris buffered saline containing 0.1% tween 20® (TBS-T) for 30 minutes at room temperature followed by incubation of primary antibodies (Suppl Table 8 targets, clones, dilution) at 4 degrees overnight. Incubation with fluorophore conjugated secondary antibodies was performed at room temperature, followed by counterstaining with DAPI. Paired ‘no primary’ or isotype control slides were used to analyse signal attributable to background or non-specific staining. Tissue sections were imaged using a Axio Scan Z1 fluroscence microscope (Zeiss). Using Zeiss software, the microscope was configured for AF405, AF488, AF568, and AF647, and the entire area of the tissue section was selected using the software for higher resolution scanning, utilizing a plan apochromat lens (40x 0.95 korr M27 objective). Images were saved on a computer for further processing using custom Fiji/Image J macros.

All histopathology analyses were performed with a senior Respiratory specialist pathologist (CC) and interstitial lung disease pulmonologist (LPH).

### Single Cell RNA sequencing data analysis

#### Alignment, hashtag demultiplexing and quality control

Raw sequence read quality was assessed using FastQC software (version 0.11.9)^59^. There were two sequencing pools for Oxford Cohort A samples. For each sequenced scRNA-Seq pool, Cellranger pipeline (version 3.1.0) from 10X Genomics, (https://support.10xgenomics.com/single-cell-gene-expression/software/downloads/latest) was used to process raw data, align reads (against hg38-v3.0.0 human reference genome from the UCSC ftp^60^. For corresponding antibody libraries, CITE-Seq Count (v 1.4.3) were used to process and obtain hashtag (HTO) antibody tag UMI for each cell barcodes. Read UMI counts were summarised using 16-base barcodes for TotalSeq antibody libraries. Antibody UMI count matrices were then further filtered against the 10X cellular barcode whitelist for the corresponding 10x version 3.0 chemistry.

Empty droplets were distinguished and removed based on mRNA count distributions. mRNA with misassigned barcode, possibly from the sequencing index swapping were removed using DropletUtils R package^61^. Empty droplets containing only ambient RNA were then removed using emptyDrops function in DropletUtils^61^. Barcodes with < 5% FDR were retained.

For the remaining cells, HTO count matrix was added to the data, normalised by CLR (Centered Log-Ratio) method and demultiplexed based on their sample of origin using R package Seurat’s HTOdemux function. In brief, normalized count for each hashed ID were fitted with a negative binomial distribution. Positive threshold was set to 99^th^ percentile of the recovered normalized unique molecular identifier (UMI) counts for the hashtag where cells below this threshold was considered negative for the tag. Cells that were negative for hashtags and positive for multiple hashtags were filtered out. For the remining demultiplexed cells, we further checked whether cells were segregated correctly based on sample-of-origin, by examining segregated expression of sex-specific gene (e.g. XIST) within tSNE embedding. After filtering out these cells, we further removed the cells with low total RNA content and those that contained a high percentage of total UMIs originating from mitochondrial RNAs. An initial clustering was performed and those clusters with low quality cells as defined above were also excluded. Thresholds were derived for each of the two sequencing pools. After QC, 26,762 cells (2349 detected genes 10770 UMI counts per cell on average) were obtained for the downstream analysis. 914 cells were removed.

### Cell type clustering and cell type identification scRNA seq

Seurat R package^62^ (version 3.3.0) was used for data normalisation, scaling, transformation, clustering, dimensionality reduction and most of the data visualisation. For each individual pools, UMI count matrix was log-normalised using function NormalizeData and ScaleData. Highly variable genes were identified by fitting the mean variance relationship. Dimension reduction was performed with principal component analysis (PCA). Scree plots were then used to determine number of principal components to use for clustering analyses for each pool. After the k-nearest neighbor (kNN) graph was constructed, cells were clustered using Louvain algorithm for modularity optimization.

For Oxford Cohort A, cells from two sequencing pools were merged into a single dataset and batch effect was corrected using Harmony (version 0.1.1)^63^. Merged cell clustering and visualization of cells from all pools was performed as before, using Louvain and UMAP algorithms with 20 harmony dimensionality reduction components as input (instead of principal components). The resolution parameter for clustering was set to 0.7. Optimal clustering resolution is settled by a combination of biology and assessment of cluster stability with R package Clustree (Version 0.5.1). Cell cluster annotation was done as described in results using a combination of known marker genes and cross classification with previously published reference atlas using Seurat label-transfer workflow.

### scRNA-Seq analysis of public data (Morse lung digest dataset)

Publicly available scRNA-Seq data (Morse et al)^16^ were first filtered for empty droplets using emptyDrops function in DropletUtils (same parameters as described above), poor quality cells with less than 300 detected genes and greater than 30% mitochondrial reads per library were removed. 48,728 cells were further analysed and clustered in Seurat using same procedures applied on our own data. 21,731 cells were assigned as ‘myeloid cells’ based on expression of cluster defining markers, and then further subclustered with Louvain clustering (clustering resolution =0.8).

### Differential expression and pathway analysis

To identify differential expressed genes between IPF and HC, DEG analysis was performed using pseudobulk approach. After generating pseudo-bulk aggregate of every cells of the same sample for each cell subsets, differential gene expression (DEG) between disease groups were computed using DESeq2 pipeline^64^. FDR corrected p-value under 0.1 were considered as significant and used for further functional enrichment analysis. Gene Ontology (GO) and Reactome pathway enrichment analyses were performed using the enrichGO function of the clusterProfiler R package (ver 4.8.1)^65^.

To identify differential expressed genes between 3 different disease state (HC, IPF-LL, IPF-UL; Figure 5) (Morses’ publicly available dataset), likelihood ratio test (LRT) was performed using the DESeq2 pipeline where full model included an intercept term and disease information while reduced model included an intercept term only. DEGs from LRT test with *Benjamini*–*Hochberg* (BH) adjusted p value < 0.05 were considered as significant. Expression pattern of time course dependent DEGs were visualized as heatmap via heatmap package (version 1.0.12) using vst normalized pseudo-counts data as input. Clusters of genes with similar expression patterns across time point groups were defined using the cuttree function.

### Transcription factor analysis

To infer regulon activity in each cell, SCENIC (single-cell regulatory network inference and clustering) workflow was used (pyscenic)^28^. The normalised single cell gene expression matrix was first filtered to exclude genes that were expressed in less than 10 cells. The RcisTarget database, containing transcription-factor motif enrichment scores for gene promoters around their transcription start site for the hg38 human reference genome was used as reference (https://resources.aertslab.org/cistarget/databases/homo_sapiens/hg38/refseq_r80/mc9nr/gene_based/). Using the genes available in RcisTarget database, gene-gene co-expression relationships between transcription factors (TF) and their potential targets were inferred by using GRNBoots2 algorithm^66^. The R package SCENIC was used to perform transcription factor network analysis to detect co-expression modules enriched for target genes of each candidate TF. The regulon or TF ‘activity score’ for each TF module for each cell was then calculated using the AUCell package, where gene-expression ranking was computed using ‘AUCell-buildRankings’ function followed by calculation of area under the curve (AUC) score based on the fraction of TF target genes within the top ranking genes for each cell. Wilcoxon test was used to compare TF activity score between IPF and HC.

### JASPAR motifs analysis

We use this method to assess association between motif presence and gene transcriptional changes in IPF. Clustered transcription factor (TF) motifs were sourced from the JASPAR 2024 Vertebrata CORE database^33^. Clusters 25 and 41 were labeled as GAS-like and ISRE-like, respectively. Position weight matrices (PWMs) were derived from motif position count matrices (PCMs) using a uniform background and pseudocounts based on the logarithm of total position counts.

Constructed PWMs were used to screen promoters of Ensembl Canonical and MANE Select protein-coding transcripts in the GRCh38 genome (Ensembl v112)^67^. Promoters were defined as ENCODE Promoter-like sequences (PLS)^68^ within 200 bp of the transcription start site (TSS). If no PLS was available, the promoter region was defined as ±175 bp around the TSS. For each motif, the maximum PWM response was calculated across the promoters of each gene and Z-normalized across genes. Responses for motifs within the same cluster were summed and Z-normalized again to produce final gene-specific ‘motif cluster score’.

Next, we assessed the association between motif presence and gene transcriptional changes in IPF. For this, all protein-coding genes for each AM subset were first filtered to retain only genes with fold change in expression (comparing IPF vs HC) that exceeded the corresponding standard error (SE). Filtered genes were then partitioned into a ‘motif cluster-positive’ group (where the motif cluster Z-score is ≥ 2, indicating that the gene is putatively regulated by the motif cluster’s TF) or ‘motif cluster-negative’ group (Z-score < 2, i.e. baseline) for each JASPAR motif cluster. The average fold change of the gene (IPF v HC) is then compared between the motif cluster-positive and motif cluster-negative groups using the Wilcoxon signed-rank test. This identifies motif clusters that are statistically significantly associated with increased gene expression in IPF(Figure 2D). This analysis was performed for all motif clusters across all cell populations, with p-values adjusted for multiple comparisons using the Benjamini–Hochberg correction.

### Trajectory analysis

For Oxford Cohort A, we applied slingshot (v.2.12.0)^38^ to infer the lineage differentiation of macrophage subtypes with potential developmental relationship. After extracting 12,922 BAL macrophage and 2,395 blood monocytes from the global data, these myeloid populations were then normalised, batch corrected and clustered using Seurat and Harmony as mentioned above using Louvain clustering resolution 0.7. Slingshot was run on this UMAP space with batch correction. The identified paths were mapped to UMAP projection for visualization.

### Code and source data availability

The source code used in this manuscript are available in the Github repository at https://github.com/jeongmin-woo/IPF_BAL_paper. The raw .fcs files used for the CYTOF analysis and all Seurat Objects used in the single cell RNAseq analysis are available at 10.5281/zenodo.15220246

### CyTOF analysis

Normalisation within batches of cytometry data was performed using ‘FCS Processing’ function in CyTOF (Standard Biotools ‘CyTOF’ package v7.0, Passport ID EQ-P13H2302_ver2) followed by concatenation. Concatenated files were de-barcoded using CyTOF ‘Debarcoder’ function with standard parameters from the Plex 20-ID debarcoder key. Individual .fcs files were then manually gated according to gaussian parameters followed by EQ bead, doublet and dead cell exclusion [FlowJo(version 10.8.1)].

Batch correction was performed using cyCombine package^69^ of arcsinh transformed marker expression data (cofactor = 5). The flowset was converted into an SCE object using the df2SCE() function from cyCombine.

Analysis of the pre-processed .fcs files containing live single cells was performed using the ‘Catalyst’ workflow (version 1.24.0)^70^. Unsupervised clustering was performed using the FlowSOM self-organising map algorithm using phenotyping (‘type’) markers at an initial resolution of 100 (dim x= 10, dim y = 10) to give 100 initial clusters followed by consensus clustering and merging of metaclusters to a maximum cluster resolution (k=20). Manual cluster annotation was performed to identify discrete populations based on arc sinh transformed marker expression. To further identify discrete monocyte populations including intermediate monocytes not resolved with initial clustering select populations (c10 – “CD14^lo^ Mono”, c12-“cDC2”, c13-“cDC1”, c14-“pDCs”, c16-“CD14^hi^ Mono”) were re-clustered using myeloid related phenotyping markers, (HLADR, CD141, DNGR1, CD11c, CD64, CD11b, CD14, CD16, CD123, CD1c, CD56). Re-clustering of the monocyte and DC populations was performed at a resolution of (81 dim x = 9, dim y = 9) following by merging of meta clusters to a maximum cluster resolution of 9.

Raw count data was used to explore differences in proportions of populations between IPF and control samples. To examine differences in response to IFNβ, arcsinh median metal intensities (MMI) values for unphosphorylated and phosphorylated proteins involved in canonical and non-canonical signalling downstream of the IFNAR were extracted. Antibodies targeting pSTAT2 were not commercially available at the time of panel design.

## Data Availability

All data have been deposited online and links will be live upon publication of manuscript

## Funding

The study is funded by combination of grants from NIHR Oxford Biomedical Research Centre grant. LPH is supported by MRC (grant CFR01480) and the NIHR Oxford Biomedical Research Centre (grant NIHR 203311). We thank Dr Rachel Rigby for previous work done on the Type 1 IFN CYTOF panel. The views expressed are those of the author(s) and not necessarily those of the NIHR.

## Author contribution

VN analysed data and results, collected and processed some samples and wrote the paper with LPH. PW and LD analysed data and results and discussed concepts and interpretations. LD performed the scRNAseq for Oxford Cohort A with PW and LPH. TH analysed and contributed to analysis of some data. Agne A and JMW performed the bioinformatic analysis with biological contribution from PW, VN, AS, LD and LPH. HC and MM ran the CyTOF studies (Oxford Cohort B) with PW, VN and HP. SZ and Andrew A collected some blood samples and clinical data and contributed to discussions. CC performed all the expert histopathology analyses in discussions with LPH. AF ran the motif analysis. JR developed the CyTOF phospho-study panels and experiment protocols. CV performed the immunofluorescence staining with VN, analysed and processed the images; and the qPCR studies (Oxford Cohort C) with LD. LPH conceived and led the study and the conceptual and intellectual development of the work with contribution from PW, LD, VN, Agne A, and JR for different parts of the work. All authors read the paper and were involved in corrections and refining the final paper.

## Notes

### Competing Interest Statement

The authors have declared no competing interest.

### Funding Statement

The study was funded by MRC and NIHR

