## Supplementary material for "Type I IFN-activated lung monocytes and macrophages as initiators and drivers of fibrosis at the alveolar barrier in IPF": Suppl Table 2

**Suppl Table 2. Intercluster DEGs for AMs from BAL (IPF & HC); Oxford Cohort A.** Statistically significant (p adj <0.01- Bonferroni correction) cluster defining genes for Oxford Cohort A BAL AM subset ranked by Log2 fold change.  Pct.1 – proportion of cells in selected cluster expressing gene. Pct .2 proportion of cells expressing gene in all other remaining clusters.

| Gene | avg_log2FC | pct.1 | pct.2 | p_val_adj | cluster |
|---|---|---|---|---|---|
| FABP4 | 1.75282531 | 0.998 | 0.918 | 7.07E-27 | FABP4hi AM |
| APOC1 | 1.30051837 | 1 | 0.988 | 1.74E-28 | FABP4hi AM |
| RBP4 | 1.29731316 | 0.763 | 0.431 | 7.67E-13 | FABP4hi AM |
| SCD | 1.23744855 | 0.947 | 0.739 | 5.22E-20 | FABP4hi AM |
| CAMP | 0.99614674 | 0.526 | 0.134 | 7.61E-13 | FABP4hi AM |
| CCL18 | 0.99582505 | 0.968 | 0.881 | 7.20E-07 | FABP4hi AM |
| NUPR1 | 0.93504013 | 0.956 | 0.742 | 1.38E-11 | FABP4hi AM |
| ABCG1 | 0.83184976 | 0.955 | 0.756 | 9.67E-18 | FABP4hi AM |
| PLIN2 | 0.82387701 | 0.978 | 0.872 | 1.69E-10 | FABP4hi AM |
| GCHFR | 0.81621992 | 0.997 | 0.942 | 1.37E-14 | FABP4hi AM |
| CES1 | 0.78256153 | 0.821 | 0.565 | 7.22E-10 | FABP4hi AM |
| APOE | 0.73236451 | 0.993 | 0.935 | 0.02458462 | FABP4hi AM |
| CFD | 0.73048571 | 0.996 | 0.952 | 1.60E-09 | FABP4hi AM |
| CTSL | 0.71147157 | 0.996 | 0.96 | 3.90E-15 | FABP4hi AM |
| FTL | 0.69081052 | 1 | 1 | 5.87E-19 | FABP4hi AM |
| DEFB1 | 0.60382772 | 0.642 | 0.329 | 3.26E-09 | FABP4hi AM |
| SERPING1 | 0.58483023 | 0.97 | 0.809 | 1.64E-12 | FABP4hi AM |
| CSTB | 0.58448538 | 1 | 0.998 | 3.31E-07 | FABP4hi AM |
| MT2A | 0.57271389 | 0.816 | 0.594 | 0.01376845 | FABP4hi AM |
| LGALS3 | 0.55278247 | 1 | 0.999 | 2.06E-18 | FABP4hi AM |
| AC026369.3 | 0.54966423 | 0.957 | 0.793 | 1.55E-10 | FABP4hi AM |
| MSR1 | 0.49540699 | 0.987 | 0.92 | 0.00023016 | FABP4hi AM |
| KLHDC8B | 0.48982898 | 0.631 | 0.226 | 9.83E-09 | FABP4hi AM |
| ASAH1 | 0.4833787 | 0.996 | 0.974 | 1.36E-11 | FABP4hi AM |
| CTSD | 0.48148652 | 1 | 0.992 | 7.76E-10 | FABP4hi AM |
| GPNMB | 0.47684371 | 0.991 | 0.92 | 7.80E-08 | FABP4hi AM |
| TFRC | 0.46615556 | 0.945 | 0.846 | 0.01467574 | FABP4hi AM |
| OASL | 0.45978957 | 0.856 | 0.642 | 2.14E-10 | FABP4hi AM |
| AKR1C3 | 0.44206454 | 0.647 | 0.367 | 5.51E-07 | FABP4hi AM |
| PPDPF | 0.44069089 | 0.995 | 0.974 | 0.0014768 | FABP4hi AM |
| NR1H3 | 0.4313547 | 0.616 | 0.235 | 1.29E-08 | FABP4hi AM |
| MME | 0.430857 | 0.79 | 0.507 | 1.14E-07 | FABP4hi AM |
| NMB | 0.43007935 | 0.736 | 0.523 | 0.00025224 | FABP4hi AM |
| BLOC1S2 | 0.42941794 | 0.942 | 0.829 | 0.00077201 | FABP4hi AM |
| HP | 0.42836354 | 0.726 | 0.493 | 2.59E-07 | FABP4hi AM |
| CYB5A | 0.40800127 | 0.95 | 0.809 | 0.00039668 | FABP4hi AM |
| TSPAN4 | 0.4032426 | 0.745 | 0.433 | 0.02402353 | FABP4hi AM |
| RMDN3 | 0.39759646 | 0.916 | 0.736 | 3.17E-05 | FABP4hi AM |
| ALDH2 | 0.39698052 | 0.998 | 0.975 | 1.16E-07 | FABP4hi AM |
| CYP27A1 | 0.38930985 | 0.982 | 0.861 | 0.03362933 | FABP4hi AM |
| PLBD1 | 0.38406723 | 0.977 | 0.909 | 0.00026914 | FABP4hi AM |
| CD63 | 0.38292514 | 1 | 0.997 | 1.55E-06 | FABP4hi AM |
| ABCA1 | 0.38085089 | 0.557 | 0.162 | 0.0013516 | FABP4hi AM |
| TREM1 | 0.37566167 | 0.986 | 0.93 | 6.79E-10 | FABP4hi AM |
| CTSA | 0.35488938 | 0.922 | 0.802 | 0.01826696 | FABP4hi AM |
| C1QB | 0.35039206 | 0.999 | 0.968 | 1.30E-05 | FABP4hi AM |
| CD68 | 0.34777452 | 0.998 | 0.99 | 9.19E-06 | FABP4hi AM |
| JPT1 | 0.34214689 | 0.989 | 0.929 | 5.16E-07 | FABP4hi AM |
| C9orf16 | 0.335663 | 0.982 | 0.942 | 0.00359818 | FABP4hi AM |
| MGLL | 0.32542089 | 0.685 | 0.359 | 3.86E-07 | FABP4hi AM |
| ADTRP | 0.31960258 | 0.696 | 0.436 | 0.0008662 | FABP4hi AM |
| SCCPDH | 0.31573112 | 0.89 | 0.748 | 0.01298307 | FABP4hi AM |
| FTH1 | 0.31440797 | 1 | 1 | 2.08E-09 | FABP4hi AM |
| APOC2 | 0.30317967 | 0.554 | 0.337 | 0.01842905 | FABP4hi AM |
| CD59 | 0.29998005 | 0.833 | 0.605 | 0.00231673 | FABP4hi AM |
| ACO1 | 0.28956713 | 0.879 | 0.702 | 0.04243243 | FABP4hi AM |
| HLA-B | 0.2759101 | 1 | 1 | 3.95E-06 | FABP4hi AM |
| B2M | 0.274534 | 1 | 1 | 3.32E-09 | FABP4hi AM |
| HES2 | 0.27309766 | 0.345 | 0.059 | 0.00257447 | FABP4hi AM |
| C1QC | 0.27305314 | 0.998 | 0.949 | 0.00018371 | FABP4hi AM |
| FHL1 | 0.26707548 | 0.75 | 0.528 | 0.02937951 | FABP4hi AM |
| FAM89A | 0.26544072 | 0.935 | 0.801 | 5.33E-05 | FABP4hi AM |
| LRPAP1 | 0.26246957 | 0.992 | 0.962 | 0.00040896 | FABP4hi AM |
| TYROBP | 0.25059142 | 1 | 0.998 | 0.01564555 | FABP4hi AM |

| gene | avg_log2FC | pct.1 | pct.2 | p_val_adj | cluster |
|---|---|---|---|---|---|
| IFI27 | 1.33426874 | 0.609 | 0.416 | 5.67E-07 | IGF1+ AM |
| IGF1 | 1.22642333 | 0.487 | 0.118 | 3.83E-18 | IGF1+ AM |
| INHBA | 0.78391312 | 0.973 | 0.725 | 1.15E-16 | IGF1+ AM |
| STXBP2 | 0.74934879 | 0.991 | 0.939 | 1.12E-13 | IGF1+ AM |
| ITIH5 | 0.68663718 | 0.7 | 0.288 | 2.05E-19 | IGF1+ AM |
| HP | 0.64281352 | 0.776 | 0.498 | 1.02E-06 | IGF1+ AM |
| HLA-DRB1 | 0.55392121 | 1 | 0.999 | 3.87E-10 | IGF1+ AM |
| LY6E | 0.54903885 | 0.998 | 0.96 | 4.92E-12 | IGF1+ AM |
| MCEMP1 | 0.53108724 | 1 | 0.939 | 1.85E-12 | IGF1+ AM |
| PARAL1 | 0.52937172 | 0.932 | 0.705 | 2.83E-07 | IGF1+ AM |
| LINC02154 | 0.51274588 | 0.461 | 0.241 | 2.86E-05 | IGF1+ AM |
| HBEGF | 0.48338998 | 0.83 | 0.591 | 3.84E-07 | IGF1+ AM |
| THBS1 | 0.47972769 | 0.763 | 0.454 | 1.05E-09 | IGF1+ AM |
| MX1 | 0.47739662 | 0.911 | 0.698 | 6.30E-08 | IGF1+ AM |
| RPS4Y1 | 0.47451061 | 0.865 | 0.571 | 6.20E-05 | IGF1+ AM |
| GLDN | 0.46875747 | 0.926 | 0.73 | 3.43E-10 | IGF1+ AM |
| LGALS3BP | 0.46457088 | 0.986 | 0.88 | 4.86E-09 | IGF1+ AM |
| FOLR3 | 0.45957099 | 0.709 | 0.492 | 5.25E-05 | IGF1+ AM |
| HLA-DQB1 | 0.45925917 | 1 | 0.994 | 2.60E-12 | IGF1+ AM |
| PCOLCE2 | 0.4586705 | 0.948 | 0.747 | 6.59E-08 | IGF1+ AM |
| IFI6 | 0.45189669 | 0.995 | 0.945 | 0.00055868 | IGF1+ AM |
| EVL | 0.44340084 | 0.965 | 0.85 | 4.59E-08 | IGF1+ AM |
| IFIT1 | 0.44251142 | 0.773 | 0.516 | 7.28E-06 | IGF1+ AM |
| AQP3 | 0.4348639 | 0.958 | 0.782 | 7.79E-06 | IGF1+ AM |
| PRSS21 | 0.4324241 | 0.732 | 0.49 | 0.00151636 | IGF1+ AM |
| SERPING1 | 0.42144283 | 0.985 | 0.818 | 1.32E-06 | IGF1+ AM |
| GCA | 0.41912589 | 0.981 | 0.904 | 1.09E-07 | IGF1+ AM |
| RSPO3 | 0.41238799 | 0.51 | 0.222 | 7.34E-09 | IGF1+ AM |
| ALOX5AP | 0.40293967 | 0.999 | 0.978 | 4.30E-13 | IGF1+ AM |
| ALAS1 | 0.39776086 | 0.873 | 0.695 | 0.0010865 | IGF1+ AM |
| HCAR2 | 0.3940609 | 0.759 | 0.491 | 0.00063142 | IGF1+ AM |
| C8B | 0.38323823 | 0.535 | 0.272 | 0.00010039 | IGF1+ AM |
| IFIT3 | 0.38275134 | 0.937 | 0.798 | 6.53E-07 | IGF1+ AM |
| LPL | 0.37413968 | 0.898 | 0.753 | 0.00484595 | IGF1+ AM |
| HLA-DQA1 | 0.37229506 | 1 | 0.991 | 5.76E-06 | IGF1+ AM |
| NNMT | 0.37157452 | 0.314 | 0.095 | 1.09E-05 | IGF1+ AM |
| IFI44L | 0.36892142 | 0.687 | 0.44 | 3.36E-09 | IGF1+ AM |
| TMEM273 | 0.36837873 | 0.68 | 0.402 | 8.69E-09 | IGF1+ AM |
| SERPINA1 | 0.35668171 | 1 | 0.983 | 3.11E-09 | IGF1+ AM |
| CXCL16 | 0.35054696 | 0.997 | 0.954 | 2.34E-06 | IGF1+ AM |
| SLC19A3 | 0.3489634 | 0.489 | 0.205 | 8.50E-07 | IGF1+ AM |
| PPIC | 0.34644661 | 0.918 | 0.712 | 3.19E-06 | IGF1+ AM |
| PPARG | 0.34162841 | 0.978 | 0.883 | 3.59E-05 | IGF1+ AM |
| TXNIP | 0.33727687 | 1 | 0.992 | 2.36E-05 | IGF1+ AM |
| TPT1 | 0.33257666 | 1 | 1 | 8.44E-17 | IGF1+ AM |
| C1QA | 0.3311782 | 1 | 0.973 | 1.40E-06 | IGF1+ AM |
| RASAL2 | 0.32676102 | 0.779 | 0.583 | 2.73E-05 | IGF1+ AM |
| MRC1 | 0.32664339 | 1 | 0.977 | 0.00469264 | IGF1+ AM |
| SVIL | 0.31608293 | 0.939 | 0.796 | 1.99E-06 | IGF1+ AM |
| PAG1 | 0.30480946 | 0.773 | 0.619 | 0.00023291 | IGF1+ AM |
| HLA-DRA | 0.30316617 | 1 | 0.999 | 0.00027763 | IGF1+ AM |
| CYSLTR1 | 0.30114138 | 0.773 | 0.591 | 0.00039575 | IGF1+ AM |
| IFIT2 | 0.29941425 | 0.807 | 0.643 | 0.00089247 | IGF1+ AM |
| ARHGAP18 | 0.29488203 | 0.984 | 0.951 | 0.00183199 | IGF1+ AM |
| PYCARD | 0.29398411 | 0.998 | 0.974 | 5.30E-05 | IGF1+ AM |
| ATP1B1 | 0.2912707 | 0.958 | 0.848 | 8.32E-05 | IGF1+ AM |
| CLEC4A | 0.28808303 | 0.726 | 0.526 | 0.00011894 | IGF1+ AM |
| FAM89A | 0.28612574 | 0.947 | 0.809 | 0.00059124 | IGF1+ AM |
| PLAC8 | 0.28611764 | 0.734 | 0.552 | 0.0038156 | IGF1+ AM |
| ACKR3 | 0.27635352 | 0.583 | 0.393 | 0.00072323 | IGF1+ AM |
| CD81 | 0.26884907 | 1 | 0.993 | 0.00049947 | IGF1+ AM |
| CCND3 | 0.26501563 | 0.845 | 0.693 | 0.00033793 | IGF1+ AM |
| XAF1 | 0.25986575 | 0.837 | 0.689 | 0.00269561 | IGF1+ AM |
| CD74 | 0.25209239 | 1 | 1 | 0.00014774 | IGF1+ AM |
| RHBDD2 | 0.25060525 | 0.879 | 0.742 | 0.00242188 | IGF1+ AM |

| gene | avg_log2FC | pct.1 | pct.2 | p_val_adj | cluster |
|---|---|---|---|---|---|
| MSMO1 | 0.88942122 | 0.708 | 0.399 | 7.26E-08 | CD206hi FN1hi AM |
| FBP1 | 0.79807593 | 1 | 0.987 | 3.40E-21 | CD206hi FN1hi AM |
| FDPS | 0.66061358 | 0.799 | 0.627 | 1.02E-05 | CD206hi FN1hi AM |
| ACAT2 | 0.65011069 | 0.619 | 0.315 | 4.84E-06 | CD206hi FN1hi AM |
| FN1 | 0.63640431 | 0.947 | 0.886 | 7.78E-06 | CD206hi FN1hi AM |
| PCSK1N | 0.56046099 | 0.255 | 0.049 | 0.00611714 | CD206hi FN1hi AM |
| MRC1 | 0.5472296 | 0.999 | 0.978 | 9.05E-14 | CD206hi FN1hi AM |
| ALOX5AP | 0.53643882 | 0.998 | 0.979 | 1.27E-12 | CD206hi FN1hi AM |
| FDFT1 | 0.52793961 | 0.758 | 0.571 | 1.88E-06 | CD206hi FN1hi AM |
| HMGCS1 | 0.52291211 | 0.438 | 0.21 | 0.01717294 | CD206hi FN1hi AM |
| RGCC | 0.51655525 | 0.991 | 0.917 | 1.69E-06 | CD206hi FN1hi AM |
| MT-CYB | 0.50275822 | 0.999 | 0.999 | 2.50E-08 | CD206hi FN1hi AM |
| CD9 | 0.50174491 | 0.995 | 0.972 | 0.00041528 | CD206hi FN1hi AM |
| SQLE | 0.49559356 | 0.527 | 0.253 | 1.68E-06 | CD206hi FN1hi AM |
| MT-CO1 | 0.48969548 | 0.999 | 1 | 4.78E-15 | CD206hi FN1hi AM |
| IDI1 | 0.48713205 | 0.658 | 0.449 | 0.00097736 | CD206hi FN1hi AM |
| IGFBP2 | 0.48401192 | 0.846 | 0.741 | 0.00019074 | CD206hi FN1hi AM |
| MT-ATP6 | 0.48227274 | 0.998 | 0.998 | 2.81E-09 | CD206hi FN1hi AM |
| INSIG1 | 0.46557327 | 0.47 | 0.191 | 4.06E-06 | CD206hi FN1hi AM |
| S100A9 | 0.45524795 | 0.998 | 0.975 | 4.69E-08 | CD206hi FN1hi AM |
| GCA | 0.44961691 | 0.978 | 0.907 | 1.12E-05 | CD206hi FN1hi AM |
| S100A13 | 0.44666964 | 0.968 | 0.898 | 0.00105432 | CD206hi FN1hi AM |
| RETN | 0.44011776 | 0.982 | 0.92 | 5.88E-05 | CD206hi FN1hi AM |
| MT-CO2 | 0.43044145 | 1 | 0.999 | 5.85E-13 | CD206hi FN1hi AM |
| GLDN | 0.42978693 | 0.879 | 0.749 | 1.09E-05 | CD206hi FN1hi AM |
| MCEMP1 | 0.42690768 | 0.998 | 0.941 | 1.46E-08 | CD206hi FN1hi AM |
| DBI | 0.42118479 | 0.995 | 0.985 | 2.46E-07 | CD206hi FN1hi AM |
| ARMH1 | 0.42063392 | 0.607 | 0.345 | 0.00065096 | CD206hi FN1hi AM |
| S100A4 | 0.41965326 | 1 | 0.998 | 0.00324314 | CD206hi FN1hi AM |
| RAC2 | 0.41882807 | 0.915 | 0.794 | 0.00035856 | CD206hi FN1hi AM |
| EBP | 0.41353834 | 0.699 | 0.521 | 0.00176319 | CD206hi FN1hi AM |
| MT-CO3 | 0.41177952 | 1 | 0.999 | 2.52E-09 | CD206hi FN1hi AM |
| FADS1 | 0.40409234 | 0.536 | 0.281 | 0.00026417 | CD206hi FN1hi AM |
| GSN | 0.38237224 | 0.97 | 0.933 | 0.00197742 | CD206hi FN1hi AM |
| FKBP1A | 0.3814077 | 0.982 | 0.969 | 4.30E-10 | CD206hi FN1hi AM |
| NRP2 | 0.37865662 | 0.764 | 0.669 | 0.00719797 | CD206hi FN1hi AM |
| RHBDD2 | 0.37287601 | 0.867 | 0.75 | 7.15E-07 | CD206hi FN1hi AM |
| PTPMT1 | 0.3666586 | 0.966 | 0.885 | 0.00192938 | CD206hi FN1hi AM |
| LTA4H | 0.36198199 | 0.998 | 0.965 | 8.05E-05 | CD206hi FN1hi AM |
| AQP3 | 0.35918445 | 0.914 | 0.799 | 0.00179813 | CD206hi FN1hi AM |
| ACTG1 | 0.34866251 | 0.999 | 0.997 | 0.0015436 | CD206hi FN1hi AM |
| CAPG | 0.34177968 | 0.999 | 0.993 | 3.27E-09 | CD206hi FN1hi AM |
| ITGAX | 0.34155123 | 0.663 | 0.458 | 0.00507371 | CD206hi FN1hi AM |
| CYBB | 0.33524853 | 0.992 | 0.947 | 0.00947391 | CD206hi FN1hi AM |
| DCSTAMP | 0.33455688 | 0.514 | 0.294 | 0.00129268 | CD206hi FN1hi AM |
| MMAB | 0.33336471 | 0.53 | 0.308 | 0.00057565 | CD206hi FN1hi AM |
| FAM213A | 0.33297236 | 0.599 | 0.47 | 0.03019848 | CD206hi FN1hi AM |
| HCST | 0.33244045 | 0.959 | 0.927 | 0.00372168 | CD206hi FN1hi AM |
| S1PR4 | 0.33039564 | 0.717 | 0.575 | 0.00601306 | CD206hi FN1hi AM |
| ACTB | 0.31797569 | 1 | 1 | 0.00014697 | CD206hi FN1hi AM |
| HMGCR | 0.31426822 | 0.45 | 0.248 | 1.77E-09 | CD206hi FN1hi AM |
| RNF149 | 0.30584333 | 0.967 | 0.926 | 0.02762825 | CD206hi FN1hi AM |
| KAT8 | 0.30432615 | 0.808 | 0.676 | 0.00292038 | CD206hi FN1hi AM |
| TKT | 0.30192358 | 0.99 | 0.962 | 3.00E-05 | CD206hi FN1hi AM |
| TSPO | 0.29950728 | 1 | 0.997 | 1.09E-08 | CD206hi FN1hi AM |
| HPGD | 0.29047632 | 0.702 | 0.539 | 0.00250191 | CD206hi FN1hi AM |
| LDLR | 0.27716659 | 0.334 | 0.109 | 0.00432217 | CD206hi FN1hi AM |
| PPIA | 0.27356946 | 0.996 | 0.995 | 0.00164853 | CD206hi FN1hi AM |
| MT-ND5 | 0.27200651 | 0.998 | 0.996 | 8.24E-05 | CD206hi FN1hi AM |
| COMT | 0.26681225 | 0.981 | 0.941 | 0.00185025 | CD206hi FN1hi AM |
| CALM1 | 0.26135145 | 0.999 | 0.996 | 0.00026172 | CD206hi FN1hi AM |
| BST2 | -0.2646273 | 0.946 | 0.956 | 0.012401 | CD206hi FN1hi AM |
| CYB5A | -0.270737 | 0.834 | 0.856 | 0.04604348 | CD206hi FN1hi AM |
| XAF1 | -0.2914263 | 0.609 | 0.754 | 0.00436296 | CD206hi FN1hi AM |
| KLHDC8B | -0.3198505 | 0.151 | 0.399 | 0.00241472 | CD206hi FN1hi AM |
| NR1H3 | -0.363053 | 0.16 | 0.4 | 0.00164168 | CD206hi FN1hi AM |
| PLTP | -0.3766048 | 0.07 | 0.232 | 0.00238656 | CD206hi FN1hi AM |
| RPS6 | -0.4070646 | 0.997 | 0.998 | 1.27E-05 | CD206hi FN1hi AM |
| HLA-DRB1 | -0.4212834 | 1 | 0.999 | 7.81E-06 | CD206hi FN1hi AM |
| CD99 | -0.4315832 | 0.77 | 0.903 | 0.00015869 | CD206hi FN1hi AM |
| EPSTI1 | -0.4444699 | 0.508 | 0.734 | 0.00011962 | CD206hi FN1hi AM |
| TMEM176A | -0.4475763 | 0.256 | 0.499 | 0.00158442 | CD206hi FN1hi AM |
| ABCA1 | -0.4610157 | 0.036 | 0.346 | 1.08E-10 | CD206hi FN1hi AM |
| IFI44L | -0.4644293 | 0.282 | 0.555 | 7.06E-06 | CD206hi FN1hi AM |
| TSPAN4 | -0.4747147 | 0.291 | 0.589 | 0.01901076 | CD206hi FN1hi AM |
| CTSH | -0.4865791 | 0.933 | 0.971 | 1.77E-10 | CD206hi FN1hi AM |
| PRSS21 | -0.5184937 | 0.33 | 0.604 | 0.00094291 | CD206hi FN1hi AM |
| MX1 | -0.527575 | 0.599 | 0.787 | 0.00766327 | CD206hi FN1hi AM |
| DEFB1 | -0.5516113 | 0.19 | 0.485 | 2.51E-08 | CD206hi FN1hi AM |
| CAMP | -0.5554348 | 0.078 | 0.298 | 3.50E-06 | CD206hi FN1hi AM |
| MARCKS | -0.5812892 | 0.073 | 0.277 | 0.00110131 | CD206hi FN1hi AM |
| RPS4Y1 | -0.6009603 | 0.392 | 0.706 | 0.00015022 | CD206hi FN1hi AM |
| TMEM176B | -0.6145951 | 0.451 | 0.655 | 6.32E-05 | CD206hi FN1hi AM |
| SCD | -0.6971859 | 0.758 | 0.813 | 0.0003798 | CD206hi FN1hi AM |
| CFD | -0.7184183 | 0.962 | 0.966 | 8.44E-06 | CD206hi FN1hi AM |
| STMN1 | -0.8384048 | 0.787 | 0.864 | 0.0032456 | CD206hi FN1hi AM |
| APOC1 | -0.8446825 | 1 | 0.989 | 3.85E-07 | CD206hi FN1hi AM |
| IFITM3 | -0.867519 | 0.483 | 0.806 | 5.77E-15 | CD206hi FN1hi AM |
| NUPR1 | -0.8991428 | 0.678 | 0.84 | 3.52E-08 | CD206hi FN1hi AM |
| HP | -0.9189205 | 0.349 | 0.62 | 0.00187118 | CD206hi FN1hi AM |

| gene | avg_log2FC | pct.1 | pct.2 | p_val_adj | cluster |
|---|---|---|---|---|---|
| CXCL10 | 3.67906636 | 0.603 | 0.05 | 9.98E-25 | CXCL10+ AM |
| CCL4 | 3.2923231 | 0.523 | 0.115 | 2.84E-13 | CXCL10+ AM |
| CCL4L2 | 2.59314687 | 0.624 | 0.133 | 2.43E-20 | CXCL10+ AM |
| CCL20 | 1.89948755 | 0.376 | 0.079 | 1.30E-07 | CXCL10+ AM |
| SOD2 | 1.89766014 | 0.958 | 0.56 | 1.34E-38 | CXCL10+ AM |
| CXCL9 | 1.69905095 | 0.223 | 0.024 | 8.08E-06 | CXCL10+ AM |
| GBP1 | 1.56455854 | 0.929 | 0.694 | 4.27E-22 | CXCL10+ AM |
| CCL3 | 1.38861351 | 0.424 | 0.126 | 1.15E-07 | CXCL10+ AM |
| TNFAIP6 | 1.36609254 | 0.513 | 0.033 | 2.07E-18 | CXCL10+ AM |
| NCF1 | 1.28139023 | 0.889 | 0.656 | 1.88E-20 | CXCL10+ AM |
| MT2A | 1.26179451 | 0.872 | 0.653 | 1.81E-09 | CXCL10+ AM |
| CXCL11 | 1.17137049 | 0.269 | 0.011 | 8.16E-10 | CXCL10+ AM |
| IFIT2 | 1.15504397 | 0.817 | 0.676 | 0.0004493 | CXCL10+ AM |
| VAMP5 | 1.08329034 | 0.754 | 0.538 | 1.37E-12 | CXCL10+ AM |
| C15orf48 | 1.08053434 | 0.853 | 0.548 | 3.07E-08 | CXCL10+ AM |
| MARCKS | 1.02433928 | 0.681 | 0.219 | 6.26E-14 | CXCL10+ AM |
| MIR3945HG | 0.98746313 | 0.723 | 0.446 | 0.00811435 | CXCL10+ AM |
| IFITM3 | 0.93571362 | 0.916 | 0.732 | 1.03E-10 | CXCL10+ AM |
| ISG15 | 0.93151892 | 0.889 | 0.852 | 0.03082515 | CXCL10+ AM |
| SNX10 | 0.9082322 | 0.981 | 0.968 | 7.03E-17 | CXCL10+ AM |
| TNIP3 | 0.90295356 | 0.38 | 0.031 | 2.49E-11 | CXCL10+ AM |
| IFIT3 | 0.88531668 | 0.918 | 0.828 | 8.53E-05 | CXCL10+ AM |
| NUPR1 | 0.83589913 | 0.926 | 0.802 | 8.11E-09 | CXCL10+ AM |
| PLEK | 0.81611936 | 0.926 | 0.809 | 3.25E-10 | CXCL10+ AM |
| PNRC1 | 0.81513115 | 0.956 | 0.861 | 1.08E-07 | CXCL10+ AM |
| GBP4 | 0.80328473 | 0.718 | 0.434 | 9.63E-14 | CXCL10+ AM |
| CCL23 | 0.78055915 | 0.765 | 0.57 | 0.01212619 | CXCL10+ AM |
| STAT1 | 0.77235579 | 0.916 | 0.733 | 2.11E-13 | CXCL10+ AM |
| WARS | 0.75626819 | 0.821 | 0.595 | 3.09E-09 | CXCL10+ AM |
| GBP5 | 0.74057673 | 0.485 | 0.188 | 0.00427134 | CXCL10+ AM |
| TNFSF13B | 0.71974288 | 0.922 | 0.767 | 6.26E-13 | CXCL10+ AM |
| RARRES3 | 0.70699858 | 0.744 | 0.524 | 3.08E-06 | CXCL10+ AM |
| TMEM176B | 0.68800514 | 0.771 | 0.607 | 0.00439685 | CXCL10+ AM |
| NINJ1 | 0.67540538 | 0.645 | 0.231 | 1.16E-13 | CXCL10+ AM |
| EPSTI1 | 0.67507335 | 0.861 | 0.68 | 9.92E-11 | CXCL10+ AM |
| PSME2 | 0.66906023 | 0.981 | 0.977 | 1.65E-13 | CXCL10+ AM |
| TNFAIP2 | 0.65392758 | 0.91 | 0.75 | 3.10E-12 | CXCL10+ AM |
| LAP3 | 0.64923009 | 0.962 | 0.922 | 1.74E-16 | CXCL10+ AM |
| IL4I1 | 0.64339844 | 0.546 | 0.105 | 2.16E-14 | CXCL10+ AM |
| PSMB9 | 0.63505015 | 0.962 | 0.888 | 7.85E-14 | CXCL10+ AM |
| CD40 | 0.63353616 | 0.819 | 0.606 | 1.43E-06 | CXCL10+ AM |
| SLAMF8 | 0.63086118 | 0.739 | 0.344 | 1.23E-09 | CXCL10+ AM |
| ISG20 | 0.62973676 | 0.321 | 0.07 | 7.96E-09 | CXCL10+ AM |
| APOBEC3A | 0.62651129 | 0.246 | 0.022 | 6.77E-07 | CXCL10+ AM |
| IL1RN | 0.61994717 | 0.664 | 0.523 | 0.02873019 | CXCL10+ AM |
| RSAD2 | 0.61507123 | 0.363 | 0.136 | 5.63E-10 | CXCL10+ AM |
| SAT1 | 0.61231005 | 0.994 | 0.985 | 3.59E-05 | CXCL10+ AM |
| BCL2A1 | 0.58212791 | 0.964 | 0.904 | 0.01013968 | CXCL10+ AM |
| NBN | 0.53882933 | 0.725 | 0.527 | 0.00037928 | CXCL10+ AM |
| TYMP | 0.53862846 | 0.989 | 0.973 | 1.18E-11 | CXCL10+ AM |
| PARP14 | 0.52870257 | 0.845 | 0.729 | 4.13E-07 | CXCL10+ AM |
| ICAM1 | 0.52710639 | 0.532 | 0.174 | 0.00071207 | CXCL10+ AM |
| CCL3L1 | 0.52698913 | 0.183 | 0.032 | 0.0153979 | CXCL10+ AM |
| STOM | 0.52228182 | 0.91 | 0.721 | 4.82E-07 | CXCL10+ AM |
| KYNU | 0.51398167 | 0.794 | 0.598 | 0.03590296 | CXCL10+ AM |
| IFI44L | 0.51362129 | 0.742 | 0.489 | 2.58E-06 | CXCL10+ AM |
| DRAM1 | 0.50500999 | 0.773 | 0.519 | 5.00E-05 | CXCL10+ AM |
| FCGR1A | 0.49467586 | 0.96 | 0.888 | 0.00118799 | CXCL10+ AM |
| HLA-A | 0.49314341 | 1 | 0.997 | 5.99E-11 | CXCL10+ AM |
| OPTN | 0.49158359 | 0.782 | 0.572 | 3.44E-06 | CXCL10+ AM |
| NAMPT | 0.49048969 | 0.899 | 0.799 | 0.00019971 | CXCL10+ AM |
| TAP1 | 0.4793627 | 0.664 | 0.372 | 9.15E-09 | CXCL10+ AM |
| UGCG | 0.47828353 | 0.603 | 0.368 | 6.88E-06 | CXCL10+ AM |
| NEXN | 0.46819913 | 0.456 | 0.237 | 5.08E-05 | CXCL10+ AM |
| IFIH1 | 0.467313 | 0.63 | 0.413 | 3.52E-05 | CXCL10+ AM |
| PSTPIP2 | 0.46492967 | 0.464 | 0.111 | 2.90E-11 | CXCL10+ AM |
| ATOX1 | 0.45638517 | 0.979 | 0.953 | 5.88E-05 | CXCL10+ AM |
| SLC1A3 | 0.44601398 | 0.515 | 0.197 | 1.11E-06 | CXCL10+ AM |
| RNF144B | 0.44420562 | 0.567 | 0.286 | 2.83E-06 | CXCL10+ AM |
| SP110 | 0.43742944 | 0.884 | 0.736 | 9.79E-06 | CXCL10+ AM |
| DYNLT1 | 0.43416114 | 0.956 | 0.885 | 3.66E-07 | CXCL10+ AM |
| CALHM6 | 0.43002474 | 0.422 | 0.164 | 9.84E-05 | CXCL10+ AM |
| DNAJA1 | 0.42536095 | 0.992 | 0.968 | 6.61E-05 | CXCL10+ AM |
| C3AR1 | 0.42405643 | 0.84 | 0.626 | 0.00014354 | CXCL10+ AM |
| FYB1 | 0.42002174 | 0.834 | 0.67 | 0.00045196 | CXCL10+ AM |
| HLA-F | 0.40462327 | 0.916 | 0.8 | 0.00072424 | CXCL10+ AM |
| SQSTM1 | 0.40316485 | 0.956 | 0.906 | 0.02501699 | CXCL10+ AM |
| APOC1 | 0.40310451 | 1 | 0.991 | 0.01180596 | CXCL10+ AM |
| GCH1 | 0.4013586 | 0.405 | 0.066 | 2.58E-09 | CXCL10+ AM |
| RARRES1 | 0.39928901 | 0.41 | 0.142 | 1.15E-06 | CXCL10+ AM |
| CD274 | 0.39650919 | 0.479 | 0.209 | 0.00140319 | CXCL10+ AM |
| LILRB4 | 0.37478647 | 0.63 | 0.396 | 0.01068485 | CXCL10+ AM |
| FCGR3A | 0.3733433 | 0.975 | 0.966 | 0.00261326 | CXCL10+ AM |
| STAT2 | 0.36645442 | 0.748 | 0.58 | 0.01641807 | CXCL10+ AM |
| HLA-E | 0.36520062 | 0.996 | 0.991 | 1.03E-11 | CXCL10+ AM |
| ATF5 | 0.362817 | 0.508 | 0.26 | 3.25E-08 | CXCL10+ AM |
| HLA-DQA1 | 0.36090267 | 1 | 0.993 | 0.0021059 | CXCL10+ AM |
| IL27 | 0.35940214 | 0.29 | 0.073 | 3.81E-05 | CXCL10+ AM |
| TSPAN4 | 0.35873306 | 0.71 | 0.52 | 0.01061291 | CXCL10+ AM |
| CD48 | 0.3567167 | 0.46 | 0.18 | 0.00029376 | CXCL10+ AM |

| gene | avg_log2FC | pct.1 | pct.2 | p_val_adj | cluster |
|---|---|---|---|---|---|
| SPP1 | 3.06654279 | 0.176 | 0.022 | 0.00389499 | mono-SPP1+ AM |
| CCL2 | 3.04549827 | 0.309 | 0.017 | 7.90E-11 | mono-SPP1+ AM |
| RNASE1 | 2.61351973 | 0.494 | 0.083 | 4.77E-16 | mono-SPP1+ AM |
| VCAN | 1.69126075 | 0.694 | 0.274 | 1.22E-15 | mono-SPP1+ AM |
| C15orf48 | 1.69125006 | 0.713 | 0.532 | 3.84E-11 | mono-SPP1+ AM |
| TIMP1 | 1.67325112 | 0.888 | 0.887 | 3.25E-06 | mono-SPP1+ AM |
| FCN1 | 1.58420441 | 0.489 | 0.099 | 9.19E-15 | mono-SPP1+ AM |
| SGK1 | 1.34394964 | 0.69 | 0.462 | 2.13E-11 | mono-SPP1+ AM |
| CCL3 | 1.3339694 | 0.275 | 0.113 | 0.00855896 | mono-SPP1+ AM |
| IER3 | 1.26818066 | 0.404 | 0.191 | 0.00057057 | mono-SPP1+ AM |
| S100A8 | 1.2610303 | 0.862 | 0.91 | 0.00167967 | mono-SPP1+ AM |
| EMP3 | 1.24989128 | 0.985 | 0.984 | 9.96E-22 | mono-SPP1+ AM |
| MARCKS | 1.24255067 | 0.481 | 0.193 | 5.38E-12 | mono-SPP1+ AM |
| EMP1 | 1.23947463 | 0.494 | 0.068 | 3.72E-18 | mono-SPP1+ AM |
| MS4A6A | 1.18137082 | 0.824 | 0.738 | 4.60E-10 | mono-SPP1+ AM |
| PLA2G7 | 1.14459097 | 0.349 | 0.042 | 3.46E-09 | mono-SPP1+ AM |
| ZFP36L1 | 1.05921034 | 0.603 | 0.344 | 1.96E-07 | mono-SPP1+ AM |
| FCGR2B | 1.03905019 | 0.386 | 0.035 | 6.28E-14 | mono-SPP1+ AM |
| FPR3 | 1.01574688 | 0.642 | 0.299 | 3.85E-15 | mono-SPP1+ AM |
| MAFB | 1.00920158 | 0.715 | 0.534 | 1.05E-09 | mono-SPP1+ AM |
| NEAT1 | 0.9926773 | 0.987 | 0.986 | 7.66E-10 | mono-SPP1+ AM |
| KLF6 | 0.98820503 | 0.889 | 0.854 | 3.62E-09 | mono-SPP1+ AM |
| FGL2 | 0.98179322 | 0.539 | 0.233 | 4.74E-16 | mono-SPP1+ AM |
| TMEM176B | 0.93754108 | 0.784 | 0.583 | 5.36E-12 | mono-SPP1+ AM |
| AP1S2 | 0.93322412 | 0.776 | 0.598 | 5.50E-12 | mono-SPP1+ AM |
| CD14 | 0.93279566 | 0.84 | 0.824 | 0.01786884 | mono-SPP1+ AM |
| CLEC5A | 0.91497548 | 0.415 | 0.062 | 9.52E-14 | mono-SPP1+ AM |
| TGFBI | 0.91019596 | 0.742 | 0.67 | 0.00033565 | mono-SPP1+ AM |
| CORO1A | 0.89605467 | 0.511 | 0.176 | 2.55E-06 | mono-SPP1+ AM |
| CTSB | 0.84494457 | 0.987 | 0.994 | 1.11E-11 | mono-SPP1+ AM |
| LMNA | 0.84161496 | 0.86 | 0.912 | 4.29E-06 | mono-SPP1+ AM |
| TYMP | 0.83670572 | 0.969 | 0.974 | 2.32E-12 | mono-SPP1+ AM |
| CD84 | 0.83545357 | 0.58 | 0.404 | 4.66E-08 | mono-SPP1+ AM |
| PLEKHO1 | 0.81148678 | 0.571 | 0.301 | 3.64E-13 | mono-SPP1+ AM |
| PMP22 | 0.80091696 | 0.575 | 0.336 | 2.67E-09 | mono-SPP1+ AM |
| S100A10 | 0.78053182 | 0.999 | 1 | 2.50E-16 | mono-SPP1+ AM |
| CD44 | 0.77970423 | 0.972 | 0.98 | 8.07E-12 | mono-SPP1+ AM |
| LIMS1 | 0.77604423 | 0.842 | 0.809 | 2.30E-12 | mono-SPP1+ AM |
| ALCAM | 0.74623222 | 0.781 | 0.726 | 2.03E-07 | mono-SPP1+ AM |
| HM13 | 0.74296783 | 0.703 | 0.602 | 8.30E-11 | mono-SPP1+ AM |
| ANPEP | 0.73765214 | 0.605 | 0.396 | 1.20E-09 | mono-SPP1+ AM |
| RBPJ | 0.71889526 | 0.891 | 0.906 | 7.40E-10 | mono-SPP1+ AM |
| BASP1 | 0.71160443 | 0.319 | 0.076 | 4.87E-08 | mono-SPP1+ AM |
| GPR183 | 0.70628225 | 0.233 | 0.011 | 2.47E-08 | mono-SPP1+ AM |
| RGS10 | 0.69755372 | 0.827 | 0.8 | 0.00165878 | mono-SPP1+ AM |
| LILRB4 | 0.69264598 | 0.54 | 0.381 | 0.00043403 | mono-SPP1+ AM |
| DUSP6 | 0.69090936 | 0.548 | 0.416 | 2.07E-07 | mono-SPP1+ AM |
| MALAT1 | 0.68638988 | 1 | 0.999 | 1.94E-14 | mono-SPP1+ AM |
| ITGAM | 0.6825095 | 0.577 | 0.307 | 4.72E-11 | mono-SPP1+ AM |
| CYBB | 0.68209455 | 0.953 | 0.957 | 2.51E-06 | mono-SPP1+ AM |
| AHNAK | 0.67691555 | 0.849 | 0.855 | 6.03E-10 | mono-SPP1+ AM |
| CSF1R | 0.67085595 | 0.546 | 0.341 | 0.00223514 | mono-SPP1+ AM |
| METRNL | 0.67055467 | 0.672 | 0.59 | 0.00023772 | mono-SPP1+ AM |
| CCR1 | 0.66601025 | 0.584 | 0.385 | 5.60E-06 | mono-SPP1+ AM |
| SPRED1 | 0.65968076 | 0.565 | 0.453 | 0.04676135 | mono-SPP1+ AM |
| LGALS1 | 0.65833822 | 0.987 | 0.994 | 9.87E-05 | mono-SPP1+ AM |
| TMEM176A | 0.63983355 | 0.609 | 0.421 | 2.92E-06 | mono-SPP1+ AM |
| LHFPL2 | 0.62491238 | 0.472 | 0.284 | 0.0026947 | mono-SPP1+ AM |
| CPM | 0.62474825 | 0.674 | 0.683 | 0.00134785 | mono-SPP1+ AM |
| SH3BGRL3 | 0.62431217 | 0.998 | 1 | 4.16E-16 | mono-SPP1+ AM |
| SDC2 | 0.61867034 | 0.258 | 0.067 | 0.00465693 | mono-SPP1+ AM |
| ZEB2 | 0.61850624 | 0.897 | 0.9 | 0.00014429 | mono-SPP1+ AM |
| FYB1 | 0.59955589 | 0.706 | 0.671 | 0.00013367 | mono-SPP1+ AM |
| LIMD2 | 0.58848548 | 0.322 | 0.036 | 1.91E-07 | mono-SPP1+ AM |
| HMGA1 | 0.58573305 | 0.514 | 0.359 | 1.78E-07 | mono-SPP1+ AM |
| QPCT | 0.58315627 | 0.422 | 0.16 | 1.63E-06 | mono-SPP1+ AM |
| TPM4 | 0.56475136 | 0.904 | 0.918 | 3.67E-09 | mono-SPP1+ AM |
| MEF2C | 0.56197165 | 0.35 | 0.126 | 0.00022714 | mono-SPP1+ AM |
| CALM1 | 0.55935236 | 0.988 | 0.998 | 4.59E-10 | mono-SPP1+ AM |
| TPM3 | 0.55895803 | 0.973 | 0.992 | 1.01E-09 | mono-SPP1+ AM |
| ARL4C | 0.54322597 | 0.265 | 0.091 | 0.02489627 | mono-SPP1+ AM |
| FOSB | 0.54210894 | 0.332 | 0.165 | 0.00020694 | mono-SPP1+ AM |
| ENO1 | 0.5330745 | 0.95 | 0.972 | 2.31E-07 | mono-SPP1+ AM |
| AQP9 | 0.51790824 | 0.503 | 0.382 | 0.02442557 | mono-SPP1+ AM |
| NPC2 | 0.51677327 | 0.987 | 0.994 | 1.32E-10 | mono-SPP1+ AM |
| VMP1 | 0.50658957 | 0.662 | 0.678 | 0.00250059 | mono-SPP1+ AM |
| CLEC10A | 0.50163388 | 0.183 | 0.005 | 4.05E-05 | mono-SPP1+ AM |
| SOCS3 | 0.5001958 | 0.341 | 0.151 | 0.01323303 | mono-SPP1+ AM |
| GAPDH | 0.49054864 | 0.991 | 0.999 | 3.85E-06 | mono-SPP1+ AM |
| CD48 | 0.48961721 | 0.295 | 0.171 | 0.00448218 | mono-SPP1+ AM |
| IGSF6 | 0.47886357 | 0.922 | 0.951 | 0.00482695 | mono-SPP1+ AM |
| PTMA | 0.45471931 | 0.999 | 1 | 1.97E-12 | mono-SPP1+ AM |
| MATK | 0.45304248 | 0.232 | 0.026 | 5.35E-06 | mono-SPP1+ AM |
| YWHAZ | 0.44875801 | 0.938 | 0.952 | 0.03304873 | mono-SPP1+ AM |
| ACTB | 0.44864475 | 0.999 | 1 | 3.57E-10 | mono-SPP1+ AM |
| SAMHD1 | 0.44631939 | 0.938 | 0.976 | 5.64E-05 | mono-SPP1+ AM |
| EMB | 0.42415178 | 0.246 | 0.033 | 2.12E-05 | mono-SPP1+ AM |
| LGALS2 | 0.42324916 | 0.162 | 0.036 | 0.00286765 | mono-SPP1+ AM |
| IQGAP1 | 0.41641315 | 0.884 | 0.917 | 0.00059016 | mono-SPP1+ AM |
| H3F3B | 0.4152059 | 0.993 | 0.997 | 0.00668188 | mono-SPP1+ AM |

| | | | | | |
|---|---|---|---|---|---|
| IFI27 | -1.7068598 | 0.277 | 0.51 | 4.57E-06 | CD206hi FN1hi AM |

| | | | | | |
|---|---|---|---|---|---|
| RNF213 | 0.35526368 | 0.95 | 0.879 | 0.01015312 | CXCL10+ AM |
| USP30-AS1 | 0.35117319 | 0.712 | 0.478 | 9.37E-05 | CXCL10+ AM |
| AQP9 | 0.3477427 | 0.664 | 0.39 | 0.00012053 | CXCL10+ AM |
| SRI | 0.34666368 | 0.943 | 0.883 | 0.00021129 | CXCL10+ AM |
| UBE2L6 | 0.34638623 | 0.908 | 0.81 | 0.04465891 | CXCL10+ AM |
| SLC39A8 | 0.34010571 | 0.332 | 0.068 | 1.01E-07 | CXCL10+ AM |
| LACTB | 0.33969821 | 0.945 | 0.836 | 0.00852468 | CXCL10+ AM |
| RELB | 0.3338458 | 0.435 | 0.097 | 7.53E-10 | CXCL10+ AM |
| APOL6 | 0.32997191 | 0.836 | 0.765 | 0.00401825 | CXCL10+ AM |
| CTSS | 0.31678927 | 0.998 | 0.996 | 9.12E-07 | CXCL10+ AM |
| BAZ1A | 0.31398941 | 0.828 | 0.696 | 0.0041716 | CXCL10+ AM |
| NUB1 | 0.30499182 | 0.842 | 0.701 | 0.00734067 | CXCL10+ AM |
| SAMSN1 | 0.29970342 | 0.536 | 0.342 | 0.01465078 | CXCL10+ AM |
| FTH1 | 0.29325056 | 1 | 1 | 6.99E-05 | CXCL10+ AM |
| RNF19B | 0.28356368 | 0.435 | 0.231 | 0.00137758 | CXCL10+ AM |
| TIFA | 0.28318769 | 0.433 | 0.223 | 3.90E-05 | CXCL10+ AM |
| SLAMF7 | 0.26524762 | 0.231 | 0.019 | 0.01242999 | CXCL10+ AM |
| XAF1 | 0.26271229 | 0.842 | 0.72 | 0.01836721 | CXCL10+ AM |
| FCGR1B | 0.25807821 | 0.735 | 0.59 | 0.02634688 | CXCL10+ AM |
| HLA-B | 0.25169953 | 1 | 1 | 7.67E-07 | CXCL10+ AM |
| ABCA1 | 0.25026424 | 0.492 | 0.274 | 0.00222694 | CXCL10+ AM |

| | | | | | |
|---|---|---|---|---|---|
| TLR2 | 0.41336988 | 0.265 | 0.09 | 0.00126134 | mono-SPP1+ AM |
| PPIB | 0.41317093 | 0.948 | 0.984 | 0.00100857 | mono-SPP1+ AM |
| F13A1 | 0.40252367 | 0.126 | 0.003 | 0.04929727 | mono-SPP1+ AM |
| BCAT1 | 0.39423695 | 0.331 | 0.16 | 0.0217633 | mono-SPP1+ AM |
| CRIP2 | 0.39135652 | 0.188 | 0.024 | 0.02107053 | mono-SPP1+ AM |
| PDXK | 0.38908709 | 0.828 | 0.881 | 0.00068669 | mono-SPP1+ AM |
| CDV3 | 0.38661547 | 0.882 | 0.922 | 0.00451301 | mono-SPP1+ AM |
| C3AR1 | 0.38542126 | 0.63 | 0.633 | 0.01469651 | mono-SPP1+ AM |
| AP2S1 | 0.38022496 | 0.964 | 0.987 | 1.30E-13 | mono-SPP1+ AM |
| CST6 | 0.37921473 | 0.164 | 0.019 | 0.01171619 | mono-SPP1+ AM |
| AC020916.1 | 0.3769628 | 0.28 | 0.178 | 0.0301303 | mono-SPP1+ AM |
| FRMD4B | 0.37665363 | 0.425 | 0.338 | 0.01652342 | mono-SPP1+ AM |
| KCNN4 | 0.37232697 | 0.206 | 0.024 | 0.03889646 | mono-SPP1+ AM |
| RPL35 | 0.36563099 | 0.992 | 0.998 | 0.00069733 | mono-SPP1+ AM |
| HOMER3 | 0.36314467 | 0.29 | 0.119 | 0.02081804 | mono-SPP1+ AM |
| SUB1 | 0.36287534 | 0.91 | 0.955 | 0.03488859 | mono-SPP1+ AM |
| CDA | 0.35685505 | 0.219 | 0.066 | 0.02573869 | mono-SPP1+ AM |
| ELK3 | 0.35139343 | 0.244 | 0.087 | 0.00101143 | mono-SPP1+ AM |
| ACTG1 | 0.34815903 | 0.991 | 0.998 | 2.65E-06 | mono-SPP1+ AM |
| PRKCB | 0.34736363 | 0.264 | 0.067 | 6.68E-06 | mono-SPP1+ AM |
| RPLP0 | 0.34363937 | 0.975 | 0.994 | 0.00208057 | mono-SPP1+ AM |
| SNCA | 0.33804861 | 0.34 | 0.179 | 0.00392681 | mono-SPP1+ AM |
| IGFBP7 | 0.33523186 | 0.275 | 0.086 | 2.09E-08 | mono-SPP1+ AM |
| ATP5F1E | 0.33455493 | 0.999 | 1 | 1.01E-05 | mono-SPP1+ AM |
| ANTXR2 | 0.31808907 | 0.203 | 0.02 | 7.91E-06 | mono-SPP1+ AM |
| GAS6 | 0.31068986 | 0.185 | 0.014 | 3.51E-06 | mono-SPP1+ AM |
| GNG5 | 0.30913249 | 0.983 | 0.996 | 0.00321227 | mono-SPP1+ AM |
| CD93 | 0.29543816 | 0.144 | 0.028 | 0.01439857 | mono-SPP1+ AM |
| ARPC4 | 0.28850554 | 0.861 | 0.928 | 0.02688029 | mono-SPP1+ AM |
| RASSF2 | 0.28524735 | 0.18 | 0.012 | 3.35E-07 | mono-SPP1+ AM |
| COPE | 0.28359723 | 0.891 | 0.948 | 0.03704581 | mono-SPP1+ AM |
| PFN1 | 0.28229068 | 0.995 | 1 | 5.16E-05 | mono-SPP1+ AM |
| RPS24 | 0.28065361 | 0.999 | 1 | 0.00766283 | mono-SPP1+ AM |
| ARPC5 | 0.28029035 | 0.983 | 0.997 | 6.22E-10 | mono-SPP1+ AM |
| S100A6 | 0.26897565 | 1 | 1 | 0.00419038 | mono-SPP1+ AM |
| ANXA6 | 0.25261838 | 0.183 | 0.026 | 0.00242465 | mono-SPP1+ AM |
