## Supplementary material for "Type I IFN-activated lung monocytes and macrophages as initiators and drivers of fibrosis at the alveolar barrier in IPF": Suppl Table 3A

**Suppl Table 3A.** Annotation of macrophage subsets in BAL from Oxford Cohort A. Each macrophage subset was manually compared to macrophage populations in published datasets with similar transcriptional profiles, cluster annotation or proposed functional state. Top 50 cluster defining genes (ranked by Log2 fold change) from BAL AMs is shown and compared to top 20 cluster defining genes from relevant published datasets. CD206hi FN1hi AMs or an equivalent population has not been previously described in a single cell RNAseq dataset ]so no comparison is shown. For mono-SPP1 AMs comparison is made to monocyte and SPP1+ macs.

FABP4hi AM

| **Dataset** | **Oxford Cohort A BAL** | | | **Morell 2023 - BAL** | **Morse 2021 - Lung Digest Control only** | **Morse 2021 - Lung Digest IPF and Control** | **Mulder 2021 - Lung Digest plus mulitple tissues[a]** | **Zhang 2021 - BAL, Lung Digest plus multiple tissues[b]** | **Aegerter 2023 - Human myeloid scRNA seq key markers** | **Sikkema 2023 - Lung Cell Atlas** |
|---|---|---|---|---|---|---|---|---|---|---|
| **Annotation** | FABP4hi AM | | | Mature AM | Cluster 0 (FABP4$^{hi}$)[c] | Cluster 0 (FABP4$^{hi}$)[c] | Cluster 16 MoMac (Alveolar Macs)[c] | MRC1$^{+}$ FABP4$^{+}$ | AMs | Alveolar macrophages_marker |
| | FABP4 | INHBA | NMB | FABP4 | FABP4 | FABP4 | C1QC | APOC1 | FABP4 | CYP27A1 |
| | APOC1 | IGFBP2 | BLOC1S2 | IFI27 | C1QB | C1QB | C1QB | C1QB | SERPING1 | MARCO |
| | RBP4 | RPS4Y1 | HP | SERPING1 | C1QA | C1QA | APOE | C1QA | APOC1 | FABP4 |
| | SCD | HP | CYB5A | C1QB | FBP1 | FBP1 | FUCA1 | APOE | CD52 | |
| | CAMP | ITIH5 | TSPAN4 | RBP4 | INHBA | C1QC | C1QA | FABP4 | PCOLCE2 | |
| | CCL18 | IFI27 | RMDN3 | CCL18 | CD52 | CD52 | DNASE1L3 | HLA-DQA1 | INHBA | |
| | NUPR1 | LGALS3BP | ALDH2 | C1QA | C1QC | MCEMP1 | HLA-DPA1 | HLA-DPB1 | MME (CD10) | |
| | ABCG1 | AC026369.3 | CYP27A1 | CES1 | RP11-598F7.3 | APOC1 | ACP5 | ALOX5AP | CES1 | |
| | PLIN2 | MSR1 | PLBD1 | NUPR1 | APOC1 | INHBA | RNASE1 | HLA-DPA1 | GPD1 | |
| | GCHFR | KLHDC8B | CD63 | C1QC | CD74 | RP11---598F7.3 | HLA-DRB1 | HLA-DQB1 | RBP4 | |
| | CES1 | ASAH1 | | FN1 | HLA--DRA | ALOX5AP | CD74 | ACP5 | ITIH5 | |
| | APOE | CTSD | | AC026369.3 | MCEMP1 | SERPINA1 | HLA-DQA1 | GCHFR | SCD | |
| | CFD | GPNMB | | LGALS3BP | HLA-DQB1 | VSIG4 | PLD3 | UBB | | |
| | CTSL | TFRC | | AC009093.3 | MSR1 | MSR1 | CD63 | HLA-DRB1 | | |
| | FTL | OASL | | INHBA | RBP4 | GCHFR | CTSC | NUPR1 | | |
| | DEFB1 | AKR1C3 | | APOC1 | HLA-DRB1 | FTL | HLA-DRB5 | HLA-DRA | | |
| | SERPING1 | PPDPF | | PHLDA3 | ALOX5AP | GRN | HLA-DQA2 | MARCO | | |
| | MT2A | NR1H3 | | PDLIM1 | HLA-DPA1 | ACP5 | PTGDS | TMSB4X | | |
| | CSTB | MME | | TFRC (CD71) | GRN | TREM1 | HLA-DRA | HLA-DMA | | |
| | | | | ALDH2 | CD68 | CD68 | CCL18 | FBP1 | | |
| Proportion of cluster defining genes in specified reference dataset that are also present in Oxford BAL AMs' top 50 cluster defining genes | | | | 0.6 | 0.25 | 0.25 | 0.2 | 0.25 | 0.67 | 0.33 |

IGF1+ AM

| **Dataset** | **Oxford Cohort A BAL** | | | **Mould 2021 - BAL** |
|---|---|---|---|---|
| **Annotation** | IGF1+ AM | | | Cluster 1 |
| | IFI27 | IFI6 | SERPINA1 | IGF1 |
| | IGF1 | EVL | CXCL16 | IFI27 |
| | INHBA | IFIT1 | GPD1 | ITIH5 |
| | STXBP2 | AQP3 | SLC19A3 | C8B |
| | ITIH5 | PRSS21 | ALOX5 | INHBA |
| | HP | SERPING1 | PPIC | SLC19A3 |
| | HLA-DRB1 | GCA | PPARG | FBN1 |
| | LY6E | RSPO3 | TXNIP | IGFBP2 |
| | MCEMP1 | ALOX5AP | CPE | ALAS1 |
| | PARAL1 | ALAS1 | TPT1 | HP |
| | LINC02154 | HCAR2 | | VGLL3 |
| | HBEGF | C8B | | GNG11 |
| | THBS1 | IFIT3 | | |
| | MX1 | LPL | | |
| | RPS4Y1 | HLA-DQA1 | | |
| | GLDN | NNMT | | |
| | LGALS3BP | IFI44L | | |
| | FOLR3 | TMEM273 | | |
| | HLA-DQB1 | GSN | | |
| | PCOLCE2 | VMO1 | | |
| Proportion of cluster defining genes in specified reference dataset that are also present in Oxford BAL AMs' top 50 cluster defining genes | | | | 0.75 |

CXCL10+ AM

| **Dataset** | **Oxford Cohort A BAL** | | | **Morell 2023 - BAL** | **Mould 2021 - BAL** | **Mulder 2021 - Lung Digest plus mulitple tissues[a]** | **Mulder 2021 - Lung Digest plus mulitple tissues[a]** | **Zhang 2021 - BAL, Lung Digest plus multiple tissues[b]** |
|---|---|---|---|---|---|---|---|---|
| **Annotation** | CXCL10+ AM | | | IFN-related | Cluster 8 | Cluster 6 MoMac (IFI44L Macs)[c] | Cluster 4 MoMac (ISG Mono)[c] | CXCL10$^{+}$ CCL2$^{+}$ |
| | CXCL10 | SNX10 | LAP3 | CXCL10 | CCL4 | CXCL10 | CXCL10 | CCL2 |
| | CCL4 | TNIP3 | IL4I1 | CCL8 | CXCL10 | CXCL9 | ISG15 | CXCL10 |
| | CCL4L2 | IFIT3 | PSMB9 | ISG15 | CCL4L2 | GBP1 | IFIT1 | IFITM3 |
| | CCL20 | IFIT1 | CD40 | IFIT2 | SOD2 | IDO1 | CCL8 | ISG15 |
| | SOD2 | NUPR1 | SLAMF8 | MX1 | CCL3 | CXCL11 | IFIT2 | CCL8 |
| | CXCL9 | PLEK | ISG20 | IFIT1 | CCL20 | STAT1 | IFIT3 | IL1RN |
| | GBP1 | PNRC1 | APOBEC3A | IFIT3 | MARCKS | IL4I1 | RSAD2 | CTSL |
| | CCL3 | GBP4 | IRF1 | RSAD2 | TNFAIP6 | GBP5 | TNFSF10 | IFITM1 |
| | TNFAIP6 | CCL23 | IL1RN | TNFSF10 | GBP1 | WARS | ISG20 | CCL3 |
| | NCF1 | STAT1 | RSAD2 | IFITM1 | C15orf48 | MMP9 | CCL2 | RSAD2 |
| | MT2A | WARS | SAT1 | IFITM3 | TNIP3 | CD40 | IFI44L | IFI27 |
| | CXCL11 | APOE | | GBP1 | CXCL8 | PPA1 | APOBEC3A | APOBEC3A |
| | IFIT2 | GBP5 | | HERC5 | NCF1 | SLAMF7 | IFITM3 | ISG20 |
| | VAMP5 | TNFSF13B | | RNF213 | CXCL5 | ANKRD22 | IFI6 | IFIT3 |
| | C15orf48 | RARRES3 | | MX2 | CCL3L1 | SOD2 | GBP1 | CCL4 |
| | MARCKS | TMEM176B | | APOBEC3A | ICAM1 | PTGDS | CXCL11 | MX1 |
| | MIR3945HG | NINJ1 | | EPSTI1 | MIR3945HG | VAMP5 | HERC5 | SOD2 |
| | TNFSF10 | EPSTI1 | | TNFSF13B | ACSL1 | TAP1 | LY6E | IFIT2 |
| | IFITM3 | PSME2 | | IRF7 | NINJ1 | CCL8 | IL1RN | IFIT1 |
| | ISG15 | TNFAIP2 | | OAS1 | CCl23 | PARP | IFI44 | GBP1 |
| Proportion of cluster defining genes in specified reference dataset that are also present in Oxford BAL AMs' top 50 cluster defining genes | | | | 0.60 | 0.70 | 0.55 | 0.75 | 0.75 |

**Footnote**

a Spleen (2), Lung (7), Liver (7), Skin (5), Blood (2), Lymph Node (1), Kidney (3), Head and Neck (1), Tonsil (2), Colon (6), Stomach (1), Ascites (1), Breast (2), Pancreas (1)

b Synovial Tissue(2), Kidney (1), Colon (1), Lung Digest (1), BAL (1)

c Annotation used in text

BAL Bronchoalveolar Lavage Cells

Overlapping genes

| Mono-SPP1+ AM | | | | SPP1 Macrophage Equivalent* | | | | | | | Monocytes | | | | | |
|---|---|---|---|---|---|---|---|---|---|---|---|---|---|---|---|---|
| **Dataset** | **Oxford Cohort A BAL** | | | **Morell 2023 - BAL** | **Morell 2023 - BAL** | **Morse 2021 - Lung Digest Control Only** | **Morse 2021 - Lung Digest IPF and Control** | **Mulder 2021 - Lung Digest plus mulitple tissues[a]** | **Aegerter 2023 - Human myeloid scRNA seq key markers** | **Aegerter 2023 - Human myeloid scRNA seq key markers** | **Morell 2023 - BAL** | **Morse 2021 - Lung Digest Control Only** | **Mulder 2021 - Lung Digest plus mulitple tissues[a]** | **Zhang 2021 - BAL, Lung Digest plus multiple tissues[b]** | **Aegerter 2023 - Human myeloid scRNA seq key markers** | **Sikkema 2023 - Lung Cell Atlas** |
| **Annotation** | Mono-SPP1$^{+}$ AM | | | Matricellular | LGMN/CD163 | Cluster 1 (SPP1$^{hi}$)[c] | Cluster 1 (SPP1$^{hi}$)[c] | Cluster 3 MoMac (TREM2)[c] | IM (Subtype 1) | IM (Subtype 2) | FCN1 | FCN1 Mono[c] | Cluster 21 MoMac (cMono)[c] | FCN1+ | recMac | Classical monocytes_ marker |
| | SPP1 | FPR3 | S100A10 | SPP1 | RNASE1 | APOE | SPP1 | APOC1 | LGMN | LGMN | FCN1 | S100A8 | S100A8 | S100A9 | S100A8 | S100A12 |
| | CCL2 | MAFB | CD44 | CHIT1 | LGMN | CCL18 | APOE | APOE | MARCKS | MARCKS | RETN | S100A9 | S100A9 | FCN1 | FN1 | FCN1 |
| | RNASE1 | NEAT1 | LIMS1 | LPL | CCL13 | HMOX1 | CCL18 | ACP5 | FOLR2 | SPP1 | LYZ | SERPINB2 | S100A12 | S100A12 | VCAN | RNASE2 |
| | VCAN | KLF6 | ALCAM | CD9 | HMOX1 | CTSB | CTSB | SPP1 | SELENOP | PLA2G7 | CORO1A | THBS1 | VCAN | VCAN | RNASE1 | |
| | C15orf48 | FGL2 | HM13 | CHI3L1 | CCL2 | CTSL | GPNMB | CTSD | F13A1 | MMP9 | LST1 | S100A12 | FCN1 | S100A4 | FCN1 | |
| | TIMP1 | TMEM176B | ANPEP | MMP7 | CTSL | CTSD | CTSL | GPNMB | SLC40A1 | HAMP | VSIR | IL1B | CSTA | IFITM2 | EMP1 | |
| | FCN1 | AP1S2 | IFITM3 | GSN | PLTP | CCL20 | CTSD | FABP5 | | | VCAN | EREG | RETN | APOBEC3A | CD14 | |
| | SGK1 | CD14 | RBPJ | TREM2 | SPP1 | FTL | LGMN | LGALS3 | | | AIF1 | FCN1 | S100A4 | TIMP1 | | |
| | CCL3 | CLEC5A | BASP1 | LIPA | CD163 | LGALS1 | FTL | CTSB | | | FOS | G0S2 | FPR1 | FOS | | |
| | IER3 | TGFBI | GPR183 | MATK | FOLR2 | GPNMB | APOC1 | PLA2G7 | | | ASGR1 | VCAN | CFP | CORO1A | | |
| | CCL13 | CORO1A | | LINC02345 | MARCKS | CD163 | CSTB | CD63 | | | LSP1 | TIMP1 | SELL | SRGN | | |
| | S100A8 | FOS | | FABP3 | STAB1 | AP2S1 | CTSZ | CD9 | | | ZFP36 | NAMPT | TSPO | LYZ | | |
| | EMP3 | IER2 | | FBP1 | CTSB | MRC1 | FABP5 | PLD3 | | | S100A4 | PLAUR | CSF3R | H3F3B | | |
| | LGMN | CTSB | | SPARC | F13A1 | MARCO | PSAP | LIPA | | | RPL35A | CD300E | MNDA | FPR1 | | |
| | MARCKS | LMNA | | CAMK1 | CTSD | GPX1 | MARCO | LGMN | | | RPS24 | RP11---1143G9.4 | IER2 | NAMPT | | |
| | EMP1 | TYMP | | SDC2 | HSPA5 | CTSZ | CD68 | TREM2 | | | RPS3A | IL1R2 | AP1S2 | COTL1 | | |
| | MS4A6A | CD84 | | A2M | GLUL | OLR1 | GPX1 | MMP9 | | | RPL41 | AIF1 | RBP7 | PLAC8 | | |
| | PLA2G7 | PLEKHO1 | | GPNMB | TGFBI | APOC1 | ACP5 | SDS | | | CCR2 | C5AR1 | CD52 | CFP | | |
| | ZFP36L1 | JUNB | | PLA2G7 | PLA2G7 | GCHFR | MS4A4A | C1QB | | | RPL18 | LST1 | CD36 | ACTB | | |
| | FCGR2B | PMP22 | | MFSD12 | CALR | AGRP | LGALS1 | CCL18 | | | ARHGDIB | PPIF | STXBP2 | IFITM3 | | |
| Proportion of cluster defining genes in specified reference dataset that are also present in Oxford BAL AMs' top 50 cluster defining genes | | | | 0.1 | 0.45 | 0.1 | 0.15 | 0.2 | 0.33 | 0.67 | 0.2 | 0.2 | 0.25 | 0.25 | 0.57 | 0.33 |
