## Supplementary material for "Type I IFN-activated lung monocytes and macrophages as initiators and drivers of fibrosis at the alveolar barrier in IPF": Suppl Table 3B

**Supplementary Table 3B.** Regulon activity in IPF AMs compared to HC (Oxford Cohort A). Statistically significant (p adj <0.01- Bonferroni correction) FC difference ranked by Log2 fold change. PCT1, Percentage of cells with detected regulon score (non -zero) in IPF cell group. pct.1 - pct.1 is the percentage of cells expressing the gene/feature in ident.1 and pct.2 is the percentage of cells that express gene/feature in ident.2.

**Top 10 regulon activity in each AM cluster in IPF and HC**

| gene | avg_log2FC | pct.1 | pct.2 | p_val_adj | cluster |
|---|---|---|---|---|---|
| USF2... | 0.044529577 | 1 | 1 | 4.80E-47 | FABP4hi AM |
| NR1H3... | 0.031640613 | 1 | 1 | 1.25E-39 | FABP4hi AM |
| HES2... | 0.037628674 | 1 | 1 | 8.3156E-31 | FABP4hi AM |
| PPARG... | 0.163120548 | 1 | 0.994 | 6.36E-27 | FABP4hi AM |
| SOX4... | 0.026714312 | 1 | 0.993 | 2.21E-24 | FABP4hi AM |
| DDIT3... | 0.03033194 | 1 | 1 | 4.53E-22 | FABP4hi AM |
| H2AFY... | 0.113352425 | 1 | 1 | 2.41E-15 | FABP4hi AM |
| JDP2... | 0.031823795 | 0.974 | 0.861 | 7.64E-11 | FABP4hi AM |
| CEBPZ... | 0.064383931 | 1 | 0.995 | 1.80E-09 | FABP4hi AM |
| HOXB7... | 0.040207915 | 0.924 | 0.642 | 3.86E-09 | FABP4hi AM |
| MYB... | 0.0522305 | 1 | 1 | 1.93E-49 | IGF1+ AM |
| TCF7L2... | 0.142423186 | 1 | 0.978 | 5.15E-35 | IGF1+ AM |
| NR3C1... | 0.087350156 | 0.968 | 0.707 | 4.72E-30 | IGF1+ AM |
| AR... | 0.031608946 | 0.995 | 0.903 | 1.49E-29 | IGF1+ AM |
| PPARG... | 0.158065699 | 1 | 0.995 | 5.03E-27 | IGF1+ AM |
| RARA... | 0.031744708 | 1 | 1 | 1.04E-22 | IGF1+ AM |
| H2AFY... | 0.100462188 | 1 | 1 | 6.21E-15 | IGF1+ AM |
| HOXB7... | 0.038299091 | 0.922 | 0.662 | 4.34E-11 | IGF1+ AM |
| SMAD7... | 0.038860447 | 0.944 | 0.837 | 1.46E-06 | IGF1+ AM |
| HOXB6... | 0.030292536 | 0.66 | 0.366 | 1.71E-05 | IGF1+ AM |
| SREBF2... | 0.040798198 | 1 | 1 | 4.62E-33 | CD206hi FN1hi AM |
| NFE2... | 0.037402311 | 1 | 1 | 2.16E-29 | CD206hi FN1hi AM |
| TCF7L2... | 0.120684531 | 1 | 0.979 | 8.33E-26 | CD206hi FN1hi AM |
| MYB... | 0.035761735 | 1 | 1 | 2.59E-22 | CD206hi FN1hi AM |
| CEBPA... | 0.028326386 | 1 | 1 | 3.42E-21 | CD206hi FN1hi AM |

| | | | | | |
|---|---|---|---|---|---|
| RARA... | 0.02915534 | 1 | 1 | 7.80E-21 | CD206hi FN1hi AM |
| IKZF2... | 0.027837878 | 0.997 | 0.932 | 2.29E-19 | CD206hi FN1hi AM |
| PPARG... | 0.096417508 | 1 | 0.995 | 6.03E-12 | CD206hi FN1hi AM |
| MYC... | 0.026684371 | 1 | 1 | 7.57E-10 | CD206hi FN1hi AM |
| H2AFY... | 0.065961643 | 1 | 1 | 1.36E-06 | CD206hi FN1hi AM |
| BCL3... | 0.102502405 | 1 | 0.948 | 1.10E-51 | CXCL10+ AM |
| HIVEP1... | 0.055255074 | 1 | 1 | 7.55E-40 | CXCL10+ AM |
| ATF5... | 0.041825837 | 1 | 0.996 | 1.19E-32 | CXCL10+ AM |
| BCL6... | 0.046142286 | 0.998 | 0.904 | 4.63E-28 | CXCL10+ AM |
| NFKB2... | 0.040900987 | 1 | 1 | 3.73E-25 | CXCL10+ AM |
| ETV7... | 0.047153211 | 1 | 1 | 3.97E-25 | CXCL10+ AM |
| ZNF267... | 0.07716277 | 0.847 | 0.484 | 9.12E-14 | CXCL10+ AM |
| H2AFY... | 0.095290927 | 1 | 1 | 2.56E-13 | CXCL10+ AM |
| HIF1A... | 0.044456859 | 0.819 | 0.558 | 1.11E-06 | CXCL10+ AM |
| PPARG... | 0.070356803 | 1 | 0.996 | 4.17E-06 | CXCL10+ AM |
| MEF2C... | 0.04590784 | 1 | 1 | 7.31E-52 | mono-SPP1+ AM |
| ATF5... | 0.051338106 | 1 | 0.995 | 1.10E-47 | mono-SPP1+ AM |
| MYC... | 0.078801189 | 1 | 1 | 5.13E-46 | mono-SPP1+ AM |
| MAFG... | 0.050983174 | 1 | 0.984 | 1.47E-44 | mono-SPP1+ AM |
| FOSL2... | 0.045723329 | 1 | 1 | 2.03E-39 | mono-SPP1+ AM |
| ARID3A... | 0.047149989 | 0.997 | 0.963 | 1.69E-28 | mono-SPP1+ AM |
| MEF2A... | 0.038759908 | 1 | 0.999 | 1.06E-23 | mono-SPP1+ AM |
| EGR1... | 0.057714391 | 0.7 | 0.383 | 4.98E-09 | mono-SPP1+ AM |
| HIF1A... | 0.073203239 | 0.723 | 0.544 | 5.80E-08 | mono-SPP1+ AM |
| SMAD7... | 0.042139147 | 0.932 | 0.848 | 3.54E-05 | mono-SPP1+ AM |
| E2F1... | 0.025327743 | 1 | 1 | 5.06E-56 | Cycling AM1 |
| BRCA1... | 0.021960682 | 1 | 1 | 5.55E-43 | Cycling AM1 |
| POLE4... | 0.033100463 | 1 | 1 | 4.84E-42 | Cycling AM1 |
| POLE3... | 0.026536748 | 1 | 1 | 5.84E-41 | Cycling AM1 |
| MYBL2... | 0.022990485 | 1 | 1 | 6.62E-38 | Cycling AM1 |
| TCF7L2... | 0.067097821 | 0.995 | 0.982 | 7.41E-09 | Cycling AM1 |
| H2AFY... | 0.070043929 | 1 | 1 | 1.14E-08 | Cycling AM1 |

| | | | | | |
|---|---|---|---|---|---|
| PPARG... | 0.078512558 | 0.998 | 0.996 | 5.07E-08 | Cycling AM1 |
| CEBPZ... | 0.025384102 | 1 | 0.996 | 2.19E-06 | Cycling AM1 |
| SMAD7... | 0.033153505 | 0.945 | 0.855 | 0.000127277 | Cycling AM1 |
| FOXM1... | 0.318173001 | 0.994 | 0.292 | 5.67E-103 | Cycling AM2 |
| MYBL2... | 0.076049057 | 1 | 1 | 6.43E-100 | Cycling AM2 |
| HMGB3... | 0.119122494 | 1 | 0.999 | 6.41E-95 | Cycling AM2 |
| E2F7... | 0.06836095 | 1 | 1 | 3.02E-94 | Cycling AM2 |
| EZH2... | 0.070999168 | 1 | 1 | 4.06E-93 | Cycling AM2 |
| MAZ... | 0.084464914 | 1 | 1 | 2.10E-92 | Cycling AM2 |
| E2F2... | 0.064781268 | 1 | 1 | 1.11E-83 | Cycling AM2 |
| NFYC... | 0.103132782 | 1 | 0.988 | 4.26E-71 | Cycling AM2 |
| CTCF... | 0.066341993 | 1 | 1 | 6.92E-62 | Cycling AM2 |
| YEATS4... | 0.105390575 | 0.611 | 0.053 | 1.09E-37 | Cycling AM2 |
