## Supplementary material for "Type I IFN-activated lung monocytes and macrophages as initiators and drivers of fibrosis at the alveolar barrier in IPF": Suppl Table 4A

**Supplementary Table 4A.** Inter-cluster DEGs for Morse lung digest myeloid cells (sub-clustered and annotated by Oxford). Pct.1 – proportion of cells in selected cluster expressing gene. Pct .2 proportion of cells expressing gene in all other remaining clusters.

| gene | avg_log2FC | pct.1 | pct.2 | p_val_adj | cluster |
|---|---|---|---|---|---|
| FABP4 | 2.59406552 | 0.984 | 0.31 | 3.38E-48 | FABP4hi AM |
| SERPING1 | 1.746397 | 0.893 | 0.215 | 2.41E-37 | FABP4hi AM |
| INHBA | 1.70547389 | 0.906 | 0.199 | 4.88E-40 | FABP4hi AM |
| C1QB | 1.57721386 | 0.998 | 0.701 | 1.88E-34 | FABP4hi AM |
| CD52 | 1.51259793 | 0.999 | 0.774 | 3.49E-37 | FABP4hi AM |
| RP11-598F7. | 1.49116848 | 0.915 | 0.212 | 6.35E-41 | FABP4hi AM |
| AKR1C3 | 1.4629071 | 0.79 | 0.196 | 3.08E-20 | FABP4hi AM |
| RND3 | 1.4465385 | 0.645 | 0.097 | 1.23E-21 | FABP4hi AM |
| C1QA | 1.44532011 | 0.999 | 0.732 | 4.12E-32 | FABP4hi AM |
| SERPINA1 | 1.36423193 | 0.991 | 0.62 | 4.67E-32 | FABP4hi AM |
| RBP4 | 1.33785496 | 0.716 | 0.15 | 1.32E-28 | FABP4hi AM |
| IFI27 | 1.32830264 | 0.386 | 0.173 | 0.22968429 | FABP4hi AM |
| FBP1 | 1.31202966 | 0.994 | 0.676 | 6.98E-26 | FABP4hi AM |
| CES1 | 1.24653892 | 0.726 | 0.228 | 3.84E-15 | FABP4hi AM |
| LY6E | 1.24190918 | 0.983 | 0.544 | 5.41E-30 | FABP4hi AM |
| LGALS3BP | 1.23207759 | 0.834 | 0.188 | 2.69E-29 | FABP4hi AM |
| MCEMP1 | 1.22548485 | 0.984 | 0.521 | 4.93E-29 | FABP4hi AM |
| PDLIM1 | 1.18628378 | 0.905 | 0.238 | 1.02E-33 | FABP4hi AM |
| PHLDA3 | 1.18366279 | 0.905 | 0.206 | 8.33E-35 | FABP4hi AM |
| FABP3 | 1.17945301 | 0.519 | 0.231 | 1.40E-09 | FABP4hi AM |
| C1QC | 1.17306371 | 0.99 | 0.62 | 1.51E-19 | FABP4hi AM |
| PLA2G16 | 1.16238312 | 0.954 | 0.313 | 1.34E-37 | FABP4hi AM |
| PCOLCE2 | 1.15139635 | 0.824 | 0.17 | 4.59E-29 | FABP4hi AM |
| ALOX5AP | 1.14663155 | 0.999 | 0.754 | 3.85E-17 | FABP4hi AM |
| ALDH2 | 1.13294572 | 0.982 | 0.624 | 7.64E-29 | FABP4hi AM |
| FHL1 | 1.12748344 | 0.828 | 0.147 | 1.01E-35 | FABP4hi AM |
| AQP3 | 1.10956446 | 0.836 | 0.178 | 8.23E-29 | FABP4hi AM |
| UBB | 1.09291623 | 0.998 | 0.855 | 1.21E-27 | FABP4hi AM |
| RP5-839B4.8 | 1.04776021 | 0.819 | 0.123 | 3.03E-32 | FABP4hi AM |
| TREM1 | 0.98076908 | 0.975 | 0.544 | 3.48E-23 | FABP4hi AM |
| LDHB | 0.97618467 | 0.934 | 0.507 | 4.32E-22 | FABP4hi AM |
| PPIC | 0.97392664 | 0.793 | 0.142 | 4.05E-36 | FABP4hi AM |
| MT1E | 0.97087993 | 0.447 | 0.22 | 0.06492712 | FABP4hi AM |
| HPGD | 0.9279588 | 0.868 | 0.274 | 2.03E-27 | FABP4hi AM |
| GPD1 | 0.9014093 | 0.668 | 0.046 | 4.92E-29 | FABP4hi AM |
| FAM89A | 0.87989284 | 0.827 | 0.199 | 7.40E-27 | FABP4hi AM |
| GLDN | 0.86754406 | 0.771 | 0.19 | 1.45E-18 | FABP4hi AM |
| MIR3945HG | 0.86079237 | 0.788 | 0.32 | 6.19E-06 | FABP4hi AM |
| CITED2 | 0.85661934 | 0.909 | 0.491 | 3.18E-14 | FABP4hi AM |
| S100A13 | 0.84947888 | 0.849 | 0.307 | 2.79E-21 | FABP4hi AM |

| | | | | | |
|---|---|---|---|---|---|
| STOM | 0.81701132 | 0.907 | 0.423 | 1.86E-20 | FABP4hi AM |
| PEBP1 | 0.80710562 | 0.98 | 0.634 | 5.55E-20 | FABP4hi AM |
| LPL | 0.80217068 | 0.91 | 0.372 | 7.41E-16 | FABP4hi AM |
| AKR1C2 | 0.79984897 | 0.4 | 0.038 | 2.30E-11 | FABP4hi AM |
| EVL | 0.79721597 | 0.782 | 0.254 | 2.77E-23 | FABP4hi AM |
| CTSC | 0.7954527 | 0.99 | 0.739 | 5.74E-15 | FABP4hi AM |
| ACOT7 | 0.79243798 | 0.733 | 0.192 | 9.98E-28 | FABP4hi AM |
| ALOX5 | 0.79006016 | 0.93 | 0.471 | 2.32E-15 | FABP4hi AM |
| DOK2 | 0.7861462 | 0.907 | 0.402 | 3.18E-15 | FABP4hi AM |
| PPARG | 0.78585689 | 0.908 | 0.38 | 1.41E-22 | FABP4hi AM |

| **gene** | **avg_log2FC** | **pct.1** | **pct.2** | **p_val_adj** | **cluster** |
|---|---|---|---|---|---|
| MARCO | 1.02444504 | 0.946 | 0.743 | 7.80E-08 | Intermediate AM_1 |
| RETN | 0.86023335 | 0.637 | 0.442 | 1 | Intermediate AM_1 |
| ACP5 | 0.85213316 | 0.987 | 0.798 | 8.05E-08 | Intermediate AM_1 |
| VSIG4 | 0.79815494 | 0.903 | 0.74 | 4.36E-08 | Intermediate AM_1 |
| CCL20 | 0.73205445 | 0.553 | 0.412 | 0.25043588 | Intermediate AM_1 |
| CYP27A1 | 0.69323117 | 0.874 | 0.595 | 4.38E-07 | Intermediate AM_1 |
| GRN | 0.68696022 | 0.987 | 0.889 | 5.84E-11 | Intermediate AM_1 |
| AGRP | 0.68250882 | 0.404 | 0.234 | 0.03470631 | Intermediate AM_1 |
| APOE | 0.62787266 | 0.869 | 0.731 | 0.1372915 | Intermediate AM_1 |
| LTA4H | 0.59663714 | 0.868 | 0.67 | 0.00248801 | Intermediate AM_1 |
| ANXA1 | 0.56880857 | 0.985 | 0.895 | 0.27147047 | Intermediate AM_1 |
| CD68 | 0.56644387 | 0.992 | 0.881 | 1.81E-07 | Intermediate AM_1 |
| HEXB | 0.55680329 | 0.956 | 0.738 | 1.15E-05 | Intermediate AM_1 |
| LRPAP1 | 0.55359976 | 0.89 | 0.661 | 2.48E-05 | Intermediate AM_1 |
| LGALS3 | 0.49640993 | 0.996 | 0.921 | 0.14113334 | Intermediate AM_1 |
| FBP1 | 0.48991947 | 0.919 | 0.73 | 0.54080636 | Intermediate AM_1 |
| CD9 | 0.48725444 | 0.911 | 0.724 | 1 | Intermediate AM_1 |
| HLA-A | 0.47491699 | 0.99 | 0.938 | 0.10568313 | Intermediate AM_1 |
| CTSA | 0.45761258 | 0.798 | 0.578 | 0.00040266 | Intermediate AM_1 |
| PLIN2 | 0.45553761 | 0.979 | 0.855 | 1 | Intermediate AM_1 |
| FCGRT | 0.44538233 | 0.966 | 0.854 | 5.94E-05 | Intermediate AM_1 |
| QPCT | 0.44215645 | 0.504 | 0.243 | 9.15E-06 | Intermediate AM_1 |
| CXCL3 | 0.43823409 | 0.842 | 0.74 | 1 | Intermediate AM_1 |
| CTSD | 0.42138388 | 0.997 | 0.933 | 4.27E-05 | Intermediate AM_1 |
| IL3RA | 0.41630849 | 0.453 | 0.255 | 1 | Intermediate AM_1 |
| HLA-C | 0.41057412 | 0.991 | 0.939 | 1 | Intermediate AM_1 |
| MGST3 | 0.40565704 | 0.98 | 0.819 | 0.00516062 | Intermediate AM_1 |
| MCEMP1 | 0.39974373 | 0.833 | 0.605 | 1 | Intermediate AM_1 |
| ALOX5AP | 0.39923493 | 0.899 | 0.801 | 0.26385332 | Intermediate AM_1 |
| CD164 | 0.39733052 | 0.927 | 0.77 | 0.67280224 | Intermediate AM_1 |
| SLC3A2 | 0.39659106 | 0.677 | 0.48 | 1 | Intermediate AM_1 |
| CD14 | 0.39654086 | 0.693 | 0.515 | 1 | Intermediate AM_1 |
| S100P | 0.39363365 | 0.225 | 0.096 | 0.50525573 | Intermediate AM_1 |
| RNH1 | 0.39173321 | 0.893 | 0.717 | 1 | Intermediate AM_1 |
| MYDGF | 0.38641613 | 0.881 | 0.703 | 0.0581426 | Intermediate AM_1 |
| S100A11 | 0.38268343 | 0.999 | 0.99 | 0.00050179 | Intermediate AM_1 |

| | | | | | |
|---|---|---|---|---|---|
| HEXA | 0.38080544 | 0.697 | 0.503 | 0.00025068 | Intermediate AM_1 |
| SLC31A2 | 0.37128235 | 0.826 | 0.638 | 0.04138047 | Intermediate AM_1 |
| ARL4A | 0.37034612 | 0.877 | 0.705 | 0.10735213 | Intermediate AM_1 |
| ANXA2 | 0.36404321 | 0.994 | 0.935 | 1 | Intermediate AM_1 |
| ATP6V0D1 | 0.3551026 | 0.862 | 0.68 | 1 | Intermediate AM_1 |
| CD63 | 0.34969982 | 0.998 | 0.97 | 1 | Intermediate AM_1 |
| HLA-B | 0.34921944 | 0.999 | 0.981 | 0.74746454 | Intermediate AM_1 |
| PPARG | 0.34883742 | 0.697 | 0.48 | 1 | Intermediate AM_1 |
| FXYD5 | 0.34507801 | 0.893 | 0.743 | 1 | Intermediate AM_1 |
| ABHD5 | 0.34388453 | 0.659 | 0.451 | 0.24108727 | Intermediate AM_1 |
| GSTO1 | 0.34235408 | 0.981 | 0.875 | 1 | Intermediate AM_1 |
| EMP3 | 0.34120248 | 0.976 | 0.906 | 1 | Intermediate AM_1 |
| ALDH2 | 0.33963481 | 0.861 | 0.689 | 1 | Intermediate AM_1 |
| SLC11A1 | 0.33581661 | 0.925 | 0.772 | 1 | Intermediate AM_1 |
| NUPR1 | 1.19324007 | 0.84 | 0.511 | 3.58E-10 | Intermediate AM_2 |
| CCL18 | 1.15748766 | 0.867 | 0.634 | 0.00707979 | Intermediate AM_2 |
| IFI27 | 1.07251678 | 0.378 | 0.212 | 1 | Intermediate AM_2 |
| FABP4 | 1.05678802 | 0.77 | 0.445 | 9.17E-05 | Intermediate AM_2 |
| RBP4 | 0.99382649 | 0.565 | 0.261 | 8.10E-07 | Intermediate AM_2 |
| SCD | 0.99210794 | 0.669 | 0.425 | 2.84E-06 | Intermediate AM_2 |
| GCHFR | 0.90089413 | 0.961 | 0.714 | 8.38E-15 | Intermediate AM_2 |
| RP11-598F7. | 0.83033083 | 0.676 | 0.354 | 1.46E-05 | Intermediate AM_2 |
| APOC1 | 0.81241417 | 0.996 | 0.813 | 5.19E-10 | Intermediate AM_2 |
| FTL | 0.69901559 | 1 | 1 | 1.13E-14 | Intermediate AM_2 |
| CES1 | 0.63751714 | 0.577 | 0.327 | 0.00977799 | Intermediate AM_2 |
| CYB5A | 0.61046576 | 0.832 | 0.631 | 4.653189340 | Intermediate AM_2 |
| LGALS3 | 0.59265393 | 0.99 | 0.925 | 0.00709065 | Intermediate AM_2 |
| MSR1 | 0.5722396 | 0.854 | 0.669 | 0.34176324 | Intermediate AM_2 |
| CSTB | 0.51772769 | 0.999 | 0.951 | 0.0070155 | Intermediate AM_2 |
| APOE | 0.49698488 | 0.931 | 0.732 | 1 | Intermediate AM_2 |
| RND3 | 0.48778835 | 0.368 | 0.213 | 1 | Intermediate AM_2 |
| PLA2G16 | 0.46761876 | 0.698 | 0.445 | 0.74354152 | Intermediate AM_2 |
| TXN | 0.4605042 | 0.976 | 0.921 | 0.55801111 | Intermediate AM_2 |
| PPDPF | 0.45951742 | 0.89 | 0.793 | 0.00146043 | Intermediate AM_2 |
| PDLIM1 | 0.45410691 | 0.605 | 0.378 | 0.00943813 | Intermediate AM_2 |
| SERPING1 | 0.42998982 | 0.587 | 0.357 | 1 | Intermediate AM_2 |
| ALDH2 | 0.42791997 | 0.88 | 0.695 | 0.62750697 | Intermediate AM_2 |
| PHLDA3 | 0.42727122 | 0.59 | 0.352 | 0.00715106 | Intermediate AM_2 |
| RMDN3 | 0.42637638 | 0.675 | 0.472 | 0.0010438 | Intermediate AM_2 |
| C1QB | 0.42591195 | 0.986 | 0.755 | 0.06508546 | Intermediate AM_2 |
| ACOT7 | 0.42571501 | 0.466 | 0.306 | 1 | Intermediate AM_2 |
| AKR1C3 | 0.40728889 | 0.548 | 0.318 | 1 | Intermediate AM_2 |
| HPGD | 0.40664212 | 0.594 | 0.399 | 0.04012545 | Intermediate AM_2 |
| GLRX | 0.40483533 | 0.922 | 0.808 | 0.49704243 | Intermediate AM_2 |
| UBB | 0.39719548 | 0.975 | 0.882 | 0.14451323 | Intermediate AM_2 |
| AKR1B1 | 0.39374856 | 0.73 | 0.558 | 0.01359432 | Intermediate AM_2 |
| ADTRP | 0.39233869 | 0.366 | 0.197 | 1 | Intermediate AM_2 |

| | | | | | |
|---|---|---|---|---|---|
| SCCPDH | 0.38993954 | 0.644 | 0.495 | 1 | Intermediate AM_2 |
| FTH1 | 0.38434778 | 1 | 1 | 0.00010614 | Intermediate AM_2 |
| BLOC1S2 | 0.38224617 | 0.748 | 0.603 | 1 | Intermediate AM_2 |
| ABCG1 | 0.36580209 | 0.602 | 0.428 | 0.03166253 | Intermediate AM_2 |
| TCEB2 | 0.35851499 | 0.957 | 0.918 | 0.02777815 | Intermediate AM_2 |
| LDHB | 0.35684143 | 0.751 | 0.596 | 1 | Intermediate AM_2 |
| S100A13 | 0.34964973 | 0.615 | 0.419 | 0.00358953 | Intermediate AM_2 |
| FAM89A | 0.34790767 | 0.531 | 0.331 | 1 | Intermediate AM_2 |
| PPIC | 0.34410147 | 0.439 | 0.283 | 1 | Intermediate AM_2 |
| PCOLCE2 | 0.33718809 | 0.506 | 0.308 | 0.59488141 | Intermediate AM_2 |
| C9orf16 | 0.32628029 | 0.836 | 0.747 | 1 | Intermediate AM_2 |
| GLUL | 0.32121371 | 0.983 | 0.949 | 1 | Intermediate AM_2 |
| CYP27A1 | 0.31111741 | 0.794 | 0.611 | 1 | Intermediate AM_2 |
| DNAJC5B | 0.30794276 | 0.261 | 0.152 | 1 | Intermediate AM_2 |
| MME | 0.3046479 | 0.347 | 0.207 | 1 | Intermediate AM_2 |
| PTPMT1 | 0.3020236 | 0.624 | 0.47 | 1 | Intermediate AM_2 |
| C1QA | 0.3000521 | 0.989 | 0.781 | 1 | Intermediate AM_2 |
| ISG15 | 3.04886678 | 1 | 0.604 | 6.14E-55 | CXCL10+ Macs |
| CXCL10 | 2.34908535 | 0.332 | 0.04 | 3.95E-10 | CXCL10+ Macs |
| IFIT1 | 2.0101065 | 0.818 | 0.094 | 1.71E-45 | CXCL10+ Macs |
| IFIT3 | 1.80837394 | 0.825 | 0.212 | 4.24E-40 | CXCL10+ Macs |
| IFIT2 | 1.66158408 | 0.682 | 0.123 | 1.45E-24 | CXCL10+ Macs |
| MX1 | 1.6302152 | 0.864 | 0.248 | 9.28E-39 | CXCL10+ Macs |
| NT5C3A | 1.50307474 | 0.711 | 0.225 | 1.87E-23 | CXCL10+ Macs |
| IFI6 | 1.41406044 | 0.968 | 0.686 | 7.89E-25 | CXCL10+ Macs |
| TNFSF10 | 1.20517924 | 0.593 | 0.094 | 4.42E-25 | CXCL10+ Macs |
| IFITM3 | 1.1857664 | 0.929 | 0.588 | 2.71E-21 | CXCL10+ Macs |
| LY6E | 1.16130041 | 0.954 | 0.641 | 6.67E-21 | CXCL10+ Macs |
| RSAD2 | 1.160833 | 0.571 | 0.034 | 7.13E-23 | CXCL10+ Macs |
| EPSTI1 | 1.1588521 | 0.779 | 0.292 | 1.07E-26 | CXCL10+ Macs |
| TNFSF13B | 1.11131887 | 0.9 | 0.478 | 3.50E-23 | CXCL10+ Macs |
| OAS1 | 1.1100375 | 0.771 | 0.286 | 1.26E-22 | CXCL10+ Macs |
| IFI44L | 1.06214219 | 0.7 | 0.141 | 8.71E-28 | CXCL10+ Macs |
| CMPK2 | 0.98551422 | 0.643 | 0.09 | 2.99E-25 | CXCL10+ Macs |
| SAMD9L | 0.98470357 | 0.75 | 0.249 | 5.52E-25 | CXCL10+ Macs |
| UBE2L6 | 0.90947496 | 0.9 | 0.555 | 2.36E-14 | CXCL10+ Macs |
| NUPR1 | 0.89141444 | 0.696 | 0.53 | 1 | CXCL10+ Macs |
| OASL | 0.87699846 | 0.657 | 0.258 | 4.47E-13 | CXCL10+ Macs |
| OAS2 | 0.87594825 | 0.7 | 0.18 | 6.91E-19 | CXCL10+ Macs |
| HERC5 | 0.87159699 | 0.586 | 0.107 | 6.08E-23 | CXCL10+ Macs |
| USP18 | 0.86680524 | 0.561 | 0.072 | 1.38E-17 | CXCL10+ Macs |
| GBP1 | 0.86394177 | 0.625 | 0.224 | 6.71E-12 | CXCL10+ Macs |
| CCL8 | 0.86285097 | 0.121 | 0.014 | 0.04381713 | CXCL10+ Macs |
| EIF2AK2 | 0.8626101 | 0.746 | 0.3 | 5.65E-21 | CXCL10+ Macs |
| IFI35 | 0.85459882 | 0.707 | 0.291 | 1.17E-15 | CXCL10+ Macs |
| LAP3 | 0.84837489 | 0.868 | 0.568 | 2.28E-17 | CXCL10+ Macs |
| DYNLT1 | 0.8453949 | 0.918 | 0.68 | 2.17E-17 | CXCL10+ Macs |

| | | | | | |
|---|---|---|---|---|---|
| IFI44 | 0.8343502 | 0.65 | 0.212 | 2.47E-15 | CXCL10+ Macs |
| CCL2 | 0.83154967 | 0.354 | 0.236 | 0.67829685 | CXCL10+ Macs |
| STAT1 | 0.8170886 | 0.725 | 0.341 | 1.05E-14 | CXCL10+ Macs |
| PSMB9 | 0.81003783 | 0.85 | 0.523 | 2.61E-14 | CXCL10+ Macs |
| RABGAP1L | 0.79859635 | 0.546 | 0.143 | 7.11E-14 | CXCL10+ Macs |
| MNDA | 0.79195076 | 0.868 | 0.598 | 1.48E-09 | CXCL10+ Macs |
| DEFB1 | 0.75655541 | 0.396 | 0.158 | 0.00880585 | CXCL10+ Macs |
| NEXN | 0.75490169 | 0.446 | 0.044 | 1.26E-15 | CXCL10+ Macs |
| OAS3 | 0.75374559 | 0.575 | 0.144 | 1.82E-19 | CXCL10+ Macs |
| IFI27 | 0.74560293 | 0.343 | 0.221 | 1 | CXCL10+ Macs |
| RNF213 | 0.73289958 | 0.746 | 0.399 | 5.22E-17 | CXCL10+ Macs |
| IRF7 | 0.73021185 | 0.675 | 0.278 | 5.53E-08 | CXCL10+ Macs |
| ISG20 | 0.72944468 | 0.45 | 0.125 | 1.73E-09 | CXCL10+ Macs |
| PLSCR1 | 0.70564956 | 0.882 | 0.616 | 1.86E-11 | CXCL10+ Macs |
| MX2 | 0.70359272 | 0.518 | 0.134 | 6.81E-16 | CXCL10+ Macs |
| CCL7 | 0.69790553 | 0.143 | 0.073 | 1 | CXCL10+ Macs |
| XAF1 | 0.68330549 | 0.543 | 0.155 | 2.05E-15 | CXCL10+ Macs |
| PARP14 | 0.67894675 | 0.661 | 0.322 | 3.70E-11 | CXCL10+ Macs |
| GMPR | 0.67225012 | 0.518 | 0.133 | 2.17E-09 | CXCL10+ Macs |
| BST2 | 0.6644064 | 0.875 | 0.603 | 6.24E-07 | CXCL10+ Macs |
| LGMN | 1.73034304 | 0.853 | 0.357 | 9.97E-26 | SPP1_2 |
| APOE | 1.56978978 | 0.931 | 0.714 | 2.96E-14 | SPP1_2 |
| CCL18 | 1.40762462 | 0.87 | 0.613 | 5.57E-11 | SPP1_2 |
| GPNMB | 1.2768442 | 0.972 | 0.737 | 5.13E-28 | SPP1_2 |
| CTSZ | 1.11861088 | 0.943 | 0.746 | 2.15E-14 | SPP1_2 |
| RNASE1 | 1.11820858 | 0.609 | 0.242 | 7.16E-09 | SPP1_2 |
| CTSB | 1.0710382 | 0.998 | 0.946 | 8.86E-26 | SPP1_2 |
| PLA2G7 | 0.96791795 | 0.721 | 0.295 | 6.55E-13 | SPP1_2 |
| SPP1 | 0.92836312 | 0.54 | 0.226 | 6.38E-05 | SPP1_2 |
| HMOX1 | 0.88779301 | 0.55 | 0.399 | 1 | SPP1_2 |
| CTSL | 0.88116469 | 0.966 | 0.842 | 2.24E-07 | SPP1_2 |
| CCL13 | 0.86685676 | 0.26 | 0.07 | 1 | SPP1_2 |
| SEPP1 | 0.86625391 | 0.356 | 0.093 | 4.75E-08 | SPP1_2 |
| CPM | 0.8168766 | 0.78 | 0.507 | 7.81E-13 | SPP1_2 |
| CTSD | 0.80016592 | 0.999 | 0.93 | 4.38E-13 | SPP1_2 |
| APOC1 | 0.79252578 | 0.973 | 0.801 | 5.95E-05 | SPP1_2 |
| LIPA | 0.7876798 | 0.815 | 0.625 | 4.09E-11 | SPP1_2 |
| CCL2 | 0.78743895 | 0.388 | 0.213 | 1 | SPP1_2 |
| MARCKS | 0.77328684 | 0.663 | 0.233 | 2.38E-09 | SPP1_2 |
| PSAP | 0.76956826 | 0.998 | 0.966 | 1.17E-20 | SPP1_2 |
| HS3ST2 | 0.72421693 | 0.263 | 0.041 | 6.52E-05 | SPP1_2 |
| PLTP | 0.7080746 | 0.449 | 0.142 | 4.01E-11 | SPP1_2 |
| CD84 | 0.70120313 | 0.608 | 0.245 | 5.54E-07 | SPP1_2 |
| PLD3 | 0.69777083 | 0.753 | 0.529 | 1.77E-08 | SPP1_2 |
| TREM2 | 0.67744394 | 0.694 | 0.496 | 0.00154558 | SPP1_2 |
| PMP22 | 0.65615756 | 0.75 | 0.416 | 7.82E-10 | SPP1_2 |
| NPC2 | 0.6557772 | 0.997 | 0.975 | 1.37E-20 | SPP1_2 |

| | | | | | |
|---|---|---|---|---|---|
| DAB2 | 0.6486678 | 0.884 | 0.665 | 9.79E-10 | SPP1_2 |
| LGALS1 | 0.63003388 | 0.997 | 0.98 | 1.19E-08 | SPP1_2 |
| RGS1 | 0.61168311 | 0.735 | 0.39 | 1.22E-12 | SPP1_2 |
| FTL | 0.60476838 | 1 | 1 | 3.80E-09 | SPP1_2 |
| CD59 | 0.60335262 | 0.84 | 0.63 | 0.0001423 | SPP1_2 |
| TGFBI | 0.5967414 | 0.7 | 0.537 | 0.23324897 | SPP1_2 |
| SGK1 | 0.59058136 | 0.809 | 0.556 | 7.21E-05 | SPP1_2 |
| ARL4C | 0.58645095 | 0.517 | 0.216 | 5.71E-06 | SPP1_2 |
| CREG1 | 0.58179398 | 0.872 | 0.686 | 0.00191775 | SPP1_2 |
| NPL | 0.5801991 | 0.683 | 0.388 | 4.28E-12 | SPP1_2 |
| ABCA1 | 0.5592859 | 0.549 | 0.221 | 2.63E-07 | SPP1_2 |
| CD36 | 0.55353246 | 0.535 | 0.286 | 2.68E-05 | SPP1_2 |
| MS4A4A | 0.55143936 | 0.879 | 0.691 | 0.17553142 | SPP1_2 |
| TSPAN4 | 0.54326094 | 0.569 | 0.303 | 7.19E-09 | SPP1_2 |
| SDS | 0.5432143 | 0.365 | 0.116 | 3.38E-05 | SPP1_2 |
| TMIGD3 | 0.53611063 | 0.261 | 0.101 | 0.43485021 | SPP1_2 |
| FPR3 | 0.53194613 | 0.642 | 0.331 | 0.00423274 | SPP1_2 |
| FOSB | 0.52556844 | 0.816 | 0.6 | 0.00252088 | SPP1_2 |
| ZFAND5 | 0.52416924 | 0.904 | 0.783 | 3.57E-05 | SPP1_2 |
| SMPDL3A | 0.51879892 | 0.399 | 0.124 | 5.17E-12 | SPP1_2 |
| FOS | 0.51364221 | 0.876 | 0.755 | 0.64696864 | SPP1_2 |
| MGLL | 0.51181978 | 0.512 | 0.257 | 0.00018903 | SPP1_2 |
| HAMP | 0.51087095 | 0.234 | 0.05 | 1.61E-05 | SPP1_2 |
| CHI3L1 | 3.30100691 | 0.674 | 0.04 | 1.48E-36 | SPP1 Mac |
| MMP7 | 2.89162516 | 0.367 | 0.03 | 4.20E-14 | SPP1 Mac |
| SPP1 | 2.61851982 | 0.868 | 0.251 | 4.26E-37 | SPP1 Mac |
| CHIT1 | 2.40267463 | 0.575 | 0.049 | 8.67E-31 | SPP1 Mac |
| CTSK | 2.09047938 | 0.481 | 0.088 | 5.42E-17 | SPP1 Mac |
| MMP9 | 2.01506801 | 0.491 | 0.06 | 9.23E-19 | SPP1 Mac |
| SDC2 | 1.74500854 | 0.916 | 0.361 | 9.01E-36 | SPP1 Mac |
| TIMP3 | 1.44910046 | 0.364 | 0.054 | 2.17E-10 | SPP1 Mac |
| LIPA | 1.44223506 | 0.954 | 0.642 | 4.37E-30 | SPP1 Mac |
| IGLC2 | 1.33166404 | 0.171 | 0.061 | 0.00659165 | SPP1 Mac |
| FDX1 | 1.31062486 | 0.928 | 0.565 | 6.03E-23 | SPP1 Mac |
| CALM3 | 1.29483945 | 0.954 | 0.707 | 3.09E-24 | SPP1 Mac |
| GPNMB | 1.27647476 | 0.99 | 0.763 | 7.90E-24 | SPP1 Mac |
| APOE | 1.26833192 | 0.966 | 0.738 | 1.71E-18 | SPP1 Mac |
| LILRB4 | 1.24277045 | 0.907 | 0.399 | 2.28E-35 | SPP1 Mac |
| MATK | 1.2148827 | 0.769 | 0.116 | 1.85E-29 | SPP1 Mac |
| TREM2 | 1.195236 | 0.933 | 0.511 | 1.66E-26 | SPP1 Mac |
| CIR1 | 1.18899517 | 0.656 | 0.324 | 1.69E-12 | SPP1 Mac |
| PLA2G7 | 1.18179736 | 0.877 | 0.339 | 6.04E-24 | SPP1 Mac |
| RARRES1 | 1.17205557 | 0.551 | 0.096 | 3.39E-25 | SPP1 Mac |
| SPARC | 1.16218623 | 0.686 | 0.206 | 2.00E-26 | SPP1 Mac |
| FABP5 | 1.15545189 | 0.997 | 0.848 | 1.39E-16 | SPP1 Mac |
| CD9 | 1.12992871 | 0.991 | 0.735 | 2.48E-18 | SPP1 Mac |
| GM2A | 1.0598098 | 0.882 | 0.459 | 1.91E-16 | SPP1 Mac |

| | | | | | |
|---|---|---|---|---|---|
| TM4SF19 | 1.05148073 | 0.555 | 0.108 | 3.97E-15 | SPP1 Mac |
| LPL | 1.04556214 | 0.9 | 0.483 | 1.89E-11 | SPP1 Mac |
| PLD3 | 1.01304625 | 0.898 | 0.549 | 6.07E-20 | SPP1 Mac |
| RP11-20G13. | 0.99147265 | 0.473 | 0.028 | 1.18E-21 | SPP1 Mac |
| CSTB | 0.97060486 | 1 | 0.953 | 2.95E-26 | SPP1 Mac |
| ARID5B | 0.96233521 | 0.88 | 0.331 | 3.06E-23 | SPP1 Mac |
| LGMN | 0.9427475 | 0.775 | 0.416 | 2.08E-11 | SPP1 Mac |
| GSN | 0.93905231 | 0.948 | 0.642 | 8.46E-15 | SPP1 Mac |
| RGCC | 0.92396237 | 0.993 | 0.855 | 7.50E-07 | SPP1 Mac |
| CAPG | 0.90112381 | 0.987 | 0.835 | 3.83E-17 | SPP1 Mac |
| DNASE2B | 0.86846371 | 0.537 | 0.158 | 5.83E-15 | SPP1 Mac |
| MFSD12 | 0.85587618 | 0.754 | 0.292 | 1.49E-13 | SPP1 Mac |
| HCST | 0.85352719 | 0.954 | 0.639 | 2.30E-16 | SPP1 Mac |
| ITGB2 | 0.84125547 | 0.948 | 0.693 | 6.16E-17 | SPP1 Mac |
| CYP27A1 | 0.82908133 | 0.937 | 0.613 | 1.57E-12 | SPP1 Mac |
| FNIP2 | 0.82093271 | 0.874 | 0.422 | 2.03E-16 | SPP1 Mac |
| APOC1 | 0.81973826 | 0.993 | 0.82 | 3.86E-10 | SPP1 Mac |
| ITGB1BP1 | 0.80949579 | 0.87 | 0.453 | 6.88E-20 | SPP1 Mac |
| BCAP31 | 0.80760042 | 0.976 | 0.718 | 4.59E-19 | SPP1 Mac |
| CD63 | 0.80423588 | 1 | 0.972 | 1.73E-17 | SPP1 Mac |
| GPC4 | 0.7978232 | 0.451 | 0.026 | 3.39E-23 | SPP1 Mac |
| MGLL | 0.7933266 | 0.716 | 0.279 | 4.08E-15 | SPP1 Mac |
| CD36 | 0.79274763 | 0.59 | 0.312 | 3.20E-06 | SPP1 Mac |
| TDRD3 | 0.76640654 | 0.672 | 0.195 | 4.02E-19 | SPP1 Mac |
| FAIM | 0.76628645 | 0.677 | 0.169 | 1.51E-25 | SPP1 Mac |
| HEXB | 0.75902401 | 0.964 | 0.753 | 2.03E-11 | SPP1 Mac |
| SEPP1 | 3.85601 | 0.853 | 0.114 | 7.94E-48 | IM |
| CCL13 | 2.73894651 | 0.488 | 0.088 | 3.94E-11 | IM |
| FOLR2 | 2.69659202 | 0.773 | 0.135 | 9.19E-43 | IM |
| CCL4L2 | 2.66590811 | 0.449 | 0.167 | 1.31E-08 | IM |
| CCL4 | 2.59733995 | 0.543 | 0.249 | 4.05E-12 | IM |
| RNASE1 | 2.35347352 | 0.771 | 0.283 | 3.03E-20 | IM |
| SLC40A1 | 2.25703194 | 0.58 | 0.053 | 1.62E-27 | IM |
| F13A1 | 2.21048906 | 0.6 | 0.053 | 5.16E-21 | IM |
| CCL3L3 | 2.18424327 | 0.408 | 0.115 | 2.12E-12 | IM |
| RGS1 | 2.16756033 | 0.869 | 0.429 | 1.04E-21 | IM |
| CCL3 | 2.10885831 | 0.645 | 0.321 | 4.67E-17 | IM |
| MS4A6A | 2.04380326 | 0.969 | 0.59 | 2.58E-32 | IM |
| PLTP | 1.9524435 | 0.655 | 0.175 | 8.42E-29 | IM |
| HLA-DQA1 | 1.92959524 | 0.888 | 0.724 | 2.30E-19 | IM |
| GPR34 | 1.60710746 | 0.663 | 0.208 | 1.20E-21 | IM |
| FOS. | 1.60160258 | 0.91 | 0.769 | 6.41E-17 | IM |
| PDK4 | 1.56948065 | 0.733 | 0.449 | 1.82E-15 | IM |
| EGR1 | 1.49985864 | 0.447 | 0.204 | 3.86E-05 | IM |
| TMEM176B | 1.49328318 | 0.686 | 0.285 | 2.53E-15 | IM |
| HSPA1A | 1.47365268 | 0.655 | 0.484 | 5.75E-06 | IM |
| HLA-DPA1 | 1.45846219 | 0.982 | 0.908 | 7.55E-19 | IM |

| | | | | | |
|---|---|---|---|---|---|
| LGMN | 1.45653631 | 0.814 | 0.418 | 6.27E-24 | IM |
| HLA-DPB1 | 1.39319475 | 0.992 | 0.909 | 2.98E-19 | IM |
| HLA-DRA | 1.3643967 | 1 | 0.975 | 3.02E-15 | IM |
| RGS2 | 1.33345224 | 0.849 | 0.566 | 3.17E-12 | IM |
| CD14 | 1.31522884 | 0.818 | 0.526 | 7.64E-12 | IM |
| TMEM176A | 1.28377237 | 0.6 | 0.211 | 2.86E-13 | IM |
| HLA-DRB1 | 1.2709972 | 0.976 | 0.9 | 3.85E-12 | IM |
| ZFP36 | 1.2600114 | 0.914 | 0.789 | 3.99E-17 | IM |
| FGL2 | 1.25959889 | 0.616 | 0.216 | 2.09E-09 | IM |
| STAB1 | 1.25706331 | 0.559 | 0.099 | 1.12E-19 | IM |
| FCGR2A | 1.25554556 | 0.873 | 0.697 | 6.52E-12 | IM |
| C1QC | 1.22475921 | 0.965 | 0.7 | 6.98E-21 | IM |
| RNASE6 | 1.22013946 | 0.747 | 0.366 | 2.38E-17 | IM |
| CFD | 1.21243501 | 0.798 | 0.666 | 3.15E-06 | IM |
| KLF6 | 1.19648085 | 0.857 | 0.767 | 9.19E-08 | IM |
| ITM2B | 1.17376983 | 0.976 | 0.912 | 4.02E-18 | IM |
| HLA-DQB1 | 1.16312003 | 0.918 | 0.795 | 7.50E-11 | IM |
| CD74 | 1.15791481 | 0.994 | 0.966 | 5.53E-17 | IM |
| MS4A4A | 1.15670895 | 0.853 | 0.715 | 2.99E-14 | IM |
| A2M | 1.12670712 | 0.576 | 0.248 | 1.55E-13 | IM |
| GPR183 | 1.09966967 | 0.81 | 0.462 | 1.53E-12 | IM |
| FOSB | 1.09735358 | 0.833 | 0.626 | 4.35E-12 | IM |
| HSPA1B | 1.08659709 | 0.49 | 0.284 | 9.52E-06 | IM |
| CEBPD | 1.07011346 | 0.831 | 0.56 | 6.12E-08 | IM |
| RBPJ | 1.06488615 | 0.741 | 0.62 | 2.77E-06 | IM |
| CH25H | 1.06135916 | 0.394 | 0.241 | 1 | IM |
| CTSC | 1.05751309 | 0.937 | 0.793 | 1.82E-11 | IM |
| MARCKS | 1.04330446 | 0.686 | 0.285 | 6.14E-08 | IM |
| TGFBI | 1.03229614 | 0.778 | 0.555 | 4.12E-09 | IM |
| HSPA1B | 3.45149326 | 0.973 | 0.282 | 3.89E-54 | HSP Macs |
| HSPA1A | 3.40864803 | 0.986 | 0.482 | 2.99E-52 | HSP Macs |
| DNAJB1 | 2.71512873 | 0.955 | 0.439 | 4.37E-44 | HSP Macs |
| HMOX1 | 2.42761666 | 0.765 | 0.417 | 3.31E-17 | HSP Macs |
| HSPA6 | 2.32280627 | 0.367 | 0.065 | 3.00E-09 | HSP Macs |
| HSPH1 | 2.22484257 | 0.914 | 0.422 | 7.56E-40 | HSP Macs |
| HSP90AA1 | 2.04657471 | 0.991 | 0.893 | 4.46E-35 | HSP Macs |
| ZFAND2A | 1.99348506 | 0.624 | 0.213 | 3.35E-18 | HSP Macs |
| JUN | 1.93222423 | 0.814 | 0.567 | 8.91E-16 | HSP Macs |
| HSPB1 | 1.87899589 | 0.91 | 0.736 | 5.83E-16 | HSP Macs |
| BAG3 | 1.75605516 | 0.588 | 0.138 | 2.15E-22 | HSP Macs |
| HSPE1 | 1.4843358 | 0.896 | 0.756 | 3.15E-23 | HSP Macs |
| FOS | 1.43317904 | 0.905 | 0.771 | 3.65E-12 | HSP Macs |
| IER5 | 1.39056828 | 0.597 | 0.331 | 1.15E-08 | HSP Macs |
| ANXA1 | 1.38597492 | 0.986 | 0.903 | 9.77E-22 | HSP Macs |
| KLF6 | 1.36363351 | 0.869 | 0.768 | 5.92E-08 | HSP Macs |
| DNAJA4 | 1.28589032 | 0.462 | 0.117 | 1.01E-10 | HSP Macs |
| HSP90AB1 | 1.28380937 | 0.941 | 0.8 | 8.06E-20 | HSP Macs |

| | | | | | |
|---|---|---|---|---|---|
| CCL18 | 1.28000788 | 0.792 | 0.648 | 1.80E-05 | HSP Macs |
| HSPD1 | 1.2784803 | 0.778 | 0.605 | 1.01E-14 | HSP Macs |
| CCL2 | 1.27520911 | 0.348 | 0.237 | 1.00E+00 | HSP Macs |
| KLF2 | 1.22033888 | 0.511 | 0.229 | 4.10E-03 | HSP Macs |
| ATF3 | 1.19856343 | 0.719 | 0.472 | 7.58E-11 | HSP Macs |
| APOE | 1.19760037 | 0.819 | 0.744 | 2.20E-06 | HSP Macs |
| PLIN2 | 1.1799052 | 0.955 | 0.866 | 3.20E-12 | HSP Macs |
| JUND | 1.16582132 | 0.783 | 0.624 | 1.13E-06 | HSP Macs |
| EGR1 | 1.10731628 | 0.33 | 0.209 | 4.19E-02 | HSP Macs |
| DNAJA1 | 1.10135586 | 0.842 | 0.661 | 1.37E-07 | HSP Macs |
| RHOB | 1.05467752 | 0.724 | 0.618 | 8.66E-06 | HSP Macs |
| SQSTM1 | 1.03634802 | 0.774 | 0.709 | 1.53E-04 | HSP Macs |
| UBC | 1.0209344 | 0.932 | 0.892 | 2.64E-11 | HSP Macs |
| HSPA8 | 0.97258278 | 0.873 | 0.77 | 8.83E-07 | HSP Macs |
| HSPA5 | 0.95881426 | 0.733 | 0.62 | 1.36E-02 | HSP Macs |
| GADD45B | 0.94224742 | 0.656 | 0.582 | 1.00E+00 | HSP Macs |
| ADM | 0.91452996 | 0.471 | 0.28 | 7.05E-01 | HSP Macs |
| PDK4 | 0.91173134 | 0.525 | 0.455 | 1.00E+00 | HSP Macs |
| NEAT1 | 0.89175668 | 0.982 | 0.952 | 1.00E+00 | HSP Macs |
| RNASE1 | 0.88765708 | 0.534 | 0.291 | 1.41E-03 | HSP Macs |
| DNAJB4 | 0.83698088 | 0.348 | 0.082 | 4.74E-07 | HSP Macs |
| FOSB | 0.82605003 | 0.701 | 0.63 | 3.11E-01 | HSP Macs |
| KLF4 | 0.81696429 | 0.833 | 0.791 | 1.58E-03 | HSP Macs |
| DNAJB6 | 0.81278618 | 0.76 | 0.687 | 7.93E-07 | HSP Macs |
| CACYBP | 0.79850298 | 0.606 | 0.521 | 4.50E-04 | HSP Macs |
| TSC22D3 | 0.79806146 | 0.805 | 0.773 | 1.52E-03 | HSP Macs |
| WBP5 | 0.79287361 | 0.489 | 0.36 | 9.94E-05 | HSP Macs |
| CHORDC1 | 0.78386692 | 0.412 | 0.228 | 9.51E-03 | HSP Macs |
| MGST1 | 0.76307624 | 0.579 | 0.502 | 1.00E+00 | HSP Macs |
| TXN | 0.75654785 | 0.968 | 0.924 | 2.82E-02 | HSP Macs |
| DUSP1 | 0.74748777 | 0.941 | 0.953 | 7.16E-01 | HSP Macs |
| MRPL18 | 0.73854484 | 0.588 | 0.51 | 2.05E-01 | HSP Macs |
| MT1X | 1.48970415 | 0.595 | 0.395 | 1 | MT Mac |
| SPP1 | 1.48495138 | 0.473 | 0.259 | 0.00114967 | MT Mac |
| MT1H | 1.38383915 | 0.129 | 0.041 | 1 | MT Mac |
| MT1E | 1.06852879 | 0.411 | 0.264 | 1 | MT Mac |
| C15orf48 | 0.92018586 | 0.722 | 0.465 | 1.93E-06 | MT Mac |
| MT2A | 0.91509884 | 0.877 | 0.812 | 1 | MT Mac |
| EMP1 | 0.86588618 | 0.727 | 0.394 | 4.99E-08 | MT Mac |
| AREG | 0.80507119 | 0.853 | 0.703 | 6.18E-06 | MT Mac |
| SDC2 | 0.69957688 | 0.579 | 0.367 | 0.1080501 | MT Mac |
| CLEC5A | 0.68784348 | 0.419 | 0.124 | 0.0006888 | MT Mac |
| IL1RN | 0.67000316 | 0.536 | 0.374 | 1 | MT Mac |
| ARID5B | 0.64837762 | 0.532 | 0.337 | 0.00742966 | MT Mac |
| HCST | 0.63959823 | 0.781 | 0.641 | 0.00129729 | MT Mac |
| GPR183 | 0.63943032 | 0.764 | 0.453 | 0.00012583 | MT Mac |
| S100A10 | 0.63321758 | 0.993 | 0.979 | 2.40E-06 | MT Mac |

| | | | | | |
|---|---|---|---|---|---|
| MATK | 0.6260184 | 0.364 | 0.123 | 9.26E-08 | MT Mac |
| ALCAM | 0.61545142 | 0.731 | 0.539 | 0.00243624 | MT Mac |
| TREM2 | 0.61249955 | 0.693 | 0.514 | 0.00154851 | MT Mac |
| IL3RA | 0.61210363 | 0.456 | 0.264 | 3.49E-08 | MT Mac |
| FCGR2B | 0.60282261 | 0.428 | 0.179 | 0.02894865 | MT Mac |
| RP11-1008C: | 0.59927403 | 0.249 | 0.148 | 1 | MT Mac |
| LPL | 0.57270558 | 0.573 | 0.492 | 1 | MT Mac |
| SH3BGRL3 | 0.562112 | 0.997 | 0.982 | 4.25E-10 | MT Mac |
| LSP1 | 0.55973611 | 0.595 | 0.401 | 0.00065357 | MT Mac |
| EMP3 | 0.55754907 | 0.98 | 0.909 | 1 | MT Mac |
| ID3 | 0.54665534 | 0.431 | 0.308 | 1 | MT Mac |
| LMNA | 0.54002112 | 0.889 | 0.791 | 0.00068867 | MT Mac |
| CAMK1 | 0.53999492 | 0.359 | 0.158 | 1 | MT Mac |
| LIMS1 | 0.53602408 | 0.883 | 0.771 | 6.06E-06 | MT Mac |
| SMIM3 | 0.53162049 | 0.612 | 0.373 | 0.00043725 | MT Mac |
| MT1F | 0.53078865 | 0.335 | 0.25 | 1 | MT Mac |
| SDS | 0.52254627 | 0.25 | 0.146 | 1 | MT Mac |
| HMGA1 | 0.51605921 | 0.67 | 0.518 | 1 | MT Mac |
| RGCC | 0.5148348 | 0.961 | 0.854 | 0.05185937 | MT Mac |
| GSN | 0.51095377 | 0.781 | 0.644 | 1 | MT Mac |
| TYMP | 0.50992852 | 0.97 | 0.905 | 0.26704005 | MT Mac |
| NEAT1 | 0.50065801 | 0.981 | 0.95 | 8.69E-07 | MT Mac |
| MRC1 | 0.49590668 | 0.857 | 0.736 | 0.00160451 | MT Mac |
| HERPUD1 | 0.49270317 | 0.793 | 0.686 | 0.00486845 | MT Mac |
| SLC16A10 | 0.48833925 | 0.675 | 0.47 | 3.08E-06 | MT Mac |
| BTG1 | 0.48450413 | 0.972 | 0.929 | 3.97E-05 | MT Mac |
| QPCT | 0.47125754 | 0.438 | 0.259 | 1 | MT Mac |
| CALM1 | 0.4653728 | 0.966 | 0.912 | 0.00124927 | MT Mac |
| NME1 | 0.45879934 | 0.579 | 0.445 | 0.196099 | MT Mac |
| BASP1 | 0.45047001 | 0.647 | 0.447 | 1 | MT Mac |
| CTSH | 0.44192477 | 0.796 | 0.696 | 1 | MT Mac |
| LGALS1 | 0.43615744 | 0.994 | 0.982 | 0.00014161 | MT Mac |
| SGK1 | 0.43320972 | 0.689 | 0.586 | 1 | MT Mac |
| RHOC | 0.43294499 | 0.523 | 0.384 | 0.00190206 | MT Mac |
| LILRB4 | 0.42366559 | 0.556 | 0.407 | 1 | MT Mac |
| S100A8 | 3.79079165 | 0.887 | 0.564 | 7.07E-30 | CMonos |
| S100A9 | 3.23543314 | 0.934 | 0.859 | 1.04E-23 | CMonos |
| IL1B | 3.1520615 | 0.76 | 0.415 | 7.98E-26 | CMonos |
| S100A12 | 3.10220768 | 0.488 | 0.034 | 4.14E-18 | CMonos |
| SERPINB2 | 3.0936022 | 0.443 | 0.037 | 1.25E-16 | CMonos |
| G0S2 | 2.88538916 | 0.736 | 0.282 | 7.70E-25 | CMonos |
| CXCL8 | 2.64294018 | 0.806 | 0.494 | 8.98E-19 | CMonos |
| THBS1 | 2.5780012 | 0.859 | 0.564 | 9.66E-25 | CMonos |
| TIMP1 | 2.53488843 | 0.951 | 0.784 | 2.52E-36 | CMonos |
| EREG | 2.45413414 | 0.807 | 0.388 | 3.85E-29 | CMonos |
| VCAN | 2.38199459 | 0.752 | 0.346 | 1.07E-31 | CMonos |
| FCN1 | 2.18898443 | 0.691 | 0.167 | 6.08E-22 | CMonos |

| | | | | | |
|---|---|---|---|---|---|
| SOD2 | 2.06728182 | 0.939 | 0.846 | 2.13E-23 | CMonos |
| NAMPT | 2.00994546 | 0.933 | 0.829 | 1.54E-30 | CMonos |
| CXCL2 | 1.98304508 | 0.845 | 0.66 | 1.42E-23 | CMonos |
| CCL20 | 1.91419688 | 0.579 | 0.412 | 0.00033008 | CMonos |
| PLAUR | 1.85842034 | 0.945 | 0.865 | 2.69E-28 | CMonos |
| SLC2A3 | 1.83409523 | 0.786 | 0.537 | 1.49E-14 | CMonos |
| IL1R2 | 1.78608239 | 0.416 | 0.099 | 1.30E-08 | CMonos |
| NLRP3 | 1.76060151 | 0.573 | 0.168 | 4.73E-11 | CMonos |
| IER3 | 1.73762214 | 0.753 | 0.462 | 3.24E-23 | CMonos |
| PTGS2 | 1.70279238 | 0.345 | 0.096 | 5.39E-13 | CMonos |
| AREG | 1.66303818 | 0.881 | 0.695 | 3.34E-21 | CMonos |
| CD93 | 1.65208909 | 0.505 | 0.108 | 1.98E-19 | CMonos |
| CCL3 | 1.64708758 | 0.369 | 0.324 | 1 | CMonos |
| CD300E | 1.64478787 | 0.518 | 0.128 | 5.23E-16 | CMonos |
| SAMSN1 | 1.63144528 | 0.857 | 0.668 | 4.37E-23 | CMonos |
| PPIF | 1.62449698 | 0.608 | 0.412 | 3.10E-10 | CMonos |
| CCL3L3 | 1.59609008 | 0.183 | 0.116 | 1 | CMonos |
| SRGN | 1.59431675 | 0.995 | 0.977 | 1.64E-38 | CMonos |
| CXCL1 | 1.59140233 | 0.211 | 0.111 | 1 | CMonos |
| CXCL3 | 1.54994151 | 0.749 | 0.75 | 0.02706392 | CMonos |
| SLC25A37 | 1.50265726 | 0.536 | 0.253 | 8.42E-12 | CMonos |
| CTB-61M7.2 | 1.48946517 | 0.415 | 0.085 | 2.38E-11 | CMonos |
| CREM | 1.43244233 | 0.787 | 0.617 | 1.79E-11 | CMonos |
| C15orf48 | 1.41968059 | 0.452 | 0.482 | 1 | CMonos |
| BCL2A1 | 1.4082114 | 0.713 | 0.704 | 2.38E-05 | CMonos |
| SERPINB9 | 1.4069376 | 0.489 | 0.155 | 2.43E-12 | CMonos |
| RP11-1143G9 | 1.3577398 | 0.672 | 0.623 | 0.00244188 | CMonos |
| MXD1 | 1.2644334 | 0.633 | 0.431 | 5.39E-10 | CMonos |
| BTG1 | 1.25754618 | 0.961 | 0.928 | 3.99E-20 | CMonos |
| VEGFA | 1.25354312 | 0.469 | 0.226 | 2.09E-06 | CMonos |
| TNFAIP3 | 1.24038986 | 0.627 | 0.542 | 0.00176147 | CMonos |
| CYP1B1 | 1.23586165 | 0.319 | 0.158 | 0.43960461 | CMonos |
| RILPL2 | 1.22396108 | 0.704 | 0.654 | 1.52E-09 | CMonos |
| ATP2B1 | 1.22295686 | 0.627 | 0.583 | 2.02E-05 | CMonos |
| NFKBIA | 1.21350891 | 0.877 | 0.915 | 1.51E-08 | CMonos |
| C5AR1 | 1.16322189 | 0.773 | 0.726 | 2.64E-09 | CMonos |
| IFITM2 | 1.16172627 | 0.574 | 0.44 | 0.0003502 | CMonos |
| TNFRSF1B | 1.15926171 | 0.469 | 0.257 | 2.10E-09 | CMonos |
| IFITM2 | 2.57978527 | 0.864 | 0.431 | 8.26E-35 | NCMonos |
| FCN1 | 2.31007287 | 0.809 | 0.183 | 9.14E-41 | NCMonos |
| IFITM3 | 2.23399529 | 0.886 | 0.578 | 4.34E-33 | NCMonos |
| CXCL10 | 2.11844054 | 0.151 | 0.039 | 1 | NCMonos |
| TIMP1 | 2.04214814 | 0.969 | 0.79 | 8.87E-35 | NCMonos |
| NAMPT | 1.83588439 | 0.966 | 0.832 | 3.84E-40 | NCMonos |
| THBS1 | 1.73177025 | 0.853 | 0.576 | 4.61E-23 | NCMonos |
| FPR2 | 1.72482458 | 0.653 | 0.307 | 1.07E-20 | NCMonos |
| APOBEC3A | 1.68588661 | 0.413 | 0.04 | 3.87E-18 | NCMonos |

| | | | | | |
|---|---|---|---|---|---|
| G0S2 | 1.61610737 | 0.633 | 0.305 | 2.64E-12 | NCMonos |
| CFP | 1.5904601 | 0.61 | 0.118 | 2.47E-26 | NCMonos |
| FGL2 | 1.58111958 | 0.664 | 0.203 | 2.69E-23 | NCMonos |
| RGS2 | 1.5358217 | 0.85 | 0.558 | 9.54E-24 | NCMonos |
| CD48 | 1.52124534 | 0.653 | 0.231 | 5.00E-21 | NCMonos |
| LST1 | 1.51545625 | 0.938 | 0.825 | 1.48E-26 | NCMonos |
| CORO1A | 1.49583142 | 0.705 | 0.293 | 1.70E-17 | NCMonos |
| HES4 | 1.49011975 | 0.432 | 0.098 | 1.40E-13 | NCMonos |
| WARS | 1.4881918 | 0.548 | 0.354 | 1.63E-07 | NCMonos |
| ISG20 | 1.45849936 | 0.492 | 0.111 | 3.03E-15 | NCMonos |
| SOD2 | 1.4554022 | 0.939 | 0.85 | 3.51E-21 | NCMonos |
| MT2A | 1.43764685 | 0.89 | 0.812 | 1.81E-07 | NCMonos |
| FAM26F | 1.43569 | 0.525 | 0.232 | 7.45E-14 | NCMonos |
| CD300E | 1.42998756 | 0.589 | 0.141 | 7.25E-19 | NCMonos |
| SRGN | 1.38766777 | 0.997 | 0.977 | 1.92E-35 | NCMonos |
| BIRC3 | 1.36316732 | 0.55 | 0.222 | 4.87E-15 | NCMonos |
| VAMP5 | 1.35428318 | 0.571 | 0.278 | 1.22E-14 | NCMonos |
| SOCS3 | 1.34232597 | 0.789 | 0.579 | 2.41E-17 | NCMonos |
| DDIT4 | 1.33278 | 0.652 | 0.374 | 3.76E-13 | NCMonos |
| PLAC8 | 1.3261919 | 0.438 | 0.182 | 2.09E-07 | NCMonos |
| DUSP2 | 1.32125215 | 0.585 | 0.291 | 3.22E-12 | NCMonos |
| SERPINB9 | 1.28560936 | 0.571 | 0.165 | 1.06E-22 | NCMonos |
| LILRA5 | 1.27828856 | 0.448 | 0.104 | 2.77E-10 | NCMonos |
| SAT1 | 1.2754691 | 1 | 0.996 | 7.61E-34 | NCMonos |
| AIF1 | 1.24830275 | 0.976 | 0.924 | 3.65E-26 | NCMonos |
| CEBPD | 1.24236246 | 0.764 | 0.557 | 9.46E-14 | NCMonos |
| FPR1 | 1.23218248 | 0.68 | 0.441 | 2.10E-13 | NCMonos |
| SAMSN1 | 1.23183113 | 0.872 | 0.675 | 9.16E-24 | NCMonos |
| LILRB2 | 1.21633587 | 0.477 | 0.176 | 1.57E-08 | NCMonos |
| C10orf54 | 1.16451641 | 0.536 | 0.238 | 5.73E-09 | NCMonos |
| CREM | 1.15915103 | 0.778 | 0.624 | 1.12E-13 | NCMonos |
| COTL1 | 1.15243419 | 0.955 | 0.911 | 2.41E-16 | NCMonos |
| EREG | 1.13371185 | 0.664 | 0.412 | 7.14E-05 | NCMonos |
| C5AR1 | 1.13108931 | 0.834 | 0.725 | 2.90E-17 | NCMonos |
| SLC2A3 | 1.12661018 | 0.712 | 0.551 | 2.03E-12 | NCMonos |
| RNF144B | 1.1202757 | 0.539 | 0.354 | 0.00017095 | NCMonos |
| CLEC4E | 1.11895621 | 0.472 | 0.248 | 1.05E-07 | NCMonos |
| INSIG1 | 1.07856402 | 0.574 | 0.391 | 1.05E-05 | NCMonos |
| HMGB2 | 1.06071533 | 0.64 | 0.557 | 6.93E-08 | NCMonos |
| ADM | 1.0504134 | 0.474 | 0.273 | 3.34E-05 | NCMonos |
| LILRA1 | 1.03790254 | 0.324 | 0.034 | 6.08E-06 | NCMonos |
| CCL2 | 1.57461397 | 0.346 | 0.225 | 1 | Transitional_MonoMacs |
| CCL7 | 1.00359063 | 0.163 | 0.063 | 1 | Transitional_MonoMacs |
| RNASE1 | 0.90218158 | 0.486 | 0.27 | 0.09235537 | Transitional_MonoMacs |
| JUND | 0.74529113 | 0.606 | 0.628 | 1 | Transitional_MonoMacs |
| SPP1 | 0.64386258 | 0.38 | 0.257 | 1 | Transitional_MonoMacs |
| LGALS1 | 0.63143831 | 0.987 | 0.982 | 0.00983055 | Transitional_MonoMacs |

| | | | | | |
|---|---|---|---|---|---|
| AP2S1 | 0.62054987 | 0.881 | 0.917 | 0.00068491 | Transitional_MonoMacs |
| ATP5I | 0.60137242 | 0.693 | 0.776 | 0.41193732 | Transitional_MonoMacs |
| C4orf48 | 0.59967684 | 0.656 | 0.724 | 1 | Transitional_MonoMacs |
| SEC61G | 0.56331654 | 0.755 | 0.843 | 1 | Transitional_MonoMacs |
| TMIGD3 | 0.55618491 | 0.183 | 0.117 | 1 | Transitional_MonoMacs |
| FTL | 0.54568784 | 1 | 1 | 0.00055916 | Transitional_MonoMacs |
| CCL20 | 0.53111196 | 0.488 | 0.418 | 1 | Transitional_MonoMacs |
| CTSL | 0.49920347 | 0.864 | 0.859 | 0.07307932 | Transitional_MonoMacs |
| MT-ATP6 | 0.49459272 | 0.99 | 0.989 | 2.87E-09 | Transitional_MonoMacs |
| POLR2L | 0.48734668 | 0.677 | 0.786 | 1 | Transitional_MonoMacs |
| ELL2 | 0.46972797 | 0.504 | 0.56 | 1 | Transitional_MonoMacs |
| TNFAIP3 | 0.43605729 | 0.544 | 0.55 | 1 | Transitional_MonoMacs |
| ATP5E | 0.42641379 | 0.984 | 0.99 | 0.01882166 | Transitional_MonoMacs |
| CTSB | 0.42224165 | 0.963 | 0.952 | 0.40345631 | Transitional_MonoMacs |
| CD163 | 0.42211917 | 0.749 | 0.804 | 1 | Transitional_MonoMacs |
| GPX4 | 0.41602745 | 0.955 | 0.961 | 2.45E-05 | Transitional_MonoMacs |
| EMP3 | 0.39570018 | 0.848 | 0.921 | 1 | Transitional_MonoMacs |
| BASP1 | 0.35773478 | 0.453 | 0.459 | 1 | Transitional_MonoMacs |
| CXCL2 | 0.35686362 | 0.727 | 0.669 | 1 | Transitional_MonoMacs |
| MCL1 | 0.34944251 | 0.853 | 0.929 | 0.02662648 | Transitional_MonoMacs |
| NDUFB2 | 0.34923039 | 0.775 | 0.88 | 1 | Transitional_MonoMacs |
| S100A6 | 0.34434196 | 0.996 | 0.995 | 1 | Transitional_MonoMacs |
| RPL35 | 0.34367012 | 0.98 | 0.991 | 0.00206018 | Transitional_MonoMacs |
| H2AFY | 0.33908112 | 0.802 | 0.893 | 1 | Transitional_MonoMacs |
| RPL12 | 0.33668992 | 0.994 | 0.998 | 5.29E-06 | Transitional_MonoMacs |
| UQCR11 | 0.32908437 | 0.805 | 0.898 | 1 | Transitional_MonoMacs |
| CTSD | 0.31537102 | 0.979 | 0.935 | 1 | Transitional_MonoMacs |
| TCEB2 | 0.31473228 | 0.883 | 0.925 | 0.01737998 | Transitional_MonoMacs |
| GPX1 | 0.30497291 | 0.985 | 0.983 | 0.04312825 | Transitional_MonoMacs |
| MT-ND3 | 0.2954957 | 0.934 | 0.962 | 1 | Transitional_MonoMacs |
| RPL41 | 0.28483119 | 0.998 | 0.999 | 0.89694293 | Transitional_MonoMacs |
| RPL36 | 0.27262846 | 0.954 | 0.982 | 1 | Transitional_MonoMacs |
| CSTB | 0.26148937 | 0.975 | 0.952 | 1 | Transitional_MonoMacs |
| RPS12 | 0.25946055 | 0.994 | 0.997 | 1 | Transitional_MonoMacs |
| MRPL23 | 0.25771006 | 0.35 | 0.519 | 1 | Transitional_MonoMacs |
| MFSD10 | -0.2508441 | 0.124 | 0.408 | 2.15E-05 | Transitional_MonoMacs |
| PPA1 | -0.2513253 | 0.243 | 0.542 | 0.01873545 | Transitional_MonoMacs |
| INSIG1 | -0.251514 | 0.261 | 0.416 | 1 | Transitional_MonoMacs |
| RHOG | -0.25183 | 0.387 | 0.701 | 1 | Transitional_MonoMacs |
| SVIL | -0.2534915 | 0.084 | 0.317 | 2.79E-05 | Transitional_MonoMacs |
| LAMP2 | -0.2537699 | 0.34 | 0.644 | 1 | Transitional_MonoMacs |
| C10orf54 | -0.2538893 | 0.1 | 0.271 | 0.33108989 | Transitional_MonoMacs |
| PLTP | -0.2540992 | 0.08 | 0.198 | 1 | Transitional_MonoMacs |
| HCK | -0.2541226 | 0.234 | 0.532 | 0.00154281 | Transitional_MonoMacs |
| CCL17 | 4.43204369 | 0.347 | 0.024 | 1.45E-13 | DC |
| GPR183 | 2.29065801 | 0.928 | 0.448 | 1.36E-33 | DC |
| RGS1 | 2.21684112 | 0.887 | 0.417 | 3.09E-32 | DC |

| | | | | | |
|---|---|---|---|---|---|
| S100B | 2.1519708 | 0.349 | 0.036 | 5.34E-11 | DC |
| HLA-DPB1 | 2.13093835 | 1 | 0.907 | 2.92E-45 | DC |
| CXCR4 | 2.08915933 | 0.86 | 0.556 | 4.78E-21 | DC |
| CST7 | 2.05430596 | 0.518 | 0.03 | 1.85E-24 | DC |
| HLA-DQA1 | 1.97756379 | 0.96 | 0.717 | 2.77E-33 | DC |
| AREG | 1.8363837 | 0.903 | 0.702 | 2.79E-20 | DC |
| INSIG1 | 1.80319678 | 0.713 | 0.384 | 7.90E-12 | DC |
| HLA-DPA1 | 1.78921088 | 0.997 | 0.906 | 7.28E-33 | DC |
| HLA-DQB1 | 1.77840176 | 0.986 | 0.789 | 1.67E-33 | DC |
| FCER1A | 1.7679413 | 0.375 | 0.005 | 1.00E-15 | DC |
| CD74 | 1.64485493 | 0.999 | 0.965 | 2.88E-38 | DC |
| HERPUD1 | 1.64221051 | 0.897 | 0.682 | 1.59E-18 | DC |
| DUSP4 | 1.5625146 | 0.62 | 0.197 | 1.74E-12 | DC |
| RGS2 | 1.5379443 | 0.866 | 0.558 | 1.57E-15 | DC |
| BIRC3 | 1.52564511 | 0.414 | 0.229 | 1 | DC |
| CLEC10A | 1.51418163 | 0.505 | 0.064 | 7.10E-14 | DC |
| CCL22 | 1.45289704 | 0.189 | 0.01 | 9.51E-05 | DC |
| REL | 1.4485663 | 0.832 | 0.625 | 8.00E-15 | DC |
| PKIB | 1.44316367 | 0.459 | 0.023 | 6.15E-20 | DC |
| CEBPD | 1.4333968 | 0.843 | 0.553 | 2.16E-14 | DC |
| HLA-DRB5 | 1.41170174 | 0.931 | 0.751 | 1.11E-15 | DC |
| HLA-DRA | 1.4093591 | 1 | 0.974 | 1.69E-31 | DC |
| ZFP36 | 1.39077393 | 0.935 | 0.785 | 1.14E-17 | DC |
| CCR7 | 1.38860328 | 0.175 | 0.011 | 0.00011239 | DC |
| HLA-DRB1 | 1.38013597 | 0.992 | 0.898 | 1.48E-24 | DC |
| CREM | 1.35113167 | 0.834 | 0.622 | 9.33E-13 | DC |
| CD1C | 1.3501395 | 0.297 | 0.008 | 2.06E-11 | DC |
| HLA-DQA2 | 1.29926375 | 0.579 | 0.33 | 4.14E-07 | DC |
| IL1R2 | 1.29016379 | 0.461 | 0.11 | 5.45E-08 | DC |
| RNASE6 | 1.27469632 | 0.739 | 0.357 | 6.79E-16 | DC |
| CD83 | 1.26599461 | 0.716 | 0.58 | 6.13E-05 | DC |
| FCGR2B | 1.2224603 | 0.535 | 0.177 | 3.60E-13 | DC |
| PPA1 | 1.20876306 | 0.699 | 0.5 | 4.12E-08 | DC |
| MS4A6A | 1.18239219 | 0.822 | 0.587 | 3.95E-11 | DC |
| BTG1 | 1.18238315 | 0.949 | 0.93 | 6.98E-12 | DC |
| LTB | 1.17267981 | 0.222 | 0.057 | 0.00065616 | DC |
| DDIT4 | 1.16918166 | 0.706 | 0.372 | 2.43E-12 | DC |
| SERPINB9 | 1.16098282 | 0.592 | 0.165 | 1.63E-12 | DC |
| RAMP1 | 1.14696566 | 0.276 | 0.013 | 5.45E-11 | DC |
| MALAT1 | 1.0916886 | 0.969 | 0.986 | 6.11E-22 | DC |
| AXL | 1.05690992 | 0.506 | 0.282 | 0.00034066 | DC |
| CSF2RA | 1.05179501 | 0.64 | 0.304 | 2.63E-16 | DC |
| CST3 | 1.05008324 | 0.982 | 0.978 | 4.81E-09 | DC |
| PPP1R14A | 1.03614914 | 0.275 | 0.019 | 8.87E-09 | DC |
| SPINT2 | 1.02152783 | 0.715 | 0.481 | 9.46E-11 | DC |
| CD1E | 1.00272529 | 0.231 | 0.004 | 1.75E-09 | DC |
| CORO1A | 1.00146768 | 0.697 | 0.294 | 1.24E-12 | DC |

| | | | | | |
|---|---|---|---|---|---|
| STMN1 | 2.48450606 | 0.953 | 0.408 | 5.85E-50 | Cycling Cells |
| KIAA0101 | 2.35711426 | 0.865 | 0.021 | 1.37E-53 | Cycling Cells |
| H2AFZ | 2.18464574 | 0.994 | 0.842 | 5.98E-49 | Cycling Cells |
| HIST1H4C | 2.07955217 | 0.674 | 0.134 | 1.74E-27 | Cycling Cells |
| TUBB | 1.86670161 | 0.974 | 0.746 | 3.59E-36 | Cycling Cells |
| TUBA1B | 1.85161495 | 0.999 | 0.855 | 1.70E-35 | Cycling Cells |
| HMGN2 | 1.79951435 | 0.981 | 0.781 | 1.73E-39 | Cycling Cells |
| UBE2C | 1.7937594 | 0.647 | 0.006 | 1.30E-39 | Cycling Cells |
| PTTG1 | 1.74729111 | 0.666 | 0.091 | 2.39E-30 | Cycling Cells |
| TYMS | 1.6751937 | 0.725 | 0.01 | 4.67E-44 | Cycling Cells |
| IGKC | 1.63530261 | 0.247 | 0.143 | 1 | Cycling Cells |
| CKS1B | 1.60799869 | 0.906 | 0.316 | 2.28E-41 | Cycling Cells |
| TK1 | 1.599076 | 0.778 | 0.028 | 2.09E-44 | Cycling Cells |
| HMGB1 | 1.42194048 | 0.993 | 0.866 | 3.13E-35 | Cycling Cells |
| HMGB2 | 1.37238922 | 0.901 | 0.55 | 1.70E-19 | Cycling Cells |
| RRM2 | 1.33165399 | 0.541 | 0.003 | 2.83E-27 | Cycling Cells |
| PCNA | 1.31735719 | 0.703 | 0.208 | 1.39E-20 | Cycling Cells |
| H2AFV | 1.30100015 | 0.935 | 0.631 | 3.96E-21 | Cycling Cells |
| CDK1 | 1.27004251 | 0.625 | 0.014 | 5.85E-33 | Cycling Cells |
| DUT | 1.25324015 | 0.872 | 0.504 | 1.05E-20 | Cycling Cells |
| DEK | 1.24662946 | 0.929 | 0.633 | 1.39E-28 | Cycling Cells |
| CDKN3 | 1.20707655 | 0.591 | 0.02 | 2.51E-30 | Cycling Cells |
| TMEM106C | 1.20407816 | 0.716 | 0.112 | 2.14E-33 | Cycling Cells |
| BIRC5 | 1.20067461 | 0.59 | 0.004 | 1.19E-32 | Cycling Cells |
| TOP2A | 1.17938337 | 0.524 | 0.003 | 6.61E-27 | Cycling Cells |
| MKI67 | 1.17459362 | 0.566 | 0.002 | 1.15E-30 | Cycling Cells |
| NUCKS1 | 1.17442725 | 0.924 | 0.542 | 4.37E-18 | Cycling Cells |
| SMC4 | 1.10444236 | 0.715 | 0.143 | 1.69E-30 | Cycling Cells |
| FN1 | 1.08999563 | 0.753 | 0.44 | 6.33E-06 | Cycling Cells |
| CENPW | 1.08461649 | 0.791 | 0.204 | 2.49E-33 | Cycling Cells |
| IDH2 | 1.0806651 | 0.809 | 0.369 | 1.07E-18 | Cycling Cells |
| NUSAP1 | 1.07867764 | 0.578 | 0.034 | 4.70E-28 | Cycling Cells |
| ANP32B | 1.07624086 | 0.937 | 0.697 | 2.87E-21 | Cycling Cells |
| CKS2 | 1.04941906 | 0.796 | 0.365 | 2.83E-13 | Cycling Cells |
| RPA3 | 1.04070884 | 0.84 | 0.397 | 2.48E-21 | Cycling Cells |
| CENPM | 1.02841445 | 0.624 | 0.01 | 4.48E-34 | Cycling Cells |
| CARHSP1 | 0.96350508 | 0.713 | 0.207 | 8.93E-21 | Cycling Cells |
| RANBP1 | 0.95458319 | 0.91 | 0.627 | 5.79E-13 | Cycling Cells |
| CENPF | 0.95298072 | 0.497 | 0.011 | 1.68E-24 | Cycling Cells |
| DTYMK | 0.92408446 | 0.722 | 0.181 | 6.81E-29 | Cycling Cells |
| SNRNP25 | 0.89401854 | 0.744 | 0.29 | 2.28E-18 | Cycling Cells |
| SKA2 | 0.86438795 | 0.694 | 0.126 | 8.22E-24 | Cycling Cells |
| STRA13 | 0.86065917 | 0.803 | 0.416 | 9.25E-13 | Cycling Cells |
| HMGB3 | 0.85499774 | 0.616 | 0.113 | 1.09E-19 | Cycling Cells |
| RAD51AP1 | 0.85022527 | 0.629 | 0.047 | 1.05E-28 | Cycling Cells |
| SIVA1 | 0.84799645 | 0.843 | 0.536 | 1.78E-13 | Cycling Cells |
| SMC2 | 0.83891804 | 0.668 | 0.109 | 3.00E-26 | Cycling Cells |
| MZT2B | 0.83289051 | 0.89 | 0.582 | 9.93E-09 | Cycling Cells |

| | | | | | |
|---|---|---|---|---|---|
| RAN | 0.83209591 | 0.96 | 0.799 | 1.20E-11 | Cycling Cells |
| HMGN1 | 0.82926681 | 0.947 | 0.727 | 5.79E-18 | Cycling Cells |
| NOP10 | 0.78163653 | 0.795 | 0.903 | 0.10103951 | Mac_UD |
| UBB | 0.6868254 | 0.812 | 0.889 | 1 | Mac_UD |
| PRDX1 | 0.6643947 | 0.75 | 0.868 | 0.88089653 | Mac_UD |
| FTL | 0.65347801 | 1 | 1 | 4.56E-05 | Mac_UD |
| VAMP8 | 0.61157592 | 0.757 | 0.904 | 1 | Mac_UD |
| RPL31 | 0.60928853 | 0.899 | 0.979 | 0.00977935 | Mac_UD |
| FABP4 | 0.5969553 | 0.615 | 0.464 | 1 | Mac_UD |
| RAB8A | 0.53554401 | 0.208 | 0.438 | 1 | Mac_UD |
| GTF2H5 | 0.52629887 | 0.243 | 0.504 | 1 | Mac_UD |
| YBX1 | 0.517464 | 0.955 | 0.982 | 0.19254684 | Mac_UD |
| PSMB6 | 0.51220216 | 0.354 | 0.668 | 1 | Mac_UD |
| PSMD8 | 0.5105298 | 0.347 | 0.633 | 1 | Mac_UD |
| S100A11 | 0.50259827 | 0.983 | 0.991 | 0.01435749 | Mac_UD |
| ATP5E | 0.50255747 | 0.944 | 0.99 | 0.00499542 | Mac_UD |
| TUBA1B | 0.4981575 | 0.715 | 0.861 | 1 | Mac_UD |
| S100A13 | 0.49795973 | 0.247 | 0.434 | 1 | Mac_UD |
| COPZ1 | 0.47968388 | 0.229 | 0.476 | 1 | Mac_UD |
| RPL10A | 0.47841459 | 0.917 | 0.977 | 0.08300112 | Mac_UD |
| RPS20 | 0.46899393 | 0.965 | 0.993 | 0.00145517 | Mac_UD |
| PSMC1 | 0.46825117 | 0.201 | 0.468 | 1 | Mac_UD |
| UQCRB | 0.46260047 | 0.795 | 0.942 | 1 | Mac_UD |
| NDUFA6 | 0.46137644 | 0.309 | 0.616 | 1 | Mac_UD |
| FTH1 | 0.45970467 | 1 | 1 | 0.04360204 | Mac_UD |
| COA6 | 0.4525513 | 0.191 | 0.398 | 1 | Mac_UD |
| PPCS | 0.44687921 | 0.247 | 0.5 | 1 | Mac_UD |
| RPS25 | 0.4456335 | 0.962 | 0.989 | 0.00015756 | Mac_UD |
| TMEM126B | 0.44545189 | 0.177 | 0.41 | 1 | Mac_UD |
| TTC1 | 0.44374287 | 0.181 | 0.336 | 1 | Mac_UD |
| MRPS7 | 0.44277152 | 0.17 | 0.411 | 1 | Mac_UD |
| SSBP1 | 0.44227585 | 0.312 | 0.616 | 1 | Mac_UD |
| HDDC2 | 0.44202816 | 0.257 | 0.501 | 1 | Mac_UD |
| TCEB2 | 0.43969623 | 0.764 | 0.923 | 1 | Mac_UD |
| RPL26 | 0.43887267 | 0.969 | 0.992 | 0.00146587 | Mac_UD |
| MRPS18B | 0.43606804 | 0.149 | 0.35 | 1 | Mac_UD |
| SNX2 | 0.43347487 | 0.306 | 0.603 | 1 | Mac_UD |
| RPL37A | 0.43327795 | 0.934 | 0.983 | 0.06118806 | Mac_UD |
| NAA38 | 0.43307247 | 0.201 | 0.507 | 0.00090965 | Mac_UD |
| CCT7 | 0.42993485 | 0.139 | 0.357 | 1 | Mac_UD |
| RPL14 | 0.42703675 | 0.92 | 0.978 | 1 | Mac_UD |
| SRI | 0.42653719 | 0.299 | 0.625 | 1 | Mac_UD |
| ATP6V1E1 | 0.42169619 | 0.247 | 0.545 | 1 | Mac_UD |
| RPL7A | 0.42155088 | 0.889 | 0.964 | 1 | Mac_UD |
| RPL22 | 0.42152109 | 0.851 | 0.967 | 1 | Mac_UD |
| EEF1A1 | 0.42055088 | 0.993 | 0.998 | 0.73319961 | Mac_UD |
| PCBD1 | 0.42002058 | 0.233 | 0.508 | 1 | Mac_UD |

| | | | | | |
|---|---|---|---|---|---|
| APEX1 | 0.41978631 | 0.222 | 0.496 | 0.4541404 | Mac_UD |
| TCP1 | 0.41676995 | 0.208 | 0.467 | 1 | Mac_UD |
| RPL38 | 0.41637928 | 0.885 | 0.973 | 0.99053164 | Mac_UD |
| TRAPPC2L | 0.41520776 | 0.233 | 0.509 | 1 | Mac_UD |
| RPAIN | 0.4130062 | 0.073 | 0.311 | 0.02915619 | Mac_UD |
