## Supplementary material for "Type I IFN-activated lung monocytes and macrophages as initiators and drivers of fibrosis at the alveolar barrier in IPF": Suppl Table 4B

**FABP4+ AM**

**Suppl Table 4B.** Annotation of myeloid cell subsets in lung digest from Morse's single cell dataset. Each re-clustered myeloid population was manually compared to macrophage populations in published datasets with similar transcriptional profiles, cluster annotation or proposed functional state. Top 50 cluster defining genes (ranked by Log2 fold change) for re-clustered Morse populations is shown and compared to top 20 cluster defining genes from relevant published datasets. Mac_UD populations are excluded from comparative analysis. UMAP of reclustered myeloid population is shown.

| Dataset | Oxford recluster and annotation | | | Morell 2023 - BAL (ARDS) | Morse 2021 - Lung Digest IPF and Control (2) | Mulder 2021 - Lung Digest plus mulitple tissues[a] | Zhang 2021 - BAL, Lung Digest plus multiple tissues[b] | Aegerter 2023 - Human myeloid scRNA seq key markers | Sikkema 2023 - Lung Cell Atlas |
|---|---|---|---|---|---|---|---|---|---|
| Annotation | FABP4+ AM | | | Mature AM | Cluster 0 (FABP4$^{hi}$)[c] | Cluster 16 MoMac (Alveolar Macs)[c] | MRC1$^{+}$ FABP4$^{+}$ | AMs | Alveolar macrophages_marker |
| | FABP4 | C1QC | STOM | FABP4 | FABP4 | C1QC | APOC1 | FABP4 | CYP27A1 |
| | SERPING1 | PLA2G16 | PEBP1 | IFI27 | C1QB | C1QB | C1QB | SERPING1 | MARCO |
| | INHBA | PCOLCE2 | LPL | SERPING1 | C1QA | APOE | C1QA | APOC1 | FABP4 |
| | C1QB | ALOX5AP | AKR1C2 | C1QB | FBP1 | FUCA1 | APOE | CD52 | |
| | CD52 | ALDH2 | EVL | RBP4 | C1QC | C1QA | FABP4 | PCOLCE2 | |
| | RP11-598F7.3 | FHL1 | CTSC | CCL18 | CD52 | DNASE1L3 | HLA-DQA1 | INHBA | |
| | AKR1C3 | AQP3 | ACOT7 | C1QA | MCEMP1 | HLA-DPA1 | HLA-DPB1 | MME (CD10) | |
| | RND3 | UBB | ALOX5 | CES1 | APOC1 | ACP5 | ALOX5AP | CES1 | |
| | C1QA | RP5-839B4.8 | DOK2 | NUPR1 | INHBA | RNASE1 | HLA-DPA1 | GPD1 | |
| | SERPINA1 | TREM1 | PPARG | C1QC | RP11---598F7.3 | HLA-DRB1 | HLA-DQB1 | RBP4 | |
| | RBP4 | LDHB | | FN1 | ALOX5AP | CD74 | ACP5 | ITIH5 | |
| | IFI27 | PPIC | | AC026369.3 | SERPINA1 | HLA-DQA1 | GCHFR | SCD | |
| | FBP1 | MT1E | | LGALS3BP | VSIG4 | PLD3 | UBB | | |
| | CES1 | HPGD | | AC009093.3 | MSR1 | CD63 | HLA-DRB1 | | |
| | LY6E | GPD1 | | INHBA | GCHFR | CTSC | NUPR1 | | |
| | LGALS3BP | FAM89A | | APOC1 | FTL | HLA-DRB5 | HLA-DRA | | |
| | MCEMP1 | GLDN | | PHLDA3 | GRN | HLA-DQA2 | MARCO | | |
| | PDLIM1 | MIR3945HG | | PDLIM1 | ACP5 | PTGDS | TMSB4X | | |
| | PHLDA3 | CITED2 | | TFRC (CD71) | TREM1 | HLA-DRA | HLA-DMA | | |
| | FABP3 | S100A13 | | ALDH2 | CD68 | CCL18 | FBP1 | | |
| Proportion of cluster defining genes in specified reference dataset that are also present in Oxford Morse's top 50 cluster defining genes | | | | 0.75 | 0.6 | 0.2 | 0.35 | 0.75 | 0.33 |

**Intermediate AM_1 and 2**

| Dataset | Oxford recluster and annotation | | | Morell 2023 - BAL (ARDS) |
|---|---|---|---|---|
| Annotation | Intermediate AM_1 | | | Intermediate AM |
| | MARCO | FCGRT | ATP6V0D1 | APOC1 |
| | RETN | QPCT | CD63 | GCHFR |
| | ACP5 | CXCL3 | HLA-B | ACP5 |
| | VSIG4 | CTSD | PPARG | FBP1 |
| | CCL20 | IL3RA | FXYD5 | APOE |
| | CYP27A1 | HLA-C | ABHD5 | VSIG4 |
| | GRN | MGST3 | GSTO1 | C1QA |
| | AGRP | MCEMP1 | EMP3 | AGRP |
| | APOE | ALOX5AP | ALDH2 | CYP27A1 |
| | LTA4H | CD164 | SLC11A1 | MARCO |
| | ANXA1 | SLC3A2 | | LTA4H |
| | CD68 | CD14 | | TREM2 |
| | HEXB | S100P | | AC009093.3 |
| | LRPAP1 | RNH1 | | ALDH2 |
| | LGALS3 | MYDGF | | C1QC |
| | FBP1 | S100A11 | | C1QB |
| | CD9 | HEXA | | UBB |
| | HLA-A | SLC31A2 | | MGST3 |
| | CTSA | ARL4A | | CD68 |
| | PLIN2 | ANXA2 | | GRN |
| Proportion of cluster defining genes in specified reference dataset that are also present in Oxford Morse's top 50 cluster defining genes | | | | 0.6 |

| Dataset | Oxford recluster and annotation | | | Morell 2023 - BAL (ARDS) |
|---|---|---|---|---|
| Annotation | Intermediate AM_2 | | | Intermediate AM |
| | NUPR1 | PDLIM1 | FAM89A | APOC1 |
| | CCL18 | SERPING1 | PPIC | GCHFR |
| | IFI27 | ALDH2 | PCOLCE2 | ACP5 |
| | FABP4 | PHLDA3 | C9orf16 | FBP1 |
| | RBP4 | RMDN3 | GLUL | APOE |
| | SCD | C1QB | CYP27A1 | VSIG4 |
| | GCHFR | ACOT7 | DNAJC5B | C1QA |
| | RP11-598F7.3 | AKR1C3 | MME | AGRP |
| | APOC1 | HPGD | PTPMT1 | CYP27A1 |
| | FTL | GLRX | C1QA | MARCO |
| | CES1 | UBB | | LTA4H |
| | CYB5A | AKR1B1 | | TREM2 |
| | LGALS3 | ADTRP | | AC009093.3 |
| | MSR1 | SCCPDH | | ALDH2 |
| | CSTB | FTH1 | | C1QC |
| | APOE | BLOC1S2 | | C1QB |
| | RND3 | ABCG1 | | UBB |
| | PLA2G16 | TCEB2 | | MGST3 |
| | TXN | LDHB | | CD68 |
| | PPDPF | S100A13 | | GRN |
| Proportion of cluster defining genes in specified reference dataset that are also present in Oxford Morse's top 50 cluster defining genes | | | | 0.4 |

**CXCL10$^{+}$ Mac**

| Dataset | Oxford recluster and annotation | | | Morell 2023 - BAL (ARDS) | Mould 2021 - BAL | Mulder 2021 - Lung Digest plus mulitple tissues[a] | | Zhang 2021 - BAL, Lung Digest plus multiple tissues[b] |
|---|---|---|---|---|---|---|---|---|
| Annotation | CXCL10$^{+}$ Mac | | | IFN-related | Cluster 8 | Cluster 6 MoMac (IFI44L Macs)[c] | Cluster 4 MoMac (ISG Mono)[c] | CXCL10$^{+}$ CCL2$^{+}$ |
| | ISG15 | OASL | RNF213 | CXCL10 | ISG15 | CXCL10 | CXCL10 | CCL2 |
| | CXCL10 | OAS2 | IRF7 | CCL8 | IFIT1 | CXCL9 | ISG15 | CXCL10 |
| | IFIT1 | HERC5 | ISG20 | ISG15 | IFIT2 | GBP1 | IFIT1 | IFITM3 |
| | IFIT3 | USP18 | PLSCR1 | IFIT2 | CXCL10 | IDO1 | CCL8 | ISG15 |
| | IFIT2 | GBP1 | MX2 | MX1 | RSAD2 | CXCL11 | IFIT2 | CCL8 |
| | MX1 | CCL8 | CCL7 | IFIT1 | TNFSF10 | STAT1 | IFIT3 | IL1RN |
| | NT5C3A | EIF2AK2 | XAF1 | IFIT3 | MX1 | IL4I1 | RSAD2 | CTSL |
| | IFI6 | IFI35 | PARP14 | RSAD2 | IFIT3 | GBP5 | TNFSF10 | IFITM1 |
| | TNFSF10 | LAP3 | GMPR | TNFSF10 | HERC5 | WARS | ISG20 | CCL3 |
| | IFITM3 | DYNLT1 | BST2 | IFITM1 | NEXN | MMP9 | CCL2 | RSAD2 |
| | LY6E | IFI44 | | IFITM3 | IFITM3 | CD40 | IFI44L | IFI27 |
| | RSAD2 | CCL2 | | GBP1 | NT5C3A | PPA1 | APOBEC3A | APOBEC3A |
| | EPSTI1 | STAT1 | | HERC5 | IFI44L | SLAMF7 | IFITM3 | ISG20 |
| | TNFSF13B | PSMB9 | | RNF213 | DDX58 | ANKRD22 | IFI6 | IFIT3 |
| | OAS1 | RABGAP1L | | MX2 | EPSTI1 | SOD2 | GBP1 | CCL4 |
| | IFI44L | MNDA | | APOBEC3A | CMPK2 | PTGDS | CXCL11 | MX1 |
| | CMPK2 | DEFB1 | | EPSTI1 | IFIH1 | VAMP5 | HERC5 | SOD2 |
| | SAMD9L | NEXN | | TNFSF13B | OAS2 | TAP1 | LY6E | IFIT2 |
| | UBE2L6 | OAS3 | | IRF7 | ISG20 | CCL8 | IL1RN | IFIT1 |
| | NUPR1 | IFI27 | | OAS1 | MX2 | PARP | IFI44 | GBP1 |
| Proportion of cluster defining genes in specified reference dataset that are also present in Oxford Morse's top 50 cluster defining genes | | | | 0.95 | 0.85 | 0.2 | 0.85 | 0.75 |

**SPP1 mid -hi mac**

| Dataset | Oxford recluster and annotation | Morell 2023 - BAL (ARDS) | Morse 2021 - Lung Digest IPF and Control | Mulder 2021 - Lung Digest plus mulitple tissues[a] | Aegerter 2023 - Human myeloid scRNA seq key markers |
|---|---|---|---|---|---|

| Dataset | Oxford recluster and annotation | Morell 2023 - BAL (ARDS) | Morse 2021 - Lung Digest IPF and Control | Mulder 2021 - Lung Digest plus mulitple tissues[a] | Aegerter 2023 - Human myeloid scRNA seq key markers |
|---|---|---|---|---|---|

**Footnote**

a Spleen (2), Lung (7), Liver (7), Skin (5), Blood (2), Lymph Node (1), Kidney (3), Head and Neck (1), Tonsil (2), Colon (6), Stomach (1), Ascites (1), Breast (2), Pancreas (1)

b Synovial Tissue(2), Kidney (1), Colon (1), Lung Digest (1), BAL (1)

c Annotation used in text

BAL Bronchoalveolar Lavage Cells

Overlapping genes

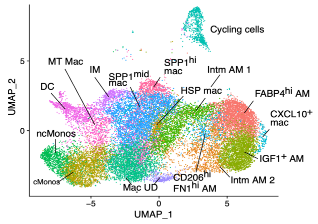

| Annotation | SPP1hi Mac | | | Matricellular | Cluster 1 (SPP1$^{hi}$)[b] | Cluster 3 MoMac (TREM2)[b] | IM (Subtype 2) |
|---|---|---|---|---|---|---|---|
| | CHI3L1 | SPARC | APOC1 | SPP1 | SPP1 | APOC1 | LGMN |
| | MMP7 | FABP5 | ITGB1BP1 | CHIT1 | APOE | APOE | MARCKS |
| | SPP1 | CD9 | BCAP31 | LPL | CCL18 | ACP5 | SPP1 |
| | CHIT1 | GM2A | CD63 | CD9 | CTSB | SPP1 | PLA2G7 |
| | CTSK | TM4SF19 | GPC4 | CHI3L1 | GPNMB | CTSD | MMP9 |
| | MMP9 | LPL | MGLL | MMP7 | CTSL | GPNMB | HAMP |
| | SDC2 | PLD3 | CD36 | GSN | CTSD | FABP5 | |
| | TIMP3 | RP11-20G13.3 | TDRD3 | TREM2 | LGMN | LGALS3 | |
| | LIPA | CSTB | FAIM | LIPA | FTL | CTSB | |
| | IGLC2 | ARID5B | HEXB | MATK | APOC1 | PLA2G7 | |
| | FDX1 | LGMN | | LINC02345 | CSTB | CD63 | |
| | CALM3 | GSN | | FABP3 | CTSZ | CD9 | |
| | GPNMB | RGCC | | FBP1 | FABP5 | PLD3 | |
| | APOE | CAPG | | SPARC | PSAP | LIPA | |
| | LILRB4 | DNASE2B | | CAMK1 | MARCO | LGMN | |
| | MATK | MFSD12 | | SDC2 | CD68 | TREM2 | |
| | TREM2 | HCST | | A2M | GPX1 | MMP9 | |
| | CIR1 | ITGB2 | | GPNMB | ACP5 | SDS | |
| | PLA2G7 | CYP27A1 | | PLA2G7 | MS4A4A | C1QB | |
| | RARRES1 | FNIP2 | | MFSD12 | LGALS1 | CCL18 | |
| Proportion of cluster defining genes in specified reference dataset that are also present in Oxford Morse's top 50 cluster defining genes | | | | 0.75 | 0.4 | 0.65 | 0.83 |

| Annotation | SPP1 mid Macs | | | Matricellular | LGMN/CD163 | Cluster 1 (SPP1$^{hi}$)[b] | Cluster 3 MoMac (TREM2)[b] | IM (Subtype 2) |
|---|---|---|---|---|---|---|---|---|
| | LGMN | HS3ST2 | TSPAN4 | SPP1 | RNASE1 | SPP1 | APOC1 | LGMN |
| | APOE | PLTP | SDS | CHIT1 | LGMN | APOE | APOE | MARCKS |
| | CCL18 | CD84 | TMIGD3 | LPL | CCL13 | CCL18 | ACP5 | SPP1 |
| | GPNMB | PLD3 | FPR3 | CD9 | HMOX1 | CTSB | SPP1 | PLA2G7 |
| | CTSZ | TREM2 | FOSB | CHI3L1 | CCL2 | GPNMB | CTSD | MMP9 |
| | RNASE1 | PMP22 | ZFAND5 | MMP7 | CTSL | CTSL | GPNMB | HAMP |
| | CTSB | NPC2 | SMPDL3A | GSN | PLTP | CTSD | FABP5 | |
| | PLA2G7 | DAB2 | FOS | TREM2 | SPP1 | LGMN | LGALS3 | |
| | SPP1 | LGALS1 | MGLL | LIPA | CD163 | FTL | CTSB | |
| | HMOX1 | RGS1 | HAMP | MATK | FOLR2 | APOC1 | PLA2G7 | |
| | CTSL | FTL | | LINC02345 | MARCKS | CSTB | CD63 | |
| | CCL13 | CD59 | | FABP3 | STAB1 | CTSZ | CD9 | |
| | SEPP1 | TGFBI | | FBP1 | CTSB | FABP5 | PLD3 | |
| | CPM | SGK1 | | SPARC | F13A1 | PSAP | LIPA | |
| | CTSD | ARL4C | | CAMK1 | CTSD | MARCO | LGMN | |
| | APOC1 | CREG1 | | SDC2 | HSPA5 | CD68 | TREM2 | |
| | LIPA | NPL | | A2M | GLUL | GPX1 | MMP9 | |
| | CCL2 | ABCA1 | | GPNMB | TGFBI | ACP5 | SDS | |
| | MARCKS | CD36 | | PLA2G7 | PLA2G7 | MS4A4A | C1QB | |
| | PSAP | MS4A4A | | MFSD12 | CALR | LGALS1 | CCL18 | |
| Proportion of cluster defining genes in specified reference dataset that are also present in Oxford Morse's top 50 cluster defining genes | | | | 0.25 | 0.6 | 0.75 | 0.6 | 0.83 |

**IM**

| Dataset | Oxford recluster and annotation | | | Morell 2023 - BAL (ARDS) | Mulder 2021 - Lung Digest plus mulitple tissues[a] | Aegerter 2023 - Human myeloid scRNA seq key markers | Sikkema 2023 - Lung Cell Atlas |
|---|---|---|---|---|---|---|---|
| Annotation | IM | | | LGMN/CD163 | Cluster 2 MoMac (HES1)[b] | IM (Subtype 1) | Interstitial Mph perivascular_marker |
| | SEPP1 | HLA-DPA1 | A2M | RNASE1 | SLC40A1 | LGMN | F13A1 |
| | CCL13 | LGMN | GPR183 | LGMN | RNASE1 | MARCKS | FOLR2 |
| | FOLR2 | HLA-DPB1 | FOSB | CCL13 | LGMN | FOLR2 | |
| | CCL4L2 | HLA-DRA | HSPA1B | HMOX1 | C1QA | SELENOP | |
| | CCL4 | RGS2 | CEBPD | CCL2 | C1QB | F13A1 | |
| | RNASE1 | CD14 | RBPJ | CTSL | STAB1 | SLC40A1 | |
| | SLC40A1 | TMEM176A | CH25H | PLTP | DAB2 | | |
| | F13A1 | HLA-DRB1 | CTSC | SPP1 | F13A1 | | |
| | CCL3L3 | ZFP36 | MARCKS | CD163 | A2M | | |
| | RGS1 | FGL2 | TGFBI | FOLR2 | MAF | | |
| | CCL3 | STAB1 | | MARCKS | HES1 | | |
| | MS4A6A | FCGR2A | | STAB1 | FUCA1 | | |
| | PLTP | C1QC | | CTSB | IGF1 | | |
| | HLA-DQA1 | RNASE6 | | F13A1 | EGR1 | | |
| | GPR34 | CFD | | CTSD | HSPA1A | | |
| | FOS | KLF6 | | HSPA5 | C1QC | | |
| | PDK4 | ITM2B | | GLUL | CCL3 | | |
| | EGR1 | HLA-DQB1 | | TGFBI | NR4A2 | | |
| | TMEM176B | CD74 | | PLA2G7 | CCL4 | | |
| | HSPA1A | MS4A4A | | CALR | CTSC | | |
| Proportion of cluster defining genes in specified reference dataset that are also present in Oxford Morse's top 50 cluster defining genes | | | | 0.45 | 0.6 | 0.83 | 0.2 |

**HSP Mac**

| Dataset | Oxford recluster and annotation | | | Mulder 2021 - Lung Digest plus mulitple tissues[a] |
|---|---|---|---|---|
| Annotation | HSP Mac | | | Cluster 12 MoMac (HSP)[b] |
| | HSPA1B | CCL2 | KLF4 | HSPA1B |
| | HSPA1A | KLF2 | DNAJB6 | HSPH1 |
| | DNAJB1 | ATF3 | CACYBP | HSPA6 |
| | HMOX1 | APOE | TSC22D3 | DNAJB1 |
| | HSPA6 | PLIN2 | WBP5 | HSPA1A |
| | HSPH1 | JUND | CHORDC1 | BAG3 |
| | HSP90AA1 | EGR1 | MGST1 | DNAJA4 |
| | ZFAND2A | DNAJA1 | TXN | ZFAND2A |
| | JUN | RHOB | DUSP1 | HSP90AA1 |
| | HSPB1 | SQSTM1 | MRPL18 | HSPB1 |
| | BAG3 | UBC | | HSPE1 |
| | HSPE1 | HSPA8 | | HSPD1 |
| | FOS | HSPA5 | | IER5 |
| | IER5 | GADD45B | | FKBP4 |
| | ANXA1 | ADM | | TNFSF14 |
| | KLF6 | PDK4 | | G0S2 |
| | DNAJA4 | NEAT1 | | APOBEC3A |
| | HSP90AB1 | RNASE1 | | CACYBP |
| | CCL18 | DNAJB4 | | NR4A1 |
| | HSPD1 | FOSB | | CHORDC1 |
| Proportion of cluster defining genes in specified reference dataset that are also present in Oxford Morse's top 50 cluster defining genes | | | | 0.75 |

**MT Mac**

| Dataset | Oxford recluster and annotation | | | Morell 2023 - BAL (ARDS) | Mulder 2021 - Lung Digest plus mulitple tissues[a] | Sikkema 2023 - Lung Cell Atlas |
|---|---|---|---|---|---|---|
| Annotation | MT Mac | | | Metallothionein | Cluster 11 MoMac (MT)[b] | Alveolar Mph MT-positive_marker |
| | MT1X | RP11-1008C21.1 | BTG1 | MT1G | MT1G | MT1M |
| | SPP1 | LPL | QPCT | MT2A | MT1X | CCL18 |
| | MT1H | SH3BGRL3 | CALM1 | MT1X | MT1E | MT1E |
| | MT1E | LSP1 | NME1 | MT1H | MT2A | |
| | C15orf48 | EMP3 | BASP1 | MT1F | MT1M | |
| | MT2A | ID3 | CTSH | MT1E | MT1F | |
| | EMP1 | LMNA | LGALS1 | MT1M | APOC1 | |
| | AREG | CAMK1 | SGK1 | MT1L | CTSD | |
| | SDC2 | LIMS1 | RHOC | TPT1 | LGMN | |
| | CLEC5A | SMIM3 | LILRB4 | RPS26 | CTSB | |
| | IL1RN | MT1F | | MIF | GLUL | |

| | | | | | |
|---|---|---|---|---|---|
| | ARID5B | SDS | RGCC | CXCL1 | |
| | HCST | HMGA1 | OLR1 | SDS | |
| | GPR183 | RGCC | THBS1 | FUCA1 | |
| | S100A10 | GSN | HCST | FTL | |
| | MATK | TYMP | IL1B | LGALS3 | |
| | ALCAM | NEAT1 | RPL13P12 | GPNMB | |
| | TREM2 | MRC1 | BTG1 | ABCA1 | |
| | IL3RA | HERPUD1 | ACP5 | HMOX1 | |
| | FCGR2B | SLC16A10 | ZFP36 | IL6 | |
| Proportion of cluster defining genes in specified reference dataset that are also present in Oxford Morse's top 50 cluster defining genes | | | 0.5 | 0.35 | 0.67 |

**cMono**

| Dataset | Oxford recluster and annotation | | | Morse 2021 - Lung Digest Control Only | Mulder 2021 - Lung Digest plus mulitple tissues[a] | Zhang 2021 - BAL, Lung Digest plus multiple tissues[b] | Aegerter 2023 - Human myeloid scRNA seq key markers | Sikkema 2023 - Lung Cell Atlas |
|---|---|---|---|---|---|---|---|---|
| Annotation | cMono | | | FCN1 Mono[c] | Cluster 21 MoMac (cMono)[c] | FCN1+ | recMac | Classical monocytes_marker |
| | S100A8 | IER3 | BTG1 | S100A8 | S100A8 | S100A9 | S100A8 | S100A12 |
| | S100A9 | PTGS2 | VEGFA | S100A9 | S100A9 | FCN1 | FN1 | FCN1 |
| | IL1B | AREG | TNFAIP3 | SERPINB2 | S100A12 | S100A12 | VCAN | RNASE2 |
| | S100A12 | CD93 | CYP1B1 | THBS1 | VCAN | VCAN | RNASE1 | |
| | SERPINB2 | CCL3 | RILPL2 | S100A12 | FCN1 | S100A4 | FCN1 | |
| | G0S2 | CD300E | ATP2B1 | IL1B | CSTA | IFITM2 | EMP1 | |
| | CXCL8 | SAMSN1 | NFKBIA | EREG | RETN | APOBEC3A | CD14 | |
| | THBS1 | PPIF | C5AR1 | FCN1 | S100A4 | TIMP1 | | |
| | TIMP1 | CCL3L3 | IFITM2 | G0S2 | FPR1 | FOS | | |
| | EREG | SRGN | TNFRSF1B | VCAN | CFP | CORO1A | | |
| | VCAN | CXCL1 | | TIMP1 | SELL | SRGN | | |
| | FCN1 | CXCL3 | | NAMPT | TSPO | LYZ | | |
| | SOD2 | SLC25A37 | | PLAUR | CSF3R | H3F3B | | |
| | NAMPT | CTB-61M7.2 | | CD300E | MNDA | FPR1 | | |
| | CXCL2 | CREM | | RP11---1143G9.4 | IER2 | NAMPT | | |
| | CCL20 | C15orf48 | | IL1R2 | AP1S2 | COTL1 | | |
| | PLAUR | BCL2A1 | | AIF1 | RBP7 | PLAC8 | | |
| | SLC2A3 | SERPINB9 | | C5AR1 | CD52 | CFP | | |
| | IL1R2 | RP11-1143G9.4 | | LST1 | CD36 | ACTB | | |
| | NLRP3 | MXD1 | | PPIF | STXBP2 | IFITM3 | | |
| Proportion of cluster defining genes in specified reference dataset that are also present in Oxford Morse's top 50 cluster defining genes | | | | 0.9 | 0.3 | 0.4 | 0.35 | 0.67 |

| Dataset | Oxford recluster and annotation | | | Morse 2021 - Lung Digest Control Only | Zhang 2021 - BAL, Lung Digest plus multiple tissues[b] |
|---|---|---|---|---|---|
| Annotation | ncMono | | | FCN1 Mono[c] | FCN1+ |
| | IFITM2 | MT2A | COTL1 | S100A8 | S100A9 |
| | FCN1 | FAM26F | EREG | S100A9 | FCN1 |
| | IFITM3 | CD300E | C5AR1 | SERPINB2 | S100A12 |
| | CXCL10 | SRGN | SLC2A3 | THBS1 | VCAN |
| | TIMP1 | BIRC3 | RNF144B | S100A12 | S100A4 |
| | NAMPT | VAMP5 | CLEC4E | IL1B | IFITM2 |
| | THBS1 | SOCS3 | INSIG1 | EREG | APOBEC3A |
| | FPR2 | DDIT4 | HMGB2 | FCN1 | TIMP1 |
| | APOBEC3A | PLAC8 | ADM | G0S2 | FOS |
| | G0S2 | DUSP2 | LILRA1 | VCAN | CORO1A |
| | CFP | SERPINB9 | | TIMP1 | SRGN |
| | FGL2 | LILRA5 | | NAMPT | LYZ |
| | RGS2 | SAT1 | | PLAUR | H3F3B |
| | CD48 | AIF1 | | CD300E | FPR1 |
| | LST1 | CEBPD | | RP11---1143G9.4 | NAMPT |
| | CORO1A | FPR1 | | IL1R2 | COTL1 |
| | HES4 | SAMSN1 | | AIF1 | PLAC8 |
| | WARS | LILRB2 | | C5AR1 | CFP |
| | ISG20 | C10orf54 | | LST1 | ACTB |
| | SOD2 | CREM | | PPIF | IFITM3 |
| Proportion of cluster defining genes in specified reference dataset that are also present in Oxford Morse's top 50 | | | | 0.5 | 0.65 |

**DC**

| Dataset | Oxford recluster and annotation | | | Morell 2023 - BAL (ARDS ) | Mulder 2021 - Lung Digest plus mulitple tissues[a] | | Aegerter 2023 - Human myeloid scRNA seq key markers | | Sikkema 2023 - Lung Cell Atlas | |
|---|---|---|---|---|---|---|---|---|---|---|
| Annotation | DC | | | DCs | Cluster 3 MNP (cDC1)[b] | Cluster 15 MNP (cDC2/DC3)[b] | cDC1 | cDC2 | DC1_marker | DC2_marker |
| | CCL17 | REL | SERPINB9 | CCL17 | CPVL | FCER1A | CLEC3A | CD1C | XCR1 | PKIB |
| | GPR183 | PKIB | RAMP1 | CD1C | IRF8 | CLEC10A | XCR1 | CD1E | CLEC9A | CLEC10A |
| | RGS1 | CEBPD | MALAT1 | WFDC21P | DNASE1L3 | CD1C | CADM1 | CLEC10A | CLNK | CD1E |
| | S100B | HLA-DRB5 | AXL | S100B | SNX3 | AREG | BTLA | FCER1A | | |
| | HLA-DPB1 | HLA-DRA | CSF2RA | CST7 | CST3 | GPR183 | IDO1 | FCR2B | | |
| | CXCR4 | ZFP36 | CST3 | HLA-DQA1 | IDO1 | CD1E | DNASEIL3 | CC17 | | |
| | CST7 | CCR7 | PPP1R14A | RGS1 | LGALS2 | PKIB | CCND1 | PKIB | | |
| | HLA-DQA1 | HLA-DRB1 | SPINT2 | IFITM1 | ID2 | NDRG2 | S100B | LGALS2 | | |
| | AREG | CREM | CD1E | NAPSB | CPNE3 | CD74 | LGALS2 | BIRC3 | | |
| | INSIG1 | CD1C | CORO1A | HLA-DPB1 | TAP1 | CXCR4 | | | | |
| | HLA-DPA1 | HLA-DQA2 | | GPR183 | NAAA | PLD4 | | | | |
| | HLA-DQB1 | IL1R2 | | HLA-DQB1 | WDFY4 | PPA1 | | | | |
| | FCER1A | RNASE6 | | HLA-DPA1 | CD74 | EIF3L | | | | |
| | CD74 | CD83 | | CD1E | SERPINF1 | CORO1A | | | | |
| | HERPUD1 | FCGR2B | | PPP1R14A | TACSTD2 | ZFP36 | | | | |
| | DUSP4 | PPA1 | | FCER1A | CCND1 | CST3 | | | | |
| | RGS2 | MS4A6A | | CLEC10A | PPA1 | LGALS2 | | | | |
| | BIRC3 | BTG1 | | ARL4C | CADM1 | ALDH2 | | | | |
| | CLEC10A | LTB | | CD74 | EEF1B2 | RPL4 | | | | |
| | CCL22 | DDIT4 | | PKIB | GSTP1 | PHACTR1 | | | | |
| Proportion of cluster defining genes in specified reference dataset that are also present in Oxford Morse's top 50 cluster defining genes | | | | 0.70 | 0.15 | 0.70 | 0.11 | 0.78 | 0.00 | 1.00 |

**Cycling**

| Dataset | Oxford recluster and annotation | | | Mulder 2021 - Lung Digest plus mulitple tissues[a] | Sikkema 2023 - Lung Cell Atlas |
|---|---|---|---|---|---|
| Annotation | Cycling | | | Cluster 11 MNP (Proliferating)[b] | Alveolar Mph proliferating |
| | STMN1 | DEK | SNRNP25 | STMN1 | TYMS |
| | KIAA0101 | CDKN3 | SKA2 | HIST1H4C | CENPW |
| | H2AFZ | TMEM106C | STRA13 | TUBB | MND1 |
| | HIST1H4C | BIRC5 | HMGB3 | H2AFZ | |
| | TUBB | TOP2A | RAD51AP1 | UBE2C | |
| | TUBA1B | MKI67 | SIVA1 | TUBA1B | |
| | HMGN2 | NUCKS1 | SMC2 | TOP2A | |
| | UBE2C | SMC4 | MZT2B | HMGB2 | |
| | PTTG1 | FN1 | RAN | TK1 | |
| | TYMS | CENPW | HMGN1 | HMGN2 | |
| | IGKC | IDH2 | | PTTG1 | |
| | CKS1B | NUSAP1 | | CDK1 | |
| | TK1 | ANP32B | | MKI67 | |
| | HMGB1 | CKS2 | | TYMS | |
| | HMGB2 | RPA3 | | HMGB1 | |
| | RRM2 | CENPM | | PCNA | |
| | PCNA | CARHSP1 | | RRM2 | |
| | H2AFV | RANBP1 | | BIRC5 | |
| | CDK1 | CENPF | | DUT | |
| | DUT | DTYMK | | NUSAP1 | |
| Proportion of cluster defining genes in specified reference dataset that are also present in Oxford Morse's top 50 cluster defining genes | | | | 1.00 | 0.66 |
