## Supplementary material for "Type I IFN-activated lung monocytes and macrophages as initiators and drivers of fibrosis at the alveolar barrier in IPF": Suppl Table 5A

**Supplementary Table 5A** . Full list of significantly differentially regulated pathways in IPF BAL AMs compared to control samples (Oxford Cohort A). Gene set enrichment analysis using Gene Ontology (GO) Hallmark database ranked by NES. ES - enrichment score. NES - normalised enrichment score – enrichment score. Only those AMs with statitiscally significant NES are shown (adj p < 0.05;

**IGF1+ AM**

| pathway | pval | padj | ES | NES | nMoreExtreme | size | leadingEdge |
|---|---|---|---|---|---|---|---|
| HALLMARK_CHOLESTEROL_HOMEOSTASIS | 0.00126743 | 0.01584284 | 0.60062198 | 1.87586306 | 0 | 65 | FADS2 ATF3 STARD4 TM7SF2 LDLR DHCR7 HMGCS1 LGMN SQLE CHKA FASN EBP TMEM97 CTNNB1 MVD ACAT2 IDI1 TRIB3 NFIL3 FDPS HMGCR LSS MVK ALCAM LPL CPEB2 CYP51A1 SREBF2 |
| HALLMARK_IL6_JAK_STAT3_SIGNALING | 0.00124688 | 0.01584284 | 0.51978481 | 1.65113857 | 0 | 73 | CXCL9 CXCL10 CXCL11 IL6 SOCS3 PF4 IL7 IL1B CXCL3 PDGFC ACVR1B A2M PIM1 IL2RA LEPR CXCL1 SOCS1 IL9R MAP3K8 |
| HALLMARK_KRAS_SIGNALING_UP | 0.00112233 | 0.01584284 | 0.45018955 | 1.55110855 | 0 | 133 | PCSK1N JUP CXCL10 SCG5 ADAMDEC1 CCL20 RGS16 TMEM176A GALNT3 TMEM176B GNG11 PLEK2 G0S2 EREG DCBLD2 TRIB2 MALL SATB1 TRIB1 IL1B CFH PTGS2 DUSP6 GPRC5B ANKH ATG10 HSD11B1 CCND2 TRAF1 |
| HALLMARK_TNFA_SIGNALING_VIA_NFKB | 0.00108578 | 0.01584284 | 0.48306388 | 1.70071649 | 0 | 179 | CXCL10 CXCL11 CCL20 IL6 ATF3 GCH1 AREG FJX1 SOCS3 NR4A3 MARCKS LDLR FOSB G0S2 NR4A1 IL1A CXCL2 PANX1 NR4A2 TRIB1 IL1B DUSP1 SDC4 PTGS2 EGR1 IL23A SLC2A3 TSC22D1 VEGFA ICAM1 CXCL3 NFIL3 ICOSLG DUSP5 KDM6B ABCA1 EGR2 CXCL1 PHLDA1 TRAF1 MAP3K8 CD69 [illegible] |
| HALLMARK_ALLOGRAFT_REJECTION | 0.01020408 | 0.10204082 | -0.3259108 | -1.4143693 | 0 | 152 | CD8B CD2 CD3E ETS1 CCL2 CD7 GZMB CCL5 CD8A LCK GZMA CD3D CD3G MAP4K1 TRAT1 LTB IL16 KLRD1 CD247 CD1D BCL3 CD79A SOCS5 |
| HALLMARK_INFLAMMATORY_RESPONSE | 0.02569832 | 0.17361111 | 0.40494443 | 1.40330971 | 22 | 145 | CXCL9 CXCL10 CXCL11 CCL20 RGS16 IL6 OSM GCH1 RASGRP1 CXCL8 KCNJ2 LDLR NLRP3 EREG DCBLD2 SCARF1 GABBR1 IL1A RGS1 IL1B RNF144B ICAM1 MSR1 ACVR1B ICOSLG ABCA1 LPAR1 LCP2 CD69 GPR183 AXL |
| HALLMARK_MTORC1_SIGNALING | 0.02494577 | 0.17361111 | 0.38651254 | 1.36600247 | 22 | 187 | FADS2 STARD4 TM7SF2 LDLR DHCR7 FADS1 HMGCS1 LGMN CTH SQLE RRM2 VLDLR EBP PSPH INSIG1 TMEM97 MLLT11 PHGDH CCNF SLC2A3 BCAT1 HK2 IDI1 TRIB3 NFIL3 GCLC DHCR24 HMGCR IFRD1 ACLY PSAT1 SRD5A1 PDK1 SLC1A4 ELOVL5 TBK1 GTF2H1 RPA1 MTHFD2L TFRC ME1 [illegible] |
| HALLMARK_SPERMATOGENESIS | 0.02777778 | 0.17361111 | 0.4685847 | 1.46713248 | 21 | 66 | PCSK1N SCG5 TTK CDK1 TNNI3 SLC12A2 OAZ3 CHFR ZC2HC1C HSPA2 CCNB2 DBF4 CDKN3 PGS1 RAD17 PIAS2 PARP2 LPIN1 CLPB |
| HALLMARK_G2M_CHECKPOINT | 0.05888768 | 0.32715376 | 0.37303401 | 1.30776853 | 53 | 166 | HMMR TTK MARCKS [illegible] |
| HALLMARK_INTERFERON_GAMMA_RESPONSE | 0.06956522 | 0.33413988 | 0.3598921 | 1.27150905 | 63 | 186 | CXCL9 CXCL10 [illegible] |
| HALLMARK_KRAS_SIGNALING_DN | 0.07351077 | 0.33413988 | 0.43318703 | 1.35293009 | 57 | 65 | NR6A1 GPRC5C [illegible] |
| HALLMARK_EPITHELIAL_MESENCHYMAL_TRANSITION | 0.08799049 | 0.36662703 | 0.39987185 | 1.32486498 | 73 | 100 | IL6 CXCL8 AREG [illegible] |
| HALLMARK_COAGULATION | 0.10121951 | 0.38930582 | 0.40183019 | 1.2980034 | 82 | 81 | MMP15 DUSP14 [illegible] |
| HALLMARK_APOPTOSIS | 0.13703284 | 0.48940301 | 0.35317461 | 1.21234469 | 120 | 131 | IL6 ATF3 F2R GCH1 [illegible] |
| HALLMARK_UV_RESPONSE_UP | 0.16018307 | 0.53394355 | 0.34871055 | 1.18875876 | 139 | 125 | IL6 ATF3 GCH1 [illegible] |
| HALLMARK_APICAL_SURFACE | 0.19772404 | 0.61788762 | 0.47442098 | 1.23917406 | 138 | 25 | CD160 PCSK9 [illegible] |
| HALLMARK_ANGIOGENESIS | 0.2301013 | 0.66642433 | 0.48503512 | 1.21968406 | 158 | 20 | PF4 FGFR1 VEGFA [illegible] |
| HALLMARK_IL2_STAT5_SIGNALING | 0.23991276 | 0.66642433 | 0.32151064 | 1.12713986 | 219 | 166 | CXCL10 ITGA6 [illegible] |
| HALLMARK_E2F_TARGETS | 0.25732899 | 0.67718155 | 0.31439651 | 1.10690531 | 236 | 178 | HMMR DIAPH3 [illegible] |
| HALLMARK_HEDGEHOG_SIGNALING | 0.30713246 | 0.74074074 | 0.4622024 | 1.15280176 | 210 | 19 | SLIT1 VLDLR NRP2 [illegible] |
| HALLMARK_MYOGENESIS | 0.31111111 | 0.74074074 | 0.32459828 | 1.08475876 | 265 | 107 | FST COL6A3 DMD [illegible] |
| HALLMARK_MITOTIC_SPINDLE | 0.35969664 | 0.786497 | 0.30024181 | 1.05698257 | 331 | 176 | TTK BCR MARCKS [illegible] |
| HALLMARK_REACTIVE_OXYGEN_SPECIES_PATHWAY | 0.36178862 | 0.786497 | -0.2998217 | -1.069411 | 88 | 48 | SRXN1 EGLN2 [illegible] |
| HALLMARK_COMPLEMENT | 0.37867247 | 0.78890098 | 0.29952119 | 1.04984172 | 347 | 162 | MMP15 IL6 [illegible] |
| HALLMARK_XENOBIOTIC_METABOLISM | 0.39569161 | 0.79138322 | 0.30188856 | 1.03849731 | 348 | 136 | JUP SMOX GCH1 [illegible] |
| HALLMARK_ANDROGEN_RESPONSE | 0.46218487 | 0.85589791 | 0.30946946 | 1.00424335 | 384 | 84 | NKX3-1 FADS1 [illegible] |
| HALLMARK_ESTROGEN_RESPONSE_EARLY | 0.46083995 | 0.85589791 | 0.29535543 | 1.01003211 | 405 | 128 | RASGRP1 AREG [illegible] |
| HALLMARK_WNT_BETA_CATENIN_SIGNALING | 0.50496454 | 0.90172239 | 0.3574963 | 0.97641879 | 355 | 31 | FZD8 NOTCH4 [illegible] |
| HALLMARK_BILE_ACID_METABOLISM | 0.56773399 | 0.94692631 | 0.29131506 | 0.92962472 | 460 | 75 | FADS2 FADS1 [illegible] |
| HALLMARK_ESTROGEN_RESPONSE_LATE | 0.56815578 | 0.94692631 | 0.28199634 | 0.95640048 | 495 | 121 | AREG DHCR7 [illegible] |
| HALLMARK_APICAL_JUNCTION | 0.6150289 | 0.9919821 | 0.27358933 | 0.92203577 | 531 | 112 | JUP CERCAM [illegible] |
| HALLMARK_ADIPOGENESIS | 0.89848812 | 1 | 0.21987062 | 0.77489409 | 831 | 174 | PHLDB1 C3 [illegible] |
| HALLMARK_DNA_REPAIR | 0.99106145 | 1 | 0.18352113 | 0.63598106 | 886 | 145 | TYMS ADCY6 [illegible] |
| HALLMARK_FATTY_ACID_METABOLISM | 0.74789916 | 1 | -0.21385 | -0.9099397 | 88 | 131 | AOC3 XIST ACOT2 [illegible] |
| HALLMARK_GLYCOLYSIS | 0.71428571 | 1 | 0.25395478 | 0.88360606 | 644 | 151 | HMMR CDK1 CTH [illegible] |
| HALLMARK_HEME_METABOLISM | 0.99553073 | 1 | 0.17324401 | 0.60036633 | 890 | 145 | SMOX SNCA TNS1 [illegible] |
| HALLMARK_HYPOXIA | 0.74161074 | 1 | 0.25084358 | 0.86879486 | 662 | 144 | IL6 ATF3 CAVIN3 [illegible] |
| HALLMARK_INTERFERON_ALPHA_RESPONSE | 0.66350711 | 1 | 0.2690953 | 0.88836792 | 559 | 95 | CXCL10 CXCL11 [illegible] |
| HALLMARK_MYC_TARGETS_V1 | 1 | 1 | 0.13881571 | 0.48997963 | 922 | 183 | TYMS U2AF1 [illegible] |
| HALLMARK_MYC_TARGETS_V2 | 0.76592978 | 1 | 0.26951107 | 0.82202699 | 588 | 57 | SLC29A2 TMEM97 [illegible] |
| HALLMARK_NOTCH_SIGNALING | 0.83898305 | 1 | 0.27276938 | 0.72004246 | 593 | 26 | FZD7 KAT2A FZD1 [illegible] |
| HALLMARK_OXIDATIVE_PHOSPHORYLATION | 1 | 1 | -0.1112164 | -0.5001239 | 62 | 200 | BCKDHA NDUFA7 [illegible] |
| HALLMARK_P53_PATHWAY | 0.8671024 | 1 | 0.22811002 | 0.80011011 | 795 | 165 | RGS16 ATF3 F2R [illegible] |
| HALLMARK_PANCREAS_BETA_CELLS | 0.87643678 | 1 | -0.2726424 | -0.6849172 | 304 | 12 | STXBP1 FOXO1 [illegible] |
| HALLMARK_PEROXISOME | 0.82360097 | 1 | 0.24654558 | 0.79714185 | 676 | 82 | FADS1 SLC25A4 [illegible] |
| HALLMARK_PI3K_AKT_MTOR_SIGNALING | 0.98793727 | 1 | 0.18380099 | 0.59807164 | 818 | 88 | PAK4 CDK1 [illegible] |
| HALLMARK_PROTEIN_SECRETION | 0.97941889 | 1 | 0.19868133 | 0.64641424 | 808 | 89 | TOM1L1 SSPN [illegible] |
| HALLMARK_TGF_BETA_SIGNALING | 0.96816976 | 1 | 0.21331684 | 0.63223631 | 729 | 47 | SPTBN1 CTNNB1 [illegible] |
| HALLMARK_UNFOLDED_PROTEIN_RESPONSE | 0.89800703 | 1 | 0.22593734 | 0.75369542 | 765 | 105 | ATF3 WFS1 VEGFA [illegible] |
| HALLMARK_UV_RESPONSE_DN | 0.8213866 | 1 | 0.24252521 | 0.80901094 | 698 | 106 | SYNJ2 LDLR [illegible] |

**CD206hiFN1hi AM**

| | | | | | | | |
|---|---|---|---|---|---|---|---|
| HALLMARK_ALLOGRAFT_REJECTION | 0.00171527 | 0.02487562 | -0.4900305 | -1.8769055 | 0 | 153 | CD79A DYRK3 [illegible] |
| HALLMARK_CHOLESTEROL_HOMEOSTASIS | 0.00220264 | 0.02487562 | 0.66507479 | 2.34405107 | 0 | 66 | TM7SF2 FASN [illegible] |
| HALLMARK_HYPOXIA | 0.00239808 | 0.02487562 | 0.37683791 | 1.50829798 | 0 | 146 | LDHC TPBG [illegible] |
| HALLMARK_MTORC1_SIGNALING | 0.00248756 | 0.02487562 | 0.38722312 | 1.59021186 | 0 | 185 | CTH TM7SF2 [illegible] |
| HALLMARK_TNFA_SIGNALING_VIA_NFKB | 0.00239808 | 0.02487562 | 0.42873269 | 1.76297645 | 0 | 178 | TNFAIP6 MARCKS [illegible] |
| HALLMARK_EPITHELIAL_MESENCHYMAL_TRANSITION | 0.01157407 | 0.08775009 | 0.39125207 | 1.47289333 | 4 | 99 | SPP1 COL1A1 [illegible] |
| HALLMARK_KRAS_SIGNALING_UP | 0.01228501 | 0.08775009 | 0.36391087 | 1.42240825 | 4 | 132 | PCSK1N SPP1 JUP [illegible] |
| HALLMARK_ANDROGEN_RESPONSE | 0.01605505 | 0.10034404 | 0.41327737 | 1.51811273 | 6 | 85 | AKAP12 CDC14B [illegible] |
| HALLMARK_MITOTIC_SPINDLE | 0.0383693 | 0.2131628 | 0.31851589 | 1.30975789 | 15 | 178 | BCR MARCKS [illegible] |
| HALLMARK_UV_RESPONSE_DN | 0.09601874 | 0.48009368 | 0.32136939 | 1.22650965 | 40 | 106 | COL1A1 NR1D2 [illegible] |
| HALLMARK_ESTROGEN_RESPONSE_EARLY | 0.12837838 | 0.58353808 | -0.3148919 | -1.1867477 | 75 | 131 | MYBL1 MSMB [illegible] |
| HALLMARK_SPERMATOGENESIS | 0.16411379 | 0.66498922 | 0.3322635 | 1.17888283 | 74 | 68 | PCSK1N LDHC [illegible] |
| HALLMARK_XENOBIOTIC_METABOLISM | 0.1728972 | 0.66498922 | 0.28924696 | 1.15023416 | 73 | 136 | JUP TMEM176B [illegible] |
| HALLMARK_ANGIOGENESIS | 0.28571429 | 0.78477444 | 0.4158104 | 1.13977829 | 137 | 20 | SPP1 FGFR1 ITGAV [illegible] |
| HALLMARK_BILE_ACID_METABOLISM | 0.26535088 | 0.78477444 | 0.30238505 | 1.08715688 | 120 | 77 | AMACR CH25H [illegible] |
| HALLMARK_COAGULATION | 0.29821429 | 0.78477444 | -0.3089749 | -1.0841126 | 166 | 80 | C8A CFB GDA [illegible] |
| HALLMARK_COMPLEMENT | 0.26510067 | 0.78477444 | -0.2825973 | -1.0926208 | 157 | 160 | GNG2 GZMA [illegible] |
| HALLMARK_GLYCOLYSIS | 0.2251816 | 0.78477444 | 0.27729578 | 1.11052484 | 92 | 150 | LDHC CTH TPBG [illegible] |
| HALLMARK_KRAS_SIGNALING_DN | 0.26824818 | 0.78477444 | -0.3320997 | -1.117476 | 146 | 64 | TAS2R4 GRID2 [illegible] |
| HALLMARK_APICAL_SURFACE | 0.39319471 | 0.84026334 | -0.3821672 | -1.0584501 | 207 | 25 | MAL CROCC [illegible] |
| HALLMARK_HEDGEHOG_SIGNALING | 0.4033264 | 0.84026334 | 0.39021237 | 1.05742925 | 193 | 19 | CELSR1 NRP2 [illegible] |
| HALLMARK_IL2_STAT5_SIGNALING | 0.39162562 | 0.84026334 | 0.25439767 | 1.02393767 | 158 | 160 | SPP1 AMACR IRF6 [illegible] |
| HALLMARK_PEROXISOME | 0.37694013 | 0.84026334 | 0.27841617 | 1.02313659 | 169 | 83 | FADS1 SLC25A4 [illegible] |
| HALLMARK_WNT_BETA_CATENIN_SIGNALING | 0.40167364 | 0.84026334 | 0.3444153 | 1.04241753 | 191 | 31 | FZD1 CTNNB1 [illegible] |
| HALLMARK_INTERFERON_ALPHA_RESPONSE | 0.43369176 | 0.86738351 | -0.2837008 | -1.0082874 | 241 | 93 | IFITM1 SELL ISG20 [illegible] |
| HALLMARK_IL6_JAK_STAT3_SIGNALING | 0.47413793 | 0.89869281 | 0.27345299 | 0.97164005 | 219 | 70 | CXCL10 SOCS3 [illegible] |
| HALLMARK_NOTCH_SIGNALING | 0.48529412 | 0.89869281 | 0.33983167 | 0.98564627 | 230 | 26 | FZD1 FZD7 JAG1 [illegible] |
| HALLMARK_UV_RESPONSE_UP | 0.51807229 | 0.92512909 | 0.24787412 | 0.96388051 | 214 | 125 | CDKN2B NR4A1 [illegible] |
| HALLMARK_INFLAMMATORY_RESPONSE | 0.57894737 | 0.99818512 | -0.2496713 | -0.9544596 | 340 | 151 | CXCR6 IFITM1 [illegible] |
| HALLMARK_ADIPOGENESIS | 0.74468085 | 1 | 0.21894968 | 0.89985631 | 314 | 174 | COL15A1 DHCR7 [illegible] |
| HALLMARK_APICAL_JUNCTION | 0.71929825 | 1 | -0.2422273 | -0.888083 | 409 | 115 | GNAI1 NRTN [illegible] |
| HALLMARK_APOPTOSIS | 0.86240786 | 1 | 0.21659576 | 0.84802915 | 350 | 133 | CTH ATF3 [illegible] |
| HALLMARK_DNA_REPAIR | 0.95971564 | 1 | 0.19360745 | 0.77369538 | 404 | 142 | GSDME SNAPC4 [illegible] |
| HALLMARK_E2F_TARGETS | 0.67865707 | 1 | 0.22420641 | 0.92116333 | 282 | 176 | GINS4 ORC6 [illegible] |
| HALLMARK_ESTROGEN_RESPONSE_LATE | 0.91452991 | 1 | -0.2163963 | -0.8020266 | 534 | 120 | TFF3 FAM102A [illegible] |
| HALLMARK_FATTY_ACID_METABOLISM | 0.97560976 | 1 | 0.19128942 | 0.74876456 | 399 | 131 | FASN HMGCS1 [illegible] |
| HALLMARK_G2M_CHECKPOINT | 0.65110565 | 1 | 0.22887116 | 0.92820683 | 264 | 165 | MARCKS CENPA [illegible] |
| HALLMARK_HEME_METABOLISM | 0.99282297 | 1 | 0.17306912 | 0.6925827 | 414 | 145 | RNF123 C3 [illegible] |
| HALLMARK_INTERFERON_GAMMA_RESPONSE | 0.9681742 | 1 | -0.1954737 | -0.765372 | 577 | 181 | BANK1 GZMA [illegible] |
| HALLMARK_MYC_TARGETS_V1 | 1 | 1 | -0.1186215 | -0.4657435 | 605 | 184 | CDC45 CCNA2 [illegible] |
| HALLMARK_MYC_TARGETS_V2 | 1 | 1 | 0.15623894 | 0.53180683 | 459 | 56 | MAP3K6 TMEM97 [illegible] |
| HALLMARK_MYOGENESIS | 0.71826087 | 1 | -0.2444301 | -0.8911229 | 412 | 111 | COL6A2 FXYD1 [illegible] |
| HALLMARK_OXIDATIVE_PHOSPHORYLATION | 1 | 1 | -0.0942788 | -0.3716937 | 601 | 200 | ATP6V0C NDUFA7 [illegible] |
| HALLMARK_P53_PATHWAY | 0.84352078 | 1 | 0.21448567 | 0.86940761 | 344 | 164 | CDKN2B ATF3 [illegible] |
| HALLMARK_PANCREAS_BETA_CELLS | 0.748 | 1 | 0.32807359 | 0.78850303 | 373 | 12 | LMO2 VDR MAFB [illegible] |
| HALLMARK_PI3K_AKT_MTOR_SIGNALING | 0.96832579 | 1 | 0.19816486 | 0.73662922 | 427 | 89 | PIK3R3 PDK1 [illegible] |
| HALLMARK_PROTEIN_SECRETION | 0.96153846 | 1 | 0.203262 | 0.75557658 | 424 | 89 | TOM1L1 SSPN [illegible] |
| HALLMARK_REACTIVE_OXYGEN_SPECIES_PATHWAY | 0.95019157 | 1 | -0.2204216 | -0.6967713 | 495 | 48 | SRXN1 IPCEF1 [illegible] |
| HALLMARK_TGF_BETA_SIGNALING | 0.79791667 | 1 | 0.24832414 | 0.82773285 | 382 | 48 | CTNNB1 SERPINE1 [illegible] |
| HALLMARK_UNFOLDED_PROTEIN_RESPONSE | 0.99065421 | 1 | 0.18203252 | 0.69232519 | 423 | 105 | ATF3 EDC4 WFS1 [illegible] |

**CXCL10+ AM**

| | | | | | | | |
|---|---|---|---|---|---|---|---|
| HALLMARK_ANDROGEN_RESPONSE | 0.00224719 | 0.0234192 | 0.4562652 | 1.75057551 | 0 | 77 | CDK6 PMEPA1 [illegible] |
| HALLMARK_CHOLESTEROL_HOMEOSTASIS | 0.00218341 | 0.0234192 | 0.63692639 | 2.38223647 | 0 | 66 | SREBF2 MVK [illegible] |
| HALLMARK_INTERFERON_ALPHA_RESPONSE | 0.00224215 | 0.0234192 | 0.52767819 | 2.10280592 | 0 | 95 | CXCL11 CMTR1 [illegible] |
| HALLMARK_INTERFERON_GAMMA_RESPONSE | 0.00234192 | 0.0234192 | 0.49768075 | 2.15441696 | 0 | 185 | IDO1 ITGB7 [illegible] |
| HALLMARK_MTORC1_SIGNALING | 0.00231481 | 0.0234192 | 0.38191957 | 1.64924506 | 0 | 175 | PSPH HMGCS1 [illegible] |
| HALLMARK_BILE_ACID_METABOLISM | 0.00867679 | 0.07230658 | 0.40970339 | 1.55054526 | 3 | 69 | PEX11G ABCA5 [illegible] |
| HALLMARK_KRAS_SIGNALING_UP | 0.01882353 | 0.13445378 | 0.33593783 | 1.39506774 | 7 | 126 | SPP1 PCSK1N [illegible] |
| HALLMARK_G2M_CHECKPOINT | 0.12322275 | 0.68457083 | 0.28179454 | 1.19084045 | 51 | 150 | SLC12A2 CCNB2 [illegible] |
| HALLMARK_PROTEIN_SECRETION | 0.11777778 | 0.68457083 | 0.31256589 | 1.23419436 | 52 | 88 | ICA1 ARFGEF2 [illegible] |
| HALLMARK_ESTROGEN_RESPONSE_LATE | 0.14981949 | 0.69292487 | -0.3046722 | -1.1806496 | 82 | 102 | PERP DUSP2 NAB2 [illegible] |
| HALLMARK_IL6_JAK_STAT3_SIGNALING | 0.16630197 | 0.69292487 | 0.3125397 | 1.18287416 | 75 | 70 | CXCL11 CXCL10 [illegible] |
| HALLMARK_KRAS_SIGNALING_DN | 0.15789474 | 0.69292487 | 0.34143066 | 1.21237979 | 71 | 52 | EDN1 CYP39A1 [illegible] |
| HALLMARK_P53_PATHWAY | 0.19093079 | 0.70194384 | 0.26258978 | 1.11872061 | 79 | 157 | VDR NDRG1 TP53 [illegible] |
| HALLMARK_TGF_BETA_SIGNALING | 0.19654428 | 0.70194384 | 0.34447286 | 1.19509235 | 90 | 46 | PMEPA1 BMPR2 [illegible] |
| HALLMARK_ADIPOGENESIS | 0.29705215 | 0.81993313 | 0.24534517 | 1.06141074 | 130 | 171 | DHCR7 MIGA2 [illegible] |
| HALLMARK_COAGULATION | 0.3430127 | 0.81993313 | -0.2890514 | -1.0645264 | 188 | 72 | FYN RAPGEF3 [illegible] |
| HALLMARK_E2F_TARGETS | 0.24824356 | 0.81993313 | 0.25438702 | 1.08541759 | 105 | 160 | TIPIN UBE2T [illegible] |
| HALLMARK_MITOTIC_SPINDLE | 0.34027778 | 0.81993313 | 0.24077815 | 1.03253885 | 146 | 166 | TBCD CCNB2 [illegible] |
| HALLMARK_MYOGENESIS | 0.33111111 | 0.81993313 | 0.26687166 | 1.05852874 | 148 | 91 | KLF5 MYOM2 [illegible] |
| HALLMARK_PANCREAS_BETA_CELLS | 0.26482213 | 0.81993313 | 0.49392641 | 1.19912992 | 133 | 11 | VDR AKT3 SRP9 [illegible] |
| HALLMARK_TNFA_SIGNALING_VIA_NFKB | 0.35652174 | 0.81993313 | -0.2506922 | -1.0404661 | 204 | 176 | PTX3 TNFSF9 [illegible] |
| HALLMARK_UV_RESPONSE_UP | 0.36077058 | 0.81993313 | -0.2575053 | -1.0298917 | 205 | 121 | CDK2 CDKN1C [illegible] |
| HALLMARK_APICAL_JUNCTION | 0.38202247 | 0.83048363 | 0.25227885 | 1.01809836 | 169 | 103 | MADCAM1 VCAM1 [illegible] |
| HALLMARK_HEDGEHOG_SIGNALING | 0.47638604 | 0.88985089 | 0.38197321 | 0.99935676 | 231 | 16 | CDK6 PML NF1 [illegible] |
| HALLMARK_INFLAMMATORY_RESPONSE | 0.45595855 | 0.88985089 | -0.2464022 | -1.0029965 | 263 | 142 | TNFSF9 SELL [illegible] |
| HALLMARK_SPERMATOGENESIS | 0.48051948 | 0.88985089 | 0.26346268 | 0.97998065 | 221 | 63 | PCSK1N SLC12A2 [illegible] |
| HALLMARK_WNT_BETA_CATENIN_SIGNALING | 0.44086022 | 0.88985089 | 0.33469999 | 1.00898984 | 204 | 27 | FZD1 TP53 NCOR2 [illegible] |
| HALLMARK_HYPOXIA | 0.52392344 | 0.93557758 | 0.23198951 | 0.97122044 | 218 | 137 | ISG20 ANKZF1 [illegible] |
| HALLMARK_ALLOGRAFT_REJECTION | 0.59569378 | 0.95119934 | 0.22572751 | 0.94500467 | 248 | 137 | PRKCB CD8B [illegible] |
| HALLMARK_ANGIOGENESIS | 0.61410788 | 0.95119934 | 0.33255465 | 0.90577374 | 295 | 18 | SPP1 FGFR1 [illegible] |
| HALLMARK_APICAL_SURFACE | 0.72097378 | 0.95119934 | -0.2913444 | -0.8114741 | 384 | 19 | SULF2 BRCA1 [illegible] |
| HALLMARK_APOPTOSIS | 0.6778043 | 0.95119934 | 0.22195499 | 0.91734512 | 283 | 124 | CD69 ISG20 GCH1 [illegible] |
| HALLMARK_EPITHELIAL_MESENCHYMAL_TRANSITION | 0.60352423 | 0.95119934 | 0.23597952 | 0.93517911 | 273 | 90 | SPP1 PMEPA1 FN1 [illegible] |
| HALLMARK_ESTROGEN_RESPONSE_EARLY | 0.59252669 | 0.95119934 | -0.2382085 | -0.9429361 | 332 | 111 | MYB FOXC1 [illegible] |
| HALLMARK_GLYCOLYSIS | 0.62884161 | 0.95119934 | 0.2230796 | 0.9386903 | 265 | 140 | GNE ISG20 [illegible] |
| HALLMARK_IL2_STAT5_SIGNALING | 0.69668246 | 0.95119934 | 0.21552964 | 0.91182026 | 293 | 148 | SPP1 CXCL10 [illegible] |
| HALLMARK_MYC_TARGETS_V2 | 0.74193548 | 0.95119934 | 0.23741651 | 0.85509732 | 344 | 54 | PRMT3 SLC19A1 [illegible] |
| HALLMARK_NOTCH_SIGNALING | 0.74115456 | 0.95119934 | -0.2776156 | -0.823751 | 397 | 25 | FZD7 FZD5 KAT2A [illegible] |
| HALLMARK_UV_RESPONSE_DN | 0.6916221 | 0.95119934 | -0.2323449 | -0.8975227 | 387 | 97 | FYN MT1E SNAI2 [illegible] |
| HALLMARK_PEROXISOME | 0.76200873 | 0.95251092 | 0.22301582 | 0.8624954 | 348 | 80 | DHCR24 EHHADH [illegible] |
| HALLMARK_COMPLEMENT | 0.88754325 | 1 | -0.1996431 | -0.8173876 | 512 | 153 | S100A12 FYN [illegible] |
| HALLMARK_DNA_REPAIR | 0.99828179 | 1 | -0.167797 | -0.6836371 | 580 | 141 | ADA POLL TAF1C [illegible] |
| HALLMARK_FATTY_ACID_METABOLISM | 0.98364486 | 1 | 0.17772139 | 0.74091598 | 420 | 128 | HMGCS1 DHCR24 [illegible] |
| HALLMARK_HEME_METABOLISM | 0.8791019 | 1 | -0.2028786 | -0.8258073 | 508 | 140 | RBM38 ARHGEF12 [illegible] |
| HALLMARK_MYC_TARGETS_V1 | 1 | 1 | -0.1038333 | -0.4332788 | 578 | 182 | CDK2 CSTF2 |
| HALLMARK_OXIDATIVE_PHOSPHORYLATION | 0.99503722 | 1 | 0.16079147 | 0.69758212 | 400 | 198 | MTRF1 NNT HTRA2 [illegible] |
| HALLMARK_PI3K_AKT_MTOR_SIGNALING | 0.996337 | 1 | -0.171861 | -0.6457426 | 543 | 83 | CDK2 CAMK4 E2F1 [illegible] |
| HALLMARK_REACTIVE_OXYGEN_SPECIES_PATHWAY | 0.96672828 | 1 | -0.1952292 | -0.6674726 | 522 | 48 | SRXN1 EGLN2 |
| HALLMARK_UNFOLDED_PROTEIN_RESPONSE | 0.99458484 | 1 | -0.1710259 | -0.6627505 | 550 | 102 | ALDH18A1 DNAJA4 [illegible] |
| HALLMARK_XENOBIOTIC_METABOLISM | 0.99310345 | 1 | 0.16817 | 0.7036883 | 431 | 130 | GCH1 CYFIP2 [illegible] |
