## Supplementary material for "Type I IFN-activated lung monocytes and macrophages as initiators and drivers of fibrosis at the alveolar barrier in IPF": Suppl Table 5B

**Supplementary Table 5B.** Analysis of top 20 differentially active regulons comparing IPF to HC, for BAL AMs.
Top 20 statistically significant regulons with highest regulon activity score in IPF for each Oxford Cohort A BAL AM cluster were identified, and arranged as follows - 1. Arange regulons for each cell type by adjusted p value for FC comparing IPF to HC and exclude those with adjusted p <0.05; 2. Rank the by fold change (Log2FC) comparing IPF to HC ; 3. Cull list to top 20 Regulons acoording to FC ; 4. Identify those which are shared across all cell types.

| **FABP4$^{hi}$ AM** | **FABP4hi AM_log2FC** | **FABP4hi AM_Padj** | **IGF1$^{+}$ AM** | **IGF1+ AM_log2FC** | **IGF1+ AM_Padj** | **CD206$^{hi}$ FN1$^{hi}$ AM** | **CD206hi FN1hi AM _log2FC** | **CD206hi FN1hi AM_Padj** | **CXCL10$^{+}$ AM** | **CXCL10$^{+}$ AM_log2FC** | **CXCL10$^{+}$ AM_Padj** | ***mono-SPP1*$^{+}$** | ***mono-SPP1+_log2FC*** | ***mono-SPP1+_Padj*** |
|---|---|---|---|---|---|---|---|---|---|---|---|---|---|---|
| ATF3... | 0.01778334 | 2.26E-40 | BHLHE40... | 0.04619853 | 9.66E-52 | BHLHE40... | 0.04637516 | 5.59E-41 | ATF2... | 0.02086348 | 0.00130449 | ATF2... | 0.02741867 | 4.20E-09 |
| BHLHE40... | 0.03377355 | 2.22E-30 | CBFB... | 0.09877034 | 5.89E-32 | CBFB... | 0.08665589 | 1.01E-27 | BHLHE40... | 0.03792456 | 2.35E-12 | BHLHE40... | 0.03739623 | 3.94E-16 |
| CBFB... | 0.08051238 | 5.40E-28 | CREM... | 0.02080163 | 3.44E-14 | CEBPA... | 0.02066146 | 2.53E-27 | CBFB... | 0.10905204 | 4.96E-19 | CBFB... | 0.07374033 | 1.03E-10 |
| ELF1... | 0.02423272 | 1.29E-50 | E2F4... | 0.02751106 | 0.0030386 | CREM... | 0.01917926 | 1.69E-13 | ELF1... | 0.0308626 | 7.18E-31 | CREM... | 0.02235691 | 4.49E-06 |
| FOS... | 0.02360779 | 2.58E-42 | ELF1... | 0.02439094 | 1.08E-55 | EGR2... | 0.02027299 | 2.17E-40 | ETV7... | 0.0440854 | 6.12E-11 | ELF1... | 0.025254 | 1.44E-42 |
| FOSB... | 0.01811276 | 8.25E-32 | ETS2... | 0.02363893 | 8.86E-45 | ELF1... | 0.02128103 | 8.20E-45 | FOS... | 0.02177559 | 4.81E-15 | ELK4... | 0.0212454 | 2.84E-10 |
| FOXO3... | 0.01924353 | 2.54E-13 | FOS... | 0.03162034 | 1.42E-71 | FOS... | 0.02707033 | 4.76E-49 | FOXO3... | 0.02275726 | 2.60E-11 | EP300... | 0.01987351 | 6.61E-16 |
| GTF2F1... | 0.01767651 | 5.68E-07 | FOSB... | 0.02304658 | 5.61E-55 | FOSB... | 0.02067954 | 3.27E-33 | IKZF2... | 0.02216367 | 3.33E-06 | ETS2... | 0.01902123 | 2.04E-10 |
| HIF1A... | 0.02230346 | 0.04276987 | HIF1A... | 0.02475661 | 0.00807137 | IKZF2... | 0.02933592 | 4.13E-23 | IRF2... | 0.03387035 | 4.54E-12 | FOS... | 0.02584356 | 3.05E-18 |
| JUN... | 0.01949216 | 5.27E-25 | IKZF2... | 0.02807235 | 5.98E-17 | JUN... | 0.01843098 | 2.50E-24 | IRF7... | 0.03154352 | 1.02E-18 | FOSB... | 0.02212274 | 2.04E-08 |
| KLF4... | 0.02099742 | 6.68E-30 | JUN... | 0.02194233 | 1.76E-44 | JUNB... | 0.02391282 | 9.57E-51 | JUN... | 0.02124495 | 1.60E-15 | FOXP1... | 0.01872444 | 3.67E-20 |
| NFIL3... | 0.01772154 | 1.84E-16 | KLF4... | 0.02038071 | 5.78E-41 | KLF4... | 0.02476977 | 1.42E-52 | KLF4... | 0.02111061 | 1.58E-16 | JUNB... | 0.02150619 | 7.30E-13 |
| NR1H3... | 0.01818513 | 7.89E-31 | LEF1... | 0.02603892 | 5.30E-06 | MAFF... | 0.02095543 | 2.04E-17 | MYC... | 0.02104661 | 9.44E-05 | KLF4... | 0.02232822 | 5.96E-20 |
| NR3C1... | 0.02091531 | 0.00564946 | MYC... | 0.02759913 | 3.45E-15 | MYC... | 0.03094174 | 9.00E-31 | NFIL3... | 0.02271941 | 5.77E-09 | MAF... | 0.02285824 | 2.58E-12 |
| PPARG... | 0.04639087 | 2.26E-10 | NFIL3... | 0.02710582 | 6.87E-42 | NFE2... | 0.02106512 | 5.86E-18 | PPARG... | 0.05160486 | 0.00020175 | MYC... | 0.03227411 | 1.94E-09 |
| SMAD7... | 0.04179694 | 1.41E-07 | SMAD7... | 0.07667704 | 4.20E-25 | PRDM5... | 0.03351116 | 3.26E-11 | SMAD7... | 0.04264971 | 0.00676589 | RREB1... | 0.02423935 | 8.31E-18 |
| SOX4... | 0.02066698 | 1.53E-26 | SREBF2... | 0.02834125 | 1.04E-31 | SMAD7... | 0.06055786 | 8.78E-14 | STAT1... | 0.02264089 | 2.26E-24 | SMAD7... | 0.05922502 | 1.85E-10 |
| STAT2... | 0.01917101 | 3.03E-39 | STAT2... | 0.02012354 | 4.45E-50 | SREBF2... | 0.04501736 | 4.55E-53 | STAT2... | 0.02867311 | 2.10E-26 | SREBF2... | 0.02260917 | 2.77E-19 |
| TCF7L2... | 0.07093806 | 4.73E-17 | TCF7L2... | 0.07480922 | 1.08E-21 | TCF7L2... | 0.10319662 | 3.15E-46 | STAT3... | 0.02125033 | 2.16E-07 | STAT5A... | 0.02351221 | 0.00363512 |
| XBP1... | 0.0194331 | 1.27E-47 | XBP1... | 0.02154795 | 3.56E-71 | XBP1... | 0.01963107 | 2.50E-57 | TCF7L2... | 0.10934199 | 2.11E-17 | TCF7L2... | 0.06208372 | 0.00093876 |

| **Regulons shared across all cell types** | **Regulons shared in 4 of 5 cell types** | **Regulons shared in 3 of 5 cell types** | **Regulons shared in 2 of 5 cell types** |
|---|---|---|---|
| BHLHE40... | FOSB... | STAT2... | HIF1A... |
| CBFB... | JUN... | XBP1... | PPARG... |
| ELF1... | NFIL3... | CREM... | ETS2... |
| FOS... | MYC... | IKZF2... | JUNB... |
| KLF4... | | SREBF2... | |
| SMAD7... | | | |
| TCF7L2... | | | |
