## Supplementary material for "Type I IFN-activated lung monocytes and macrophages as initiators and drivers of fibrosis at the alveolar barrier in IPF": Suppl Table 5C

**Supplementary Table 5C.** Functional analysis of regulons with increased activity in IPF, that are found only in the specified AM subset

| Transcription Factor | Overview Transcription Factor function, mechanisms of action and effects across all cell types | Known or proposed function in monocytes, macrophages or related myeloid cells reported in literature | Summary of transcription factor function in macrophages |
|---|---|---|---|
| **FABP4$^{hi}$** | | | |
| *ATF3...* | Activating trnascription facor 3 (ATF3) acts as a hub of the cellular adaptive-response network. Multiple extracellular signals, such as endoplasmic reticulum (ER) stress, cytokines, chemokines, and LPS, are connected to ATF3 induction. Can modulate a wide range of effects including metabolism, glucose and adipose homeostasis. In immune response acts as transcriptional repressor downregulating pro-inflammatory responses. (1) | The role of ATF3 in host defence is through the regulation of immune responses. In immune cells, such as macrophages, NK, Th2 lymphocyte and neutrophil cells, ATF3 is expressed at low levels. After activation of these immune cells by different signalling pathways initiated by TLRs, LPS, IFNα/β/γ, pneumolysin, IL-12, anti-CD28 antibody and ovalbumin. ATF3 is upregulated (1-2). Subsequently, ATF3 downregulates the expression of target genes, including cytokines (IL-1β, IL-4, IL-5, IL-6, IL-12p40, IL-12b, IL-13, TNF, IFNβ/γ) and pro-apoptotic gene (Bak and Bax) by binding to the promoters (4). Mouse AFT3-/- BMDM display significantly lower basal and PRR-inducible IFN-β expression and AFT3 acts a transcriptional repressor with ATF3 binding to a regulatory region in the distal INFb promoter region that to control the magnitude of IFN-β promoter activity. ATF3 is also an ISG induced by T1 IFNs however modulates the expression of inflammatory ISGs thereby regulating the T1 IFN Inflammatory ISG response. (5) | Anti/counter-inflammatory effect, ATF3 is induced after pro inflammatory activation but works to repress the inflammatory response |
| *GTF2F1...* | General transcription facotr IIf subunit 1 (GTF2F1) is one of 3 subunits that makes up the TF II F. TF II F is a general transcription factor that along with other can stabilise RNA Pol II to enable gene transcription. i.e. not a specific Tf with a distinct function rather upregulates global transcription of genes | No additional studies in myeloid cells | |
| *NR1H3...* | Nuclear receptor Subfamily 1 Group H member 3 (NR1H3) Also known as Liver X receptor alpha (LXR alpha). Is inducible in macrophages, lung, liver, adipose tissue, kidney and gut. (Whilst LXR-beta is ubiquitously expressed). Play a central role in lipid metabolism including cholesterol uptake, efflux from cells, transport and excretion via the bile. In immune cells act as cholesterol sensors and can regulate inflammatory responses. Also can influence lipid and fatty acid metabolism via activating additional TFs including SREBP1c (the aster regulator of lipid metabolism and transport) and several other TFs including ABCA1 and fatty acid synthase (FAS). (6) | Considered a key regulator of macrophage homeostatic function in particular in relation to the phagocytosis of oxidized lipoprotein but also controlling transcriptional programs involved in lipid metabolism and inflammation . LXRs are critical in macrophage cholesterol efflux and regulating intracellular cholesterol levels and counteract the development of foam cells - this is via activating the TF ABCA1 after LXR ligands and LXR receptor complexes (7) and reverse cholesterol transport receptors APOE/APOC (8). LXRs will also repress pro-inflammatory gene expression on murine BMDM and RAW 2after stimulation with TNF and LPS and bacterial infection including iNOS, IL-6, IL1b and CCL2 and antagonises NFkB signalling (9) in vitro and in vivo. | Important in lipid metabolism, cholesterol efflux, is induced by efferocytosis and has a immune repressive effect esp. if activated following stim with pro inflammatory cytokines |
| *NR3C1...* | Nuclear Receptor subfamily 3 Group C member 1 (NR3C1) Encodes the glucocorticoid receptor (GR) , which upon glucocorticoid binding NR3C1 is translocated acting as a TF for glucocorticoid response elements thereby mediating a range of effects including i) upregulation of genes related to metabolism PCK1, TAT and G6PC, ii) trans repression via tethering of pro-inflammatory TF including NF-kB, AP-1 and STATs, resulting in repression of pro-inflammatory genes and pathways and iii) upregulation of immune regulators inc. IL-10. The effects and transcription programme of NR3C1 can be highly divergent dependent on differing isoforms and post translational modification, exposure duration, context and cell type although a degree of immune suppression via tethering of NF-kB is consistently and widely reported (10-11) | Endogenous glucocorticosteroids induce predominately anti-inflammatory / immuno-regulatory programmes via NR3C1 (Glucocorticoid receptor) in macrophages with a negative relationship between NR3C1 and TLR4, NF-kB and IFN receptor pathways. Upregulation of NR3C1 may result in cells to be more sensitive to the effects of endogenous gluococorticosteroids. Ligand activated NR3C1 TF in Mon-Macrophage effects include i) repression of activities of pro-inflammatory TFs including NFkB, AP-1 and IRF-3 via trans repression and tethering, ii) enhancing transcription of anti-inflammatory genes Dusp1, Anax1 and in myeloid cells Sphk1 (12, 13), and ii)in COVID19 myeloid NR3C1 expression negatively correlates with CXCL8 expression and neutrophil infiltration (14). However in certain contexts iv) GC-NR3C1 have been reported to induce proinflammatory effects including induction of NLRP3 transcription in monocyte like THP-1 cells (15), whilst chronic NR3C1 activation, with exogenous GCs, in macrophages results in downregulation of adaptive immune system activation but upregulation of innate immune pathways including chemokine expression and leucocyte recruitment (16). Finally, v) GC-NR3C1 promoted macrophage phagocytosis, efferocytosis and promotes wound healing and tissue repair (17) | Context specific response in Macrophages and can induce pro and anti inflammatory effects. More commonly associated with anti inflammatory effects in the literature |
| *SOX4...* | SRY-box transcription factor 4 (SOX4) is an essential developmental transcription factor that regulates stemness, differentiation, progenitor development, and multiple developmental pathways including PI3K, Wnt, and TGFβ signalling. Its expression is upregulated in response to TGF-b. Principally in epithelial and cancer cells promotes EMT | | |
| **IGF1$^{+}$ AM** | | | |
| *E2F4...* | E2F Transcription factor 4 (E2F4) belong to the family of E2F transcription factors involved in cell cycling and DNA replication. E2F4 is predominantly considered a transcriptional repressor whose activity is critical to engage and maintain cell cycle arrest in G0/G1. However in rapidly proliferating cells (inc Intestinal epithelial cells and tumour cells) can promote cell cycling and proliferation in a non canonical manner. | Specific function in macrophages is poorly described | unknown in macrophages |
| *LEF1...* | Lymphoid Enhancing Factor 1 (LEF1) plays a crucial role in mediating WNT/bCatenin signalling. It forms a complex with beta Catenin to promote transcription of genes downstream of WNT/bCatenin signalling. Expression of LEF1 is induced by ERG expression (in turn upregulated by TGF-b). Central component of organogenesis, tissue remodelling and specifically EMT. Associated with cancer where it is overexpressed. Also important In B cells important in differentiation of B progenitors to mature cells and proliferation (implicated in induction of AML). (18) | Multiple studies investigating LEF1 in AML / leukaemia and development of solid tumours but limited studies in the role of LEF1 in macrophages | unknown in macrophages |
| **CD206$^{hi}$ FN1$^{hi}$AM** | | | |
| *CEBPA...* | CCAAT Enhancer Binding Protein binding alpha (CEPBPA) is CCAAT enhancers family member with a wide range of functions. Involved in the differentiation and maturation of common myeloid progenitors to granulocyte / monocyte progenitors. Important in neutrophil maturation and terminal differentiation of neutrophils. CEBPA inhibits cell proliferation. CEBPA is downregulated in AML and mutations in CEBPA are thought to promote AML via several mechanisms (19). CEBPA can promote the transdifferentiating of lymphocytic cell types towards a myeloid state by upregulating the activity of PU.1 and CEPBb and upregulation of myeloid gene expression (20) | Limited information on the function of CEBPA in mature macrophages, upregulation of CEBPa resulted in downregulation of NFKB1, ISG15, CCR3, CCL4 and integrin ITGB1 amongst circulating mononuclear cells. Amongst tumour associated macrophages upregulation of CEFBA was associated with downregulation of genes associated with NF-kB, Type 1 interferon signalling, IL1B and STAT4 signalling although and Arg1 and NOS2 upregulation of genes associated with PGE2 signalling was observed suggesting CEFBA promoted a broad anti-inflammatory effect amongst tumour associated macrophages (21). | Broad anti-inflammatory effect but in TAMs where it has been studied |

| | | | |
|---|---|---|---|
| *MAFF...* | MAF BZIP Transcription Factor F (MAFF) belongs to the MAF family of transcription factors comprised of large MAF members and small MAF members including MAFF, MAFG and MAFK. Small MAFs can form homodimers with one another and exert a transcriptional repressive function or heterodimers with several other TFs including NFE2, NRF1, NRF2, NRF3. Thus MAFF proteins may have a wide range of effects depending on which TF they dimerise with. MAFF KO mice (all cells) do not have any specific phenotypic differences (22, 23). | No specific studies on MAFf KO and function in macrophages (nearly all studies are of c-MAF and MAFB which are large MAF TFs. MAFF heterodimers with NFE2 and can promote in transcriptional activity (see NfE2 below) | unknown in macrophages |
| *NFE2...* | Nuclear Factor Erythroid 2 (NFE2) transcription factor regulates expression of >250 oxidative/xenobiotic stress response genes expressing the anti-oxidant response element (ARE) binding site responsible for anti-oxidant homeostasis. Principal functions include i) protects against oxidative damage via ROS, ii) supress inflammation including via repression of the NLRP3 inflammasome, iii) promotes mitochondrial autophagy and degradation. Additionally NRF2 promoted erythroid and megakaryocyte poesis and differentiation (24). | NRF2 confers a anti-inflammatory phenotype in macrophages; macrophages from NRF2 -/- mice have increased ROS, activation of TLR, IRF-3 and NFkB signalling following LPS and increased pro-inflammatory cytokine production (25) via NADPH oxidase. The anti-inflammatory effects of NRF2 are mediated by both a Redox (anti-oxidant) dependent (26) and independent mechanism (25). In a latter study NRF2 was also shown to directly bind to sites in the proximity of several pro-inflammatory genes IL1a, IL1β and IL-6 and repress RNA pol mediated transcription (27). Specific increased NRF2 activity (by depletion of its repressor Keap1) in myeloid cells protects against caecal puncture induced sepsis, with lower mortality and pro-inflammatory marker expression compared to wild type and NRF2-/- models (28). | Suppresses pro-inflammatory and IFN signalling |
| *PRDM5...* | PR Set Domain 5 (PRDM5) is part of the zinc zipper PRDM family of TFs. PRDM5 like several members of the family plays a role in cell cycle and growth arrest and thought to confer a tumour suppressive function with overexpression promoting cell cycle arrest (29). Several tumour suppressive mechanisms have been suggested including antagonising bcatenin signalling. PRDM5 may also interact with GFI1 (growth factor independent 1) which is crucial in haematopoiesis with PDRM5 and GLI1 activating transcription. (30) | No specific studies investigating PDRM5 function in macrophages however Osteoblasts a related cell type with shared progenitors) PRDM5 is recognised to play a central role in promoting the transcription and expression of Collagen 1. PRDM5-/- osteoblastic cell lines and primary osteoblasts from PRDM5-/- mice have reduced Col1A (and Decron) mRNA expression whilst murine CHIpseq analysis revealed PRDM5 motifs upstream of 42 of 43 collagen genes with increased enrichment of PRDM5 targets in COL1A1 and COL1A2. Whilst genes targeted by PRDM5 are enriched for signalling pathways related to collagen fibril organisation and ECM organisation (31). | Specific function in macrophages not explored but in related osteoblasts promotes a pro fibrogenic effect |
| **CXCL10$^+$ AM** | | | |
| *ETV7...* | ETS Variant Transcription factor 7 (ETV7) is an ISG with expression upregulated by exposure to T1 and T2 IFNs and viral infection in line with IRF-1, STAT1 and NF-kB (32). ETV7 confers an immune regulatory effect supressing the expression of a subset of ISGs following T1 IFN (33) where overexpression of ETV7 in vitro in HEK293 or Lung epithelial cells resulted in repression of ISGs whilst functioETV7 loss enhances anti-viral ISG expression and reduced IFN mediated control of several strains of influenza virus. | ETV7 is an ISG upregulated in response to T1 IFNs and serves to regulate the Interferon / ISG repones although this has not been specifically studied in macrophages | ISG upregulated with IFN signalling - primarily limits the IFN response |
| *IRF2...* | Interferon Regulatory Factor 2 (IRF2) is generally considered to function as a transcriptional repressor and expressed in most immune and non immune cells. Expression is induced in response to T1 and T2 INF signalling where it primarily opposes the activity of IRF1 via several described mechanisms including competing with IRF1 promotor regions and inhibiting IRF1 nuclear translocation and may also interfere with NFkB, STAT1 and IRF9 signalling (34-35). However T1 IFNs may restrict IRF2 promotor sequence binding however and in unstimulated cells IRF2 is involved in maintaining TLR3 gene expression and maintains access to TLR3 genes. (36) | Whilst considered a negative regulator of T1 IFN signalling by antagonising IRF1 signalling following exposure to INFb, IRF2 appears to exert a complex role on pro-inflammatory signalling following responses to LPS and TNFa, where PS / TNFa stimulated BMDM from IRF2-/- mice has reduced IL-1b, IL12, IL6 and INFg mRNA expression and protein secretion (37). Similarly in response to bacterial ligands IRF2 is upregulated and promotes INFa expression, whilst INFa, proinflammatory cytokine production were reduced in IRF2 silenced murine macrophages (38). These results highlight the effects of IRF2 is heterogenous and mixed varying according to the type and duration of stimulation. | ISG also a negative regulator of IFN response |
| *IRF7...* | Interferon Regulatory Factor 7 (IRF7) Considered the 'master regulator of T1 IFN induction' along with IRF3 with both IRFs promoting INFa/b production following activation of a range of pattern recognition receptors. IRF7 Is highly expressed in pDCs snf whilst IRF3 is responsible for INFa/b transcription in most cell types, IRF7 at low levels 'primes' INFa/b production and is (unlike IRF3) is upregulated following INFa/b production resulting in a feed forward of INF signalling. Is considered a positive regulator of T1 INF signalling (39). | IRF7 promotes T1 IFN production and can amplify the T1 IFN response however may modulate and repress other pro-inflammatory pathways following PRR signalling. In BMDM IRF7 was required to induce T1 IFN production and inhibit viral replication however induction of an IRF7 protein lacking transcriptional activity resulted in reduced NFkB signalling suggesting an anti-inflammatory effect of IRF7 not related to its transcriptional activity (40). Similarly IRF7 may polarise monocytes and BMDM macrophages towards an anti-inflammatory phenotype with increase in pro-inflammatory signalling in IRF7 silenced BMDM. Here authors also highlighted IRF7 expression is upregulated in response to TGF-b. (41). | Promoted T1 IFN signalling and fine tune the inflammatory response towards an antiviral response by downregulating associated pro-inflammatory cytokine expression |
| *STAT1...* | Signal Transducer And Activator Of Transcription 1 (STAT1) Transduces signals from T1, II and III INFs to the nuclease. Phosphorylated STAT1 homo and heterodimers (with STAT2) binds to gamma activated sequences (GAS) and Interferon stimulated response element (ISRE) motifs in promotor regions of both pro-inflammatory and anti viral ISGs mediating a wide range of effects including cell activation, differentiation and innate immunity. Also transduces signals of IL-12 and IL-12 associated cytokines IL23,27 and 35. | Transduces and promotes T1 and T2 Interferon signalling and promotes proinflammatory cytokine expression following both INFg and LPS. Promotes an M1 like macrophage phenotype and antimicrobial and antiviral functions (42, 43). | Critical mediator of T1 and T2 INF signalling promoting pro-inflammatory and anti viral effects of INF signalling. Also enhances INF signalling in feed forward loop |
| *STAT3...* | Signal Transducer And Activator Of Transcription 3 (STAT3) Transduces signals from the cell surface to nuclease of a wide range of cytokines and growth factors including, T1 and T2 Interferons, IL-5, IL-6, IL-10, IL-12, IL-23 and growth factors including G-CSF, GM-CSF, EGF, FGF and PDGF exerting a wide range of effects depending on the cytokine and environmental context. | STAT3 mediates both pro-inflammatory signalling and gene transcription for example in response to IL6 and anti-inflammatory effects of IL-10 in addition to interacting with a wife range of additional signalling pathways with >40 ligands. Therefore the effect of STAT3 signalling on macrophage function has resulted in many wide ranging and contradictory conclusions. STAT3 activation and signalling is associated with promoting both pro and anti-inflammatory effects. In addition associated with promoting and mediating fibrotic process in cancer and both pulmonary and renal fibrosis. The effect of STAT3 on influencing macrophage phenotypes can be variable and is in part dependent on the stimuli, macrophage population studied and the interplay of several interacting signalling pathways (44). Specifically following T1/T2 IFN signalling of viral infection STAT3 activation and nuclear translocation results in expression of IL-10 and restraining the INF response via an number of mechanisms (45, 46). | Mediates IFNAR signalling and limits the ISG response via upregulation of IL-10. Implicated as a profibrotic mediator also |
| **mono-SPP1$^+$ AM** | | | |

| | | | |
|---|---|---|---|
| *ATF2...* | Activating Transcrition Factor 2 (ATF2) Is a member of the AP1 family of TFs. It can interact with and form heterodimers with many AP1 family TFs (including AP1, cJun, cFos, CREB1, and consequently has been implicated in modulating a diverse range of cellular functions. The AT2 protein can also exist as a number of splice variants can interact with different AP1 family TFs along with different post translational modifications contributing to this functional diversity. ATF2 has been implicated in inducing and repressing cellular activities relating to cell cycling, cell death and immune mediated and inflammatory responses in response to a range of different stimuli (47). | In one study AT2F function on THP-1 cells reveal AT2F targets the PPM1A promotor which is upregulated in monocyte to macrophage differentiation, whilst overexpression of AT2F in THP-1 cells results in M1 like rounded circular inflammatory monocytes. AT2F overexpression in monocytes sensitised to INFg and LPS stimulation results in upregulation of HLADR expression, enhanced antibacterial function and enhanced IL-1b and CCL2 expression post Mtb infection, compared to control or AT2F -/- THP-1 cells. Additionally AT2F overexpression promoted increased glycolytic metabolism associated with pro-inflammatory macrophage states. Overall AT2F promotes monocyte to macrophage differentiation and enhances pro-inflammatory macrophage responses (48). ATF2 is also known to bind to the THF-a promoter with impaired TNF-a production in ATF2 -/- THP-1 cells following Mtb infection via a p38 MAPK dependent mechanism (49) and increased TNFa following LPS (50). | Promotes monocyte to macrophage differentiation, in THP-1 cells induces glycolysis, pro-inflammatory cytokine production and antibacterial properties. Overall a pro-inflammatory effect |
| *ELK4...* | ETS Transcription Factor 4 (ELK4( is a member of the ETS TF family and binds to the c-fos promoter region and a critical regulator of c-fos transcriptional expression so is a recognised proto-oncogene that promotes cell proliferation, malignant cell transformation and tumour progression. Additionally ELK4 has been implicated as an oncogene via c-fos independent mechanisms (51). | Single study has investigated function of ELK 4 in tumor associated macrophages. In TAMs ELK4 knockdown increased expression of proinflammatory cytokines IL-1b and TNFa and NOS whilst Arg1, Fizz1 and Ym1 and IL-10 were reduced indicating ELK4 promotes an M2 macrophage phenotype (52). The effects of ELK4 on macrophage function may also be context dependent. A separate study using murine peritoneal macrophages with ELK4 knockdown treated with zymosan from yeast resulted in transcription of genes relating to stress responses suggesting ELK4 may have a different function in macrophages following response to infection (53). | Limited studies but effects appear context dependent to a degree in macrophages but in TAMs induces an M2 anti-inflammatory phenotype with increase IL-10, Arg 1 and Fizz1 expression |
| *EP300...* | E1A Binding Protein P300 (EP300) Is a transcriptional coactivator protein. It binds to activated CREB protein and promotes transcription of genes related to cell proliferation and differentiation and has been implicated in the development of leukaemia and is tumorigenic. Additionally functions as a acetyltransferase and is recognised to acetylate nucleosome histones enabling it influence transcription at an epigenetic level. Finally can acetylate and act as a coactivator of a number of transcription factors resulting in a broad range of actions including response to heat shock (via heat shock transcription factor 1) and cellular stress as a co-activator of p53 signalling. (54, 55) and hypoxia as a co-activator to HIF1a (56). There is growing evidence of EP300 contributing to the development of fibrosis in multiple organs via a number of proposed mechanisms and promoting the expression of profibrotic mediators amongst fibroblasts and epithelial - mesenchymal transformation of epithelial cells and through promoting the effects of TGF-b signalling (57). | No specific studies investigating the role of EP300 in monocyte or macrophage subsets | No specific studies in macrophages but in other cell types appears to promote fibrosis |
| *FOXP1...* | Forkhead Box p1 (FOXP1) is a member of the fork head box transcription factor family. Acts primarily as a transcriptional repressor though modulates wide range of tissue and cell specific effects, i) is involved in the development and organogenesis of a range of tissues (including cardiac, muscle, neurological and lung airway development) and angiogenesis. ii) is involved in the maturation of both T and B cells and in regulatory T cels function, iii) can promote proliferation and pluripotency amongst embryonic stem cells and iv) modulate macrophage activity and range of principle macrophage functions. | FOXP1 is upregulated in monocytes and becomes downregulated during monocyte to macrophage differentiation FOXP1 regulates CSF1R expression where overexpression of FOXP1 reduces CSF1R cell surface levels. FOXP1 monocyte/macrophage overexpression reduces peritoneal macrophage cytokine (Il1b, IL-12, INFγ, CSF1, CCL1) expression in response to LPS, and reduced phagocytosis in vitro and reduced bacterial clearance in vivo compared to WT (58). Similarly hypoxia induces increased pro-inflammatory cytokine expression in RAW 264.7 cells with downregulation of HMGB1 and increased FOXP1 expression. Whilst knockdown of HMGB1 which reduces FOXP1 expression is associated with reduced pro-inflammatory cytokine expression post hypoxia stimulation. (59). Overall increased FOXP1 in mono-SPP1+ AMs in IPF could reflect i) monocyte abundance in this population or ii) an anti-inflammatory phenotype of macrophages in this cluster | Regulates CSF1R levels on monocytes |
| *MAF...* | MAF BZIP Transcription Factor (MAF) is a member of the large MAF transcription factor family. Also known as c-MAF. Acts as a transcriptional activator and repressor and confers a wide range of cell specific effects notably in tissue development and considered a pro-oncogene. Has a wide range of effects on many immune cell types including T cells where it may partner with additional transcription factors and modulate the effects of different T cell subsets (notably Th2, TFH and Th17 subsets although broadly promotes IL-10 production amongst T cells (60). | cMAF promotes an M2 like anti-inflammatory phenotype with MAF -/- resulting in increased pro-inflammatory cytokine expression, IL-12, IL1b, IL-6 and reduced expression of IL-10 but also significant impairment of factors involved in wound healing and repair including TGF-b, VEGF and Arginase. cMAF also induces CSF1R expression and cMAF blockade impairs CSF1R expression in murine BMDMs. cMAF expression is induced by IL-10 but not IL-4 or IL-13.(61,62). cMAF expression was increased in hypertrophic scar tissue whilst cMAF overexpression in RAW 264.7 cells cocultured with fibroblasts resulted in increased fibroblast proliferation, migration and ECM protein (COl1,3 and aSMA) compared to WT RAW 264.7 cells (63). | Promotes an anti-inflammatory profibrotic phenotype |
| *RREB1...* | Ras responsive element binding protein 1 (RREB1) is a transcription factor downstream of RAS-MAPK signalling and can act as a transcriptional activator and repressor. RREB1 is involved in several cellular processes including DNA damage and repair, cell growth and proliferation, differentiation, adipogenesis, fasting glucose balance, insulin production and calcium homeostasis and calcitonin and zinc uptake. It has been shown to promote p53 expression in response to UV radiation. Expression has been associated with the development of several cancers as downregulation is associated with the development of type 2 diabetes (64). RREB1 induced by RAS has been observed as a cofactor of SMAD transcription factors downstream of TGF-b signalling and inhibition of RREB1 prevented EMT in pancreatic epithelial cells and cancer models. Additionally RREB1 silencing downregulated ECM components (collagen, laminin and proteases) and genes associated with fibrosis (Wisp1, Ctgf, IL-11 andPDGFB) in both epithelial cells and fibroblasts. (65) | No studies investigating function in monocytes or macrophages | Not explored in macs but in other cell types promotes and pro-fibrotic phenotype, is upregulated by TGF-b signalling and through an unknown mechanism result in activation of neighbouring fibroblasts |
| *STAT5A...* | Signal Transducer And Activator Of Transcription 5A (STAT5A) and STAT5B proteins are 90% homologous with shared effectors function and both impact a range of cellular processes with compensatory redundant function. In non hematopoietic lineages respond to prolactin during mammary gland formation and lactation. In haematopoietic cells involved in myelopoiesis, lymphoid development, megakaryopoiesis and myeloid and granulocyte function. STAT5a/b are key signalling molecules downstream of a range of cytokines and GF's ( IL-2, -3, -4, -5, -7, -9, -13, -15, -21, EPO, thrombopoietin (TPO), GH, prolactin, stem cell factor (SCF), Flt3, CSF2 and CSF3. Therefore have a wide range of effects that are context dependent (66). | CSF2 mediated STAT5a activity and function has been studied and STAT5 regulation of monocyte / macrophage proliferation, differentiation and activation is recognised. In human BMDM STAT5A silencing resulted in reduced pro-inflammatory cytokine expression (IL-6, TNF-a and IL-8) following LPS and increased phagocytosis and reduced cholesterol uptake (the latter change associated with reduced PPARg transcription). STAT5A silencing also resulted in increased expression of apoptosis markers and macrophage proliferation compared to WT hBMDM in keeping with the known effects of CSF2 signalling on macrophage proliferation and survival. These findings suggest STAT5A (as (observed in CSF2 inhibition) increases pro-inflammatory cytokines secretion, and macrophage survival proliferation in addition promotes cholesterol and lipid uptake whilst supressing phagocytic activity (67). Similar findings are observed in murine BMDM where STAT5A KO resulted in reduced pro-inflammatory signalling with upregulation of anti-inflammatory gene sets as well as genes associated with fibrosis including Col1a1m Col5a2 and Vegfa (68). Suggesting STAT5a supresses genes associated with fibrosis. | STAT5A promotes pro-inflammatory signalling, antigen presentation (via effects of GM-CSF) with downregulation of pro-fibrotic gene sets. |

| **Abbreviations** | **Abbreviations** |
|---|---|
| ABCA1 | ATP Binding Cassette Subfamily A Member 1 |
| AML | Acute Myeloid Leukaemia |
| APOC | Apolipoprotein C |
| APOE | Apolipoprotein E |
| Arg1 | Arginase 1 |
| aSMA | alpha smooth muscle actin |
| BMDM | Bone Marrow Derived Macrophage |
| CCL | CC Motif Chemokine Ligand |
| COVID | Coronavirus |
| COX | Cyclooxygenase |

| | |
|---|---|
| CREB | CAMP Responsive Element Binding Protein |
| CSF2 | Colony Stimulating Factor 2 |
| CSF3 | Colony Stimulating Factor 3 |
| CXCL | CXC motif chemokine ligand |
| ECM | Extra Cellular Matrix |
| EGF | Epithelial Growth Facotr |
| EMT | Epithelial Mesenchymal Transition |
| ER | Endoplasmic Reticulum |
| FAS | Fatty Acid Synthetase |
| FGF | Fibroblast Growth Factor |
| Fizz-1 | Cyctseine Rich secreted protein |
| FLt3 | Fims receptor related Tyrosine Kinase 3 |
| G-CSF | Granulocyte Colony Stimulating Factor |
| G0 | Growth Phase 0 |
| G1 | Growth Phase 1 |
| G6PC | Glucose-6-Phosphatase Catalytic |
| GAS | Gamm activated Sequence |
| GM-CSF | Granulocyte Macrophage Colony Stimulating Factor |
| HIF1a | Hypoxia Inducible Factor 1 |
| IFNg | Interferon gamma |
| IL | Interleukin |
| INF | Interferon |
| INFb | Interferon beta |
| iNOS | inducible Nitric Oxide Synthase |
| IRF | Inteferon Regulatory Factor |
| ISG | Inteferon Stimulated Gene |
| ISRE | Inteferon Sensitive Response Element |
| JUN | JUN Proto-Oncogene |
| KO | Knock Out |
| LPS | Lipopolysacchiride |
| LXR | Liver X Receptor |
| NFkB | Nuclear Factor kappa B |
| NK | Natural Killer |
| PCK1 | Phosphoenolpyruvate carboxykinase 1 |
| PDGF | Platlet Derived Growth Factor |
| PDGFB | Platlet Derived Growth Factor Beta |
| PI3K | Phosphoinositide-3-Kinase |
| SCF | Stem Cell Factor |
| SphK1 | SPHK1 Interactor, AKAP Domain Containing |
| TAT | Tyrosine Aminotransferase |
| TF | Trancription Factor |
| TGF-b | Transforming Grwoth Factor b |
| Th | T helper |
| TNF | Tumor Necrosis Factor |
| VEGF | Vascular Endothelial Growth Facotr |
| WISP | WNT1-inducible-signaling pathway protein genes |
| WT | Wild Type |
