## Supplementary material for "Type I IFN-activated lung monocytes and macrophages as initiators and drivers of fibrosis at the alveolar barrier in IPF": Suppl Table 6

**Details of antibodies used in suspension mass CyTOF**

| | Marker | Clone | Metal | Cat no | Vendor | RRID | Conjugated In house? | Diltuion |
|---|---|---|---|---|---|---|---|---|
| CD45 (HI30) - 89Y | CD45 | HI30 | 89Y | 3089003B | Standard Biotools | AB_2938863 | No | 1:100 |
| CD14 (M5E2) - 114Cd | CD14 | M5E2 | 114Cd | 301843 | BioLegend | AB_2562813 | Yes | 1:100 |
| CD11c (6.9) - 141Pr | CD11c | 6.9 | 141Pr | 301602 | BioLegend | AB_314172 | Yes | 1:100 |
| CD11b (ICRF44) - 142Nd | CD11b | ICRF44 | 142Nd | 301337 | BioLegend | AB_2562811 | Yes | 1:200 |
| CD45RA (HI100) - 143Nd | CD45RA | HI100 | 143Nd | 304102 | BioLegend | AB_314406 | Yes | 1:50 |
| HLA-DR (L243) - 144Nd | HLA-DR | L243 | 144Nd | 307602 | BioLegend | AB_314680 | Yes | 1:400 |
| CD4 (RPA-T4) - 145Nd | CD4 | RPA-T4 | 145Nd | 300570 | BioLegend | AB_2810427 | Yes | 1:800 |
| CD19 (HIB19) - 146Nd | CD19 | HIB19 | 146Nd | 302247 | BioLegend | AB_2562815 | Yes | 1:100 |
| CD20 (2H7) - 147Sm | CD20 | 2H7 | 147Sm | 302343 | Biolegend | might need to request | Yes | 1:200 |
| CCR6 (G034E3) - 148Nd | CCR6 | G034E3 | 148Nd | 353401 | BioLegend | AB_10918626 | Yes | 1:100 |
| CD56 (QA18A21) - 149Sm | CD56 | QA18A21 | 149Sm | 398802 | BioLegend | AB_2832780 | Yes | 1:50 |
| pSTAT5 (Y694) - Nd150 | pSTAT5 | Y694 | 150Nd | 3150005A | Standard Biotools | AB_2744690 | No | 1:50 |
| CD45RO (UCHL1) - 151Eu | CD45RO | UCHL1 | 151Eu | 304239 | BioLegend | AB_2563752 | Yes | 1:100 |
| CD27 (O323) - 152Sm | CD27 | O323 | 152Sm | 302839 | BioLegend | AB_2562817 | Yes | 1:100 |
| pSTAT1 (Y701) - 153Eu | pSTAT1 | Y701 | 153Eu | 3153003A | Standard Biotools | AB_2811248 | No | 1:50 |
| CD1c (L161) - 154Sm | CD1c | L161 | 154Sm | 331502 | BioLegend | AB_1088995 | Yes | 1:100 |
| CD123 (6H6) - 155Gd | CD123 | 6H6 | 155Gd | 306027 | BioLegend | AB_2562823 | Yes | 1:200 |
| pp38 (T180/Y182) - 156Gd | pp38 | T180/Y182 | 156Gd | 3156002A | Standard Biotools | AB_2661826 | No | 1:50 |
| pSTAT3 (Y705) - 158Gd | pSTAT3 | Y705 | 158Gd | 3158005A | Standard Biotools | AB_2811100 | No | 1:50 |
| pMAPKAPK2 (T334) - 159Tb | pMAPKAPK2 | T334 | 159Tb | 3159010A | Standard Biotools | might need to request | No | 1:50 |
| CD3 (UCHT1) - 160Gd | CD3 | UCHT1 | 160Gd | 300438 | BioLegend | AB_11146991 | Yes | 1:100 |
| DNGR1 (8F9) - 161Dy | DNGR1 | 8F9 | 161Dy | 353802 | BioLegend | AB_10983070 | Yes | 1:50 |
| IFNAR2 (MMHAR-2) - 162Dy | IFNAR2 | MMHAR-2 | 162Dy | NBP2-89447 | Novus Bio | AB_3435134 | Yes | 1:50 |
| STAT1 (246523) - 163Dy | STAT1 | 246523 | 163Dy | MAB1490 | R AND D SYSTEMS | AB_2159797 | Yes | 1:50 |
| CD161 (HP-3G10) - 165Ho | CD161 | HP-3G10 | 165Ho | 339919 | BioLegend | AB_2562836 | Yes | 1:400 |
| pNFkBp65 (S529) - 166Er | pNFkBp65 | S529 | 166Er | 3166006A | Standard Biotools | AB_2847867 | No | 1:50 |
| CCR7 (150503) - 167Er | CCr7 | 150503 | 167Er | MAB197 | R AND D SYSTEMS | AB_2072803 | Yes | 1:100 |
| pSTAT6 (Y641) - 168Er | pSTAT6 | Y641 | 168Er | 3149004A | Standard Biotools | might need to request | No | 1:50 |
| CD64 (10.1) - 169Tm | CD64 | 10.1 | 169Tm | 305029 | BioLegend | AB_2563759 | Yes | 1:100 |
| CD141 (M80) - 170Er | CD141 | M80 | 170Er | 344102 | BioLegend | AB_2201808 | Yes | 1:100 |
| pERK12 (D13.14.4E) - 171Yb | pERK12 | D13.14.4E | 171Yb | 3171010A | Standard Biotools | AB_2811250 | No | 1:50 |
| CD38 (HIT2) - 172Yb | CD38 | HIT2 | 172Yb | 303502 | BioLegend | AB_314354 | Yes | 1:100 |

| | | | | | | | | |
|---|---|---|---|---|---|---|---|---|
| STAT3 (15H2B45) - 173Yb | STAT3 | 15H2B45 | 173Yb | 371802 | BioLegend | AB_2629732 | Yes | 1:50 |
| pSTAT4 (Y693) - 174Yb | pSTAT4 | Y693 | 174Yb | 3148006A | Standard Biotools | AB_2811098 | No | 1:50 |
| CCR4 (205410) - 175Lu | CCR4 | 205410 | 175Lu | MAB1567-10 | R AND D SYSTEMS | | Yes | 1:50 |
| CXCR3 (G025H7) - 176Yb | CXCR3 | G025H7 | 176Yb | 353702 | BioLegend | AB_10983073 | Yes | 1:100 |
| CD8 (RPA-T8) - 198Pt | CD8 | RPA-T8 | 198Pt | 301074 | BioLegend | AB_2814117 | Yes | 1:200 |
| CD16 (3G8) - 209Bi | CD16 | 3G8 | 209Bi | 3209002B | Standard Biotools | | No | 1:100 |

**Other reagents**

| | | |
|---|---|---|
| Water, LiChrosolv grade | Merck | 1-15333.2500 |
| Methanol, LiChrosolv grade | Merck | 1.06035.2500 |
| Benzonase | Thermo Scien | Cat# 88700; RRID: N/A |
| Maxpara Cell Staining Buffer | Fluidigm | Cat# 201068; RRID: N/A |
| Maxpara Nuclear Antigen Staining Buffer Set | Fluidigm | Cat# 201063; RRID: N/A |
| Cell-ID Cisplatin Pt198 | Fluidigm | Cat# 201198; RRID: N/A |
| Cell-ID Intercalator-Ir | Fluidigm | Cat# 201192B; RRID: N/A |
| EQ Four Element Calibration Beads | Fluidigm | Cat# 201078; RRID: N/A |
| Maxpar Cell Acquisition Solution (CAS) | Fluidigm | Cat# 201240; RRID; N/A |
| Maxpar Water | Fluidigm | Cat# 201069; RRID: N/A |
| FcX | BioLegend | Cat# 422301 |
| Cell-ID 20-Plex Pd Barcoding Kit | Fluidigm | Cat# 201060; RRID: N/A |
| Cell-ID 20-Plex Pd Barcoding Kit | Fluidigm | Cat# 201060; RRID: N/A |
| Maxpar X8 Multimetal Labeling Kit (40 rxn) | Fluidigm | Cat# 201300; RRID: N/A |
