## Supplementary material for "Type I IFN-activated lung monocytes and macrophages as initiators and drivers of fibrosis at the alveolar barrier in IPF": Suppl Table 7

**Supplementary Table 7. Primer list of RT-qPCR assay.**

| Gene | Accession number | Forward primer | Reverse primer | Amplicon length | Protein Transcript |
|---|---|---|---|---|---|
| STAT1 | NM_007315.3 | Taqman | Taqman | 66 | signal transducer and activator of transcription 1 |
| IRF7 | NM_001572.5 | Taqman | Taqman | 77 | interferon regulatory factor 7 |
| MX1 | NM_001144925.2 | Taqman | Taqman | 53 | MX dynamin like GTPase 1 |
| MX2 | NM_002463.2 | Taqman | Taqman | 72 | MX dynamin like GTPase 2 |
| ISG15 | NM_005101.4 | Taqman | Taqman | 100 | ISG15 ubiquitin-like modifier |
| IFI44L | NM_006820.4 | Taqman | Taqman | 78 | interferon induced protein 44 like |
| OASL | NM_003733.3 | Taqman | Taqman | 107 | 2'-5'-oligoadenylate synthetase like |
| RSAD2 | NM_080657.4 | Taqman | Taqman | 76 | radical S-adenosyl methionine domain containing 2 |
| IFNB1 | NM_002176.3 | Taqman | Taqman | 73 | interferon beta 1 |
| GAPDH | NM_001256799.2 | Taqman | Taqman | 157 | Glyceraldehyde 3- phosphate dehydrogenase |
| STAT6 | NM_001178078.1 | Taqman | Taqman | 96 | signal transducer and activator of transcription 6 |
